# Genetic influences on the intrinsic and extrinsic functional organizations of the cerebral cortex

**DOI:** 10.1101/2021.07.27.21261187

**Authors:** Bingxin Zhao, Tengfei Li, Stephen M. Smith, Zirui Fan, Xiaochen Yang, Yilin Yang, Juan Shu, Di Xiong, Xifeng Wang, Yue Yang, Tianyou Luo, Ziliang Zhu, Yue Shan, Yujue Li, Zhenyi Wu, Heping Zhang, Yun Li, Jason L. Stein, Hongtu Zhu

## Abstract

The human cerebral cortex is a vital component of brain function, but the genetic influences on cortical functional organization remain poorly understood. In this study, we used a parcellation-based approach to process resting-state and task-evoked functional magnetic resonance imaging (fMRI) from over 48,000 individuals in UK Biobank and ABCD studies. We identified 47 loci associated with functional areas and networks at rest, 15 of which also affected functional connectivity during task performance. We observed patterns of heritability and locus-specific genetic effects across different brain functional areas and networks. Our findings suggest that specific functional areas and networks share genetic influences with cognition, mental health, and major brain disorders such as Alzheimer’s disease and schizophrenia. For example, the *APOE* ε4 locus strongly associated with Alzheimer’s disease was particularly associated with the visual cortex in the secondary visual and default mode networks in both resting and task fMRI. This study contributes to our understanding of the genetic determinants of cerebral cortex function by analyzing biobank-scale fMRI data in high-resolution brain parcellation. Additionally, it prioritizes genetically associated fMRI traits for specific brain disorders.

---

The human cerebral cortex is the largest part of the human brain and controls complex brain functions. Based on known functional and topographic specializations at different scales, the cerebral cortex of the human brain can be divided into distinct areas and networks, providing insight into the brain’s functional architecture^1,2^. To define such brain partitions, a few brain parcellations have been developed over the past decade^3^. In functional magnetic resonance imaging^4,5^ (fMRI), cerebral cortex functions can be evaluated by measuring functional connectivity (or correlation of blood-oxygen-level dependent [BOLD] activity) among multiple cortical areas along a given parcellation. In particular, resting-state fMRI captures the intrinsic functional organization of the cortex without any explicit stimuli, whereas task-evoked fMRI measures extrinsic cortical interaction and temporal synchrony in response to a specific task. A variety of clinical applications of both task-evoked and resting-state fMRI have revealed changes in brain function in multiple neurological and psychiatric disorders, such as schizophrenia^6,7^, Alzheimer’s disease^8^, Parkinson’s disease^9^, autism spectrum disorders^10^, and major depressive disorder (MDD)^11,12^.

Recently, growing literature suggested that fMRI measures may have lower test re-test reliability than imaging measures of brain structures^13,14^. As a result, a large sample size is particularly important in fMRI studies to generate reproducible findings^14,15^. For example, fMRI images from thousands of individuals might be needed in association analysis with human behavioral traits^14^. Moreover, although twin studies have consistently established that brain resting and task fMRI traits are moderately heritable^16–23^ (for example, the heritability range of resting fMRI was (0.2,0.6) in a recent review^24^), large imaging genetic cohorts are required to identify the specific genetic variants associated with the functional brain. Fortunately, a few big fMRI databases have become publicly available in recent years, including the Adolescent Brain Cognitive Development^25^ (ABCD) and the UK Biobank^26^ (UKB). Particularly, the UKB imaging study aimed to collect data from 100,000 subjects^27^, providing a new opportunity for fMRI research in one large-scale cohort. Using a whole brain spatial independent component analysis (ICA)^28–30^ approach in the UKB study, the narrow sense single-nucleotide polymorphism (SNP) heritability of resting fMRI traits was reported to be around 10% across the entire brain and higher than 30% in some functional regions^31^. A few genome-wide association studies (GWAS)^31–33^ have also been recently conducted on resting fMRI using these whole brain ICA-based traits. The whole brain ICA is a parcellation-free dimension reduction method that estimates the functional brain regions (i.e., ICA components/regions) directly from the fMRI data. Although the whole brain ICA is a powerful and popular fMRI tool, it is a data-driven method, which might limit its generalizability and interpretability^34^. Specifically, the whole brain ICA attempts to capture major variations in the data. As a result, ICA regions typically have large sizes, limiting their ability to capture high-resolution details of brain functionality. For example, an earlier study^30^ defined 55 ICA components in the UKB^35^ dataset, most of which are distributed across multiple brain areas and networks^32^. It may be difficult for ICA to prioritize specific brain networks and regions for specific brain disorders, limiting the use of fMRI phenotypes in clinical/translational research. Moreover, it might be difficult to use ICA to compare intrinsic and extrinsic functional architectures, since the ICA components estimated in resting and task fMRI may not be well-aligned.

To overcome these limitations, here we used a parcellation-based approach to provide fine-grained details about the genetic architecture of cerebral cortex functional organizations. A recently developed human brain parcellation^1^, which partitioned the cerebral cortex into 360 areas (referred to as the Glasser360 atlas hereafter, **Table S1** and **Fig. 1A**), was used to analyze resting and task fMRI data in the UKB study. The task implemented in the UKB fMRI study was an emotional processing task^36,37^, known to robustly activate the amygdala and visual systems. The Glasser360 atlas was constructed using high-quality multi-modality data from the Human Connectome Project (HCP^38^) and greatly improved the neuroanatomical resolution of human cerebral cortex annotations. The 360 cortical areas were grouped into 12 functional networks^39^, including four well-known sensory networks (the primary visual, secondary visual, auditory, and somatomotor), four cognitive networks (the cingulo-opercular, default mode, dorsal attention, and frontoparietal), the language network, and three recently identified networks (the posterior multimodal, ventral multimodal, and orbito-affective) (**Fig. 1A**). In addition to pairwise functional connectivity among areas, we developed a parcellation-based dimension reduction procedure to generate network-level fMRI traits via a combined principal component analysis (PCA) and ICA methods^31^ in a training-validation design (**Fig. S1**). Briefly, area-level functional connectivity pairs within each network and between each pair of networks were summarized into network-specific traits to quantify the strength of functional connectivity within each network and two different networks. These network-specific traits have been mapped back to the 360 cortical areas to facilitate better visualization and biological interpretation (Methods). Furthermore, we examined the amplitude, which was a heritable measure of brain activity^40^ used in previous fMRI GWAS^31–33^. Together, there were 8,531 area-level traits and 1,066 network-level traits for resting fMRI and 8,531 area-level traits and 919 network-level traits for task fMRI. Genetic architectures were examined at both area- and network-levels for brain functions using these fMRI traits. The large number of traits 1) enabled the comparison between intrinsic and extrinsic functional architectures using both resting and task fMRI; and 2) uncovered much more and finer detail on the genetic influences on specific functional areas and networks and their genetic links with brain-related complex traits and disorders. We also processed the resting fMRI data from the ABCD study as a replication dataset. Most of the ICA components used in previous resting fMRI GWAS^31–33^ overlapped with multiple Glasser360 areas and networks (**Figs. 1B** and **S2**). Thus, the current parcellation-based study provided more details and actionable findings for future clinical research. For example, we found that the *APOE* ε4 locus was particularly associated with the functional connectivity of the visual cortex in the secondary visual and default mode networks, prioritizing fMRI biomarkers for practical applications in Alzheimer’s disease research.

**Fig. 1.**
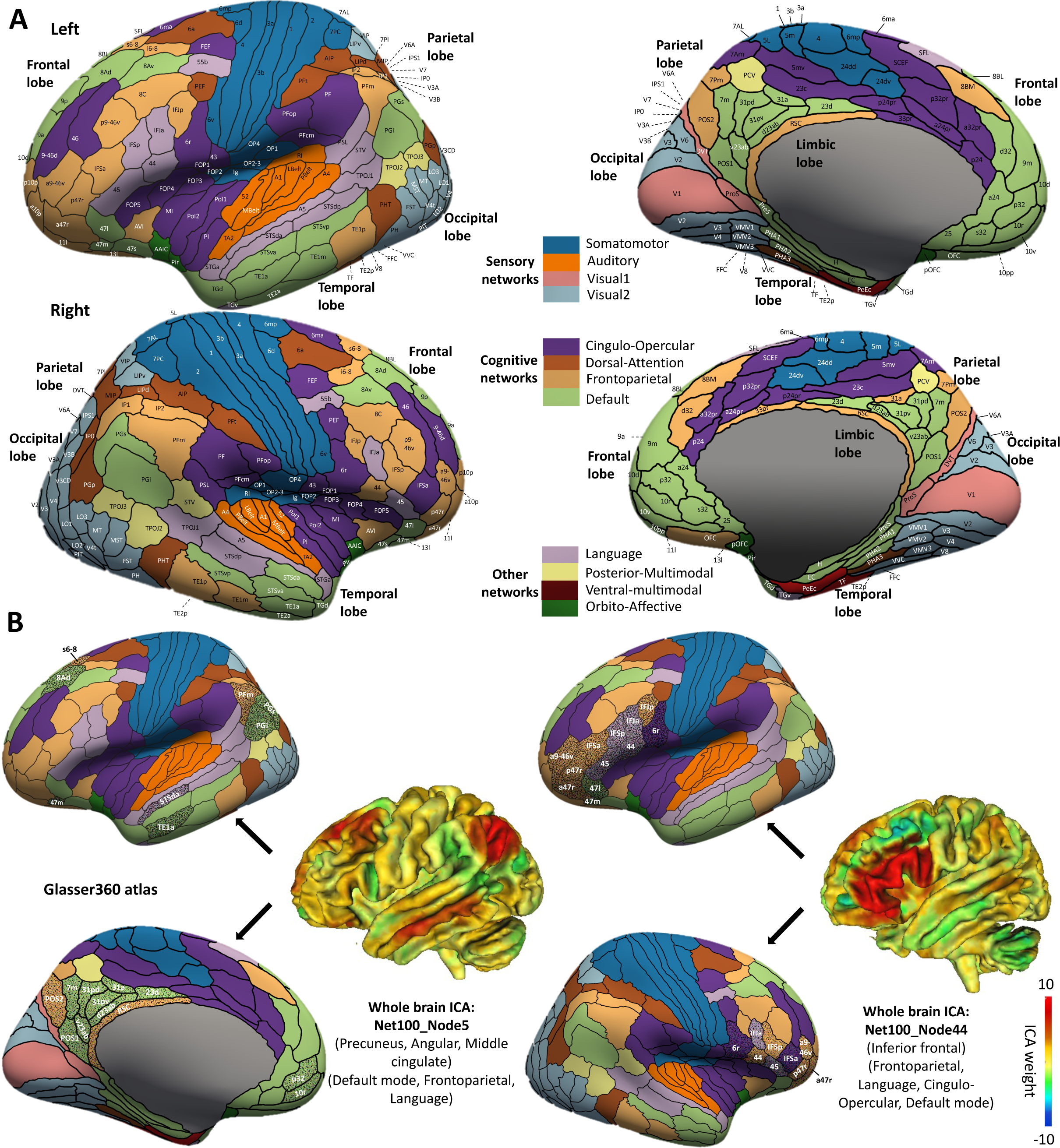
Illustration of the parcellation-based brain areas in the Glasser360 atlas and the comparison with selected whole brain ICA-defined brain regions. **(A)** We illustrate the 360 functional areas and 12 networks defined in the Glasser360 atlas. We generated functional connectivity traits based on these brain areas in both resting and task fMRI to investigate genetic influences on brain function. Table S1 provides information on these Glasser360 areas. Visual1, the primary visual network; Visual2, the secondary visual network. **(B)** We illustrate the spatial overlaps between the Glasser360 atlas and two selected whole brain ICA-defined brain regions (Net100_Node5 and Net100_Node44 in the left and right panels, respectively). The two ICA regions were previously generated from a 100-dimensions ICA analysis^30^ and estimated to have a high level of heritability^32^. Here we show that these two ICA regions distribute across multiple brain areas and networks in the Glasser360 atlas, as highlighted by dots with names being labeled. Specifically, the Net100_Node5 region (mainly in the precuneus, angular, middle cingulate) overlapped with 20 areas in the default mode, frontoparietal, and language networks, and the Net100_Node44 region (mainly in the inferior frontal) overlapped with 21 areas in the frontoparietal, language, cingulo-opercular, and default mode networks. A summary of such overlaps and additional examples of ICA regions can be found in Figure S2.

## RESULTS

### Heritability of human cerebral cortex functional connectivity at rest and during a task

We examined the heritability pattern of functional connectivity traits across different functional areas and networks. Using the UKB individuals of white British ancestry (*n* = 34,641 for resting and 32,144 for task), SNP heritability was estimated via GCTA^41^ for the 8,531 area-level within-network functional connectivity traits in both resting and task fMRI. The mean heritability (*h*^2^) was 10.4% for resting and 6.6% for task fMRI. Overall, the SNP heritability of 97.9% (8,349/8,531, *h*^2^ range = (2.4%, 27.0%)) functional connectivity traits in resting fMRI and 80.8% (6,894/8,531*, h*^2^ range = (2.9%, 24.1%)) functional connectivity traits in task fMRI remained significant after adjusting for multiple comparisons using the Benjamini-Hochberg procedure to control the false discovery rate (FDR) at 0.05 level (**Table S2**). We also estimated the heritability of the 1,985 network-level traits (1,066 for resting and 919 for task), 1049 resting fMRI traits and 893 task fMRI traits were significant at the FDR 0.05 level. The mean *h*^2^ was 32.2% for amplitude traits (*h*^2^ range = (20.8%, 46.1%)) and 12.2% for functional connectivity traits in resting fMRI (*h*^2^ range = (3.3%, 32.7%)); and that was 19.6% for amplitude traits (*h*^2^ range = (11.3%, 23.7%)) and 10.6% for functional connectivity traits (*h*^2^ range = (3.4%, 24.9%)) in task fMRI (**Table S3**). To better visualize the observed patterns, we mapped the area-level and network-level traits back to the 360 cortical areas and created the surface maps of the average heritability. Consistent with the above observations, we found that amplitude traits were more heritable than functional connectivity traits (*P* < 2.2 × 10^-16^, Wilcoxon rank test). Furthermore, among functional connectivity traits, network-level traits exhibited higher heritability than area-level traits (*P* < 2.2 × 10^-16^, as shown in **Figs. S3** and **S4A**). The higher heritability of network-level fMRI traits may suggest that our dimension reduction approach reduces noise by aggregating fMRI signals or that genetics have a stronger influence on broader brain networks rather than specific area pairs. In addition, the average heritability of Glasser360 parcellation-based traits was higher than that of the whole brain ICA-based traits^31–33^ estimated from almost the same UKB resting fMRI dataset (mean = 9.4% and 27.2% for functional connectivity and amplitude traits, respectively, *P* < 2.2 × 10^-16^). The results were summarized in **Figures S4B-S4E**.

Our parcellation-based traits revealed area- and network-specific information about genetic influences on the human brain. **Figures 2A** and **S5A** illustrate the heritability pattern across different networks. The mean heritability was highest in the ventral multimodal network in resting fMRI (mean = 20.4%). The ventral multimodal is a recently identified network, consisting of four cortical areas (left/right TF and PeEc) on the ventral surface of the temporal lobe^39^. One possible function of this network is to represent higher-order semantic categories^39^. In addition, the mean heritability of task fMRI was lower than that of resting fMRI in all networks except for the secondary visual network, where more than half (52.32%) of connectivity traits had greater heritability in task fMRI than in resting fMRI (**Fig. S5B**). This might be partly because the visual cortex is highly activated when performing the emotional processing task^36,37^. We also compared the estimates of genetic variance between the resting and task fMRI. We observed a general reduction in genetic variance in task fMRI (*P* < 2.2 × 10^-16^), and the spatial patterns of genetic variance were similar to those of heritability (**Figs. S6-S7**). The correlation between all heritability estimates for resting and task fMRI traits was 0.223, with a range of 0.024 to 0.386 in most networks (**Fig. S8**). Overall, large-scale fMRI data indicate different genetic influences in resting and task functional connectivity measurements.

**Fig. 2.**
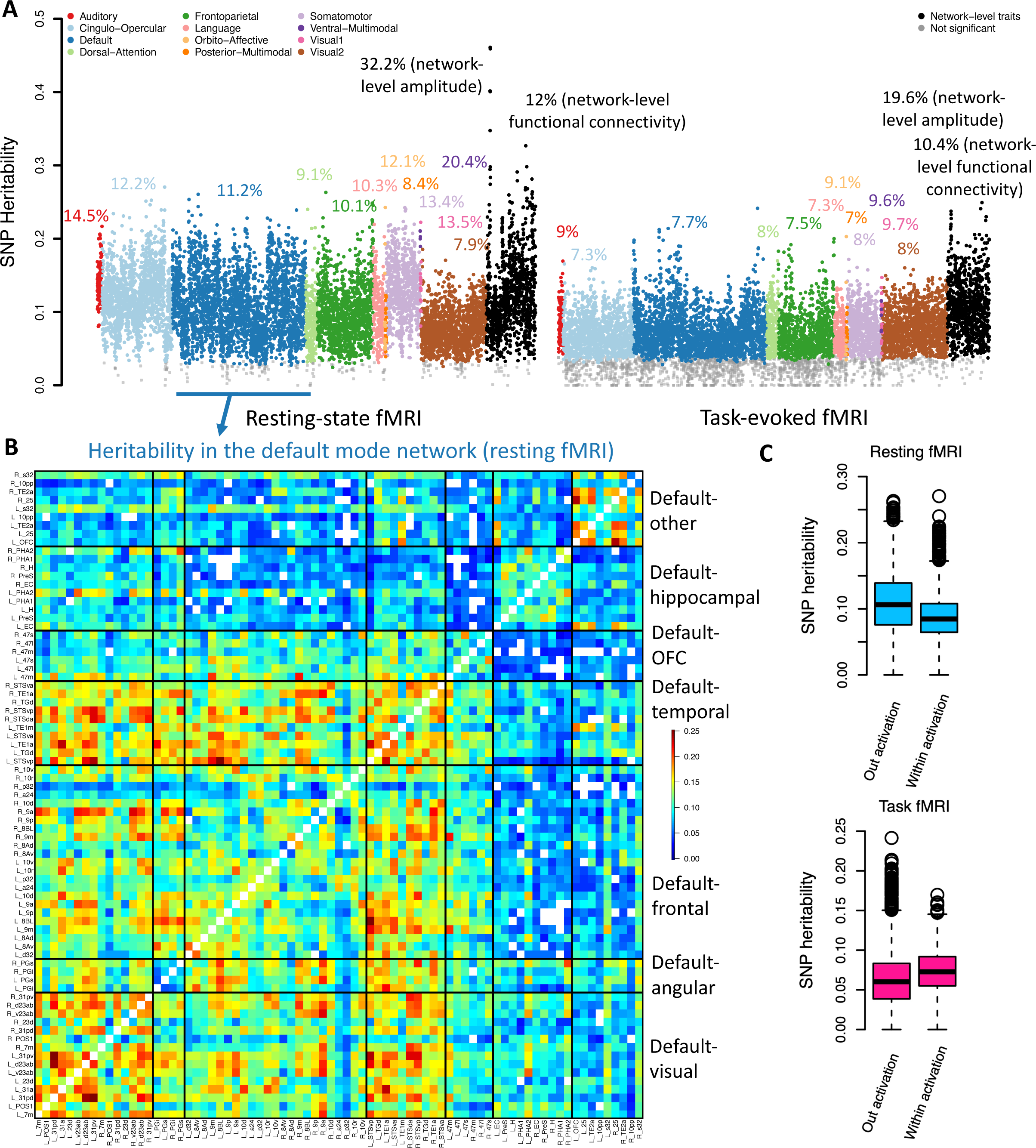
SNP heritability pattern in resting and task fMRI. **(A)** The dots represent the SNP heritability estimates of fMRI traits, including 8,531 area-level and 1,066 network-level traits in resting fMRI (left panel), and 8,531 area-level traits and 919 network-level traits in task fMRI (right panel). The dots with gray color represent non-significant heritability estimates (after controlling for multiple testing at a false discovery rate of 5%). The mean heritability of each group is labeled, for example, the mean heritability of traits in the auditory network was 14.5% in resting fMRI. **(B)** Significant SNP heritability estimates of the functional connectivity traits within the default mode network in resting fMRI (white dots are insignificant estimates). We grouped all brain areas of the default mode network into seven clusters, which were mainly organized by their physical locations (Fig. S7). OFC, orbitofrontal complex. The *x* and *y* axis represent the names of the brain areas. **(C)** Comparison of SNP heritability between the activated areas (within the activation, defined by the Shapes activation in task fMRI) and the nonactivated areas (out activation) in resting fMRI (upper panel) and task fMRI (lower panel). More information about the Shapes activation is available in Alfaro-Almagro, et al. ^30^.

In each network, the heritability pattern across functional areas was identified. For example, according to physical locations, the areas of the default mode network can be divided into seven clusters (**Fig. S9**). There is no significant correlation between the physical distance of these default mode area pairs and their heritability measurements (*P* = 0.563). In the visual cluster (including the precuneus, calcarine, and cingulate) and the temporal cluster, the default mode connectivity traits were most heritable. Additionally, the connectivity traits between the two clusters and the angular and frontal clusters showed high heritability, indicating the high degree of genetic control of functional interaction among these physically disconnected regions (**Fig. 2B**). In the somatomotor network, the left/right 3a, 3b, and 4 areas (in the postcentral, precentral, and paracentral) and the left/right 7AL and 7PC areas (in the superior parietal) form two separate connectivity clusters. We found that the 3a, 3b, and 4 areas had the highest heritability within the somatomotor network, while the heritability of the 7AL and 7PC areas was low. In addition, the connectivity traits associated with the left OP2-3 area (in the Rolandic operculum and insula) had high heritability (**Fig. S10**). In the cingulo-opercular network, the insula-related areas (e.g., the left/right FOP5, PoI1, FOP3, FOP1, FOP4, MI, and Pol2) exhibited the highest heritability. However, the heritability of the adjacent para-insular area (the left/right Pl, in the temporal pole and superior temporal) was low (**Fig. S11**). The insula is a functionally diverse part of the cortex involved in multiple functions, including emotion, cognition, and sensory perception^42,43^. The insula has been found to have the highest heritability in surface curvature analysis of cortical morphometry^22^. Additionally, the connectivity traits of several specific areas showed consistently high heritability in resting fMRI, including the left/right IPS1 areas (in the superior occipital) of the secondary visual network and the right TE1m and left/right TE1p areas (in the inferior temporal and middle temporal) of the frontoparietal network (**Fig. S12**). In task fMRI, we also observed a few areas that had higher heritability than others, including the visual cluster, left OFC (orbitofrontal complex), and left/right 25 areas (in the olfactory cortex) of the default mode network (**Fig. S13**); the left/right RSC (in the middle cingulate), POS2 (in the precuneus and cuneus), 7Pm (in the precuneus) areas of the frontoparietal network (**Fig. S14**); and the middle cingulate-related areas (e.g., the left/right p24pr, a24pr, and left 33pr) of the cingulo-opercular network (**Fig. S15**). Furthermore, we investigated the relationship between heritability estimates and the size of cortical areas. We found that a greater signal-to-noise ratio in larger areas may account for some of the observed differences in amplitude heritability (R-squared range = [0.03, 0.07], *P* < 7.47 × 10^-4^), but has a weaker effect on functional connectivity heritability (R-squared range = [0, 0.02], *P* > 5.14 × 10^-3^, **Fig. S16**). In summary, our results demonstrate the diverse genetic influences on the cerebral cortex and highlight the crucial functional areas and their interactions that are significantly impacted by genetic factors.

The heritability pattern was also correlated with the activation maps defined in task fMRI (Shapes activation, Faces activation, and Faces-Shapes contrast)^30^. In resting fMRI, the heritability of functional connectivity traits located in task-defined activated regions was lower than that of nonactivated functional connectivity traits (**Fig. 2C**, mean = 8.9% vs. 10.8%, *P* < 2.2 × 10^-16^). However, in task fMRI, activated traits had higher heritability than nonactivated ones (mean = 7.4% vs. 6.3%, *P* < 2.2 × 10^-16^). Similar results were also observed on genetic variance estimates (*P* < 2.2 × 10^-16^, **Fig. S6B**). These differences can be partially explained by the task-related changes in brain functional activities and inter-regional connectivities^44^. Our findings may also be related to previous observations that the correlation between two activated regions increases during task performance, whereas the correlation between other regions is decreased^45^. Overall, these results provide insights into the genetic influences on intrinsic and extrinsic functional architectures.

### Genetic loci associated with cerebral cortex functional areas and networks

GWAS was performed for the 8,531 × 2 within-network area-level connectivity traits in resting and task fMRI using the UKB individuals of white British ancestry (Methods). The LDSC intercepts^46^ were close to one, suggesting no genomic inflation of test statistics due to confounding factors (mean intercept = 1.0002, range = (0.987, 1.019)). At a stringent significance level 2.93 × 10^-12^ (5 × 10^-8^/8,531/2, additionally adjusted for the number of traits studied), we identified independent (linkage disequilibrium [LD] *r*^2^ < 0.1) significant associations in 32 genomic regions (cytogenetic bands) for resting fMRI, 9 of which were also associated with task fMRI (**Fig. 3A** and **Table S4**).

**Fig. 3.**
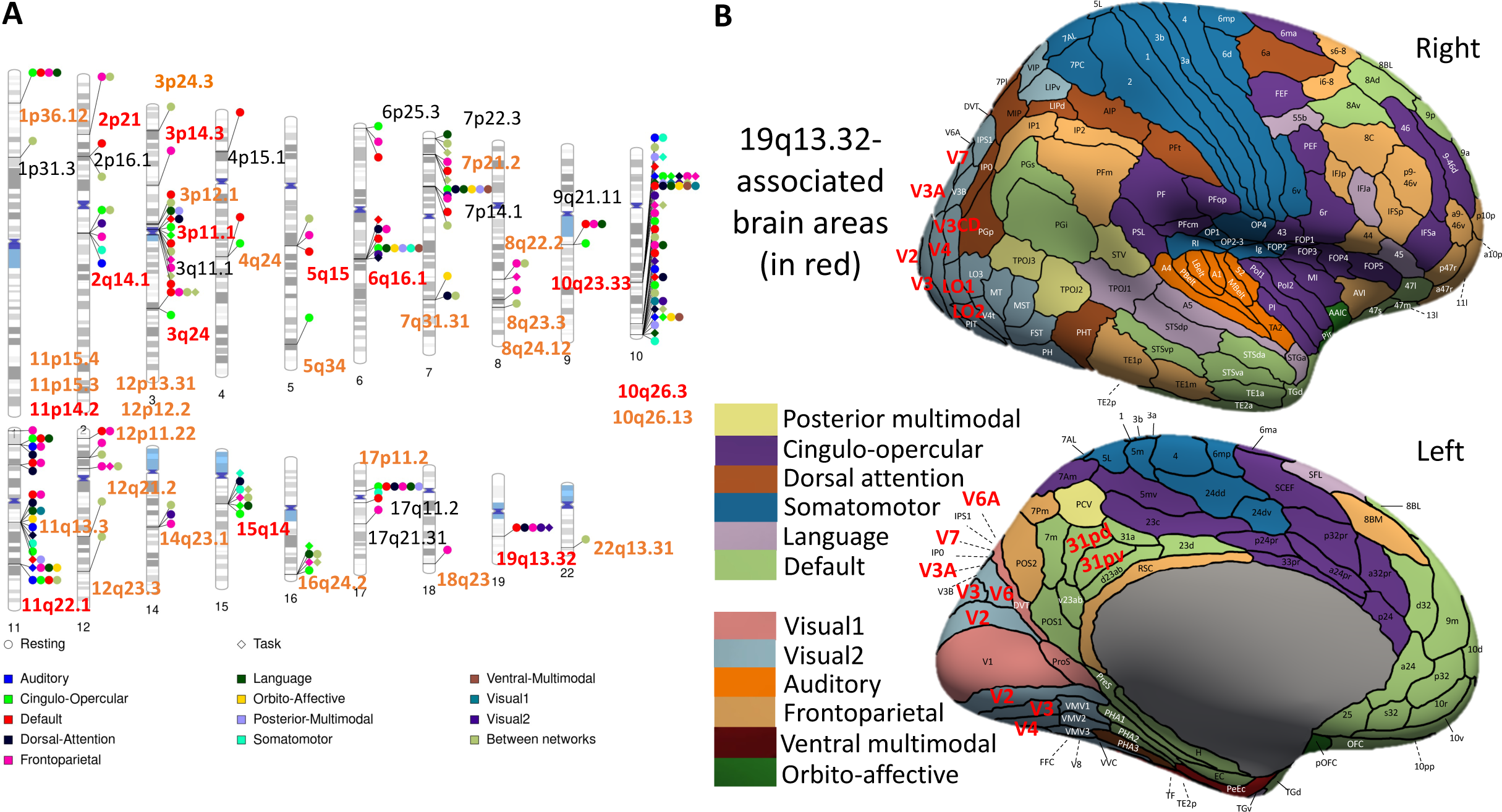
The associated genomic regions of fMRI traits. **(A)** Ideogram of 47 genomic regions influencing fMRI traits, with 32 regions identified by area-level traits (*P* < 2.93 × 10^-12^) and 15 more identified by network-level traits (*P* < 2.51 × 10^-11^). Each dot represents a genomic region, and the colors correspond to the 12 networks (and their interactions, named Between networks). A signal dot indicates that at least one fMRI trait (either area-or network-level) of the corresponding network is associated with the genomic region. Red and brown labels indicate genomic regions that were replicated at the Bonferroni significance level and nominal significance level, respectively. **(B)** The 19q13.32 genomic region was found to be associated with several functional areas (*P* < 2.93 × 10^-12^), most of which were in the secondary visual network. The colors of the brain areas represent different networks, and significant brain areas are highlighted with red labels.

Enrichment of locus-specific genetic effects was observed across different areas and networks in resting fMRI. Many of the fMRI-associated genetic variants are known to affect gene expressions in previously published human brain expression quantitative trait loci (eQTL) datasets^47,48^. We have highlighted the associated brain areas and networks for identified genomic loci in **Figures S17-S45** and summarized these results in **Table S4**. For example, most of the associations of the 2q14.1 locus were in the somatomotor network. The 2q14.1 locus was also particularly associated with the right MST and right V6 areas (in the middle temporal and cuneus) of the secondary visual network and a few areas (e.g., the left/right 43 in the Rolandic operculum and the p24pr in the middle cingulate) of the cingulo-opercular network (**Fig. S17**). **Figure S18** illustrates the genes presented at this locus and shows that the index variant (and its proxy variants, LD *r*^2^ > 0.8) are associated with the expression of *PAX8* and *FOXD4L1* in brain tissues^47^, suggesting that the two genes are relevant to neuronal function. Interestingly, all associations at the 19q13.32 locus, which was the major genetic risk factor of the late-onset Alzheimer’s disease, were in the secondary visual network (such as the left LO1 and right V3CD in middle occipital, left/right V3A in superior occipital, and left/right V6 in cuneus), with one exception in the visual cluster of the default mode network (**Figs. 3B** and **S29-S30**). These results highlight the close relationship between the 19q13.32 locus and the functional connectivity measurements of the visual cortex. For task fMRI, the 10q23.33 locus was mainly associated with the visual cluster of the default mode network (**Figs. S43-S44**), especially the left/right 31pv areas (in the middle cingulate) (**Fig. S45**). In other genomic loci (e.g., 10q26.3, 3p11.1, and 19q13.32), the associated networks in task fMRI were similar to those in resting fMRI, although the number of connectivity traits surviving the stringent significance level became much smaller. We examined the pairwise genetic correlations between the 8,531 connectivity traits in resting and task fMRI via the cross-trait LD score regression^49^ (Methods). The average genetic correlation among all the 8,531 pairs was 0.554, 3,598 of which were significant at the FDR 5% level (**Fig. S46**, mean = 0.710, standard error = 0.192). Although it was more difficult to identify associated loci for task fMRI, these strong genetic correlations suggest the overall similarity of the genetic architecture on brain functions at rest and during a task.

Next, we performed GWAS for the 1,985 network-level traits to identify variants associated with network-specific functional connectivity measurements (within each of the 12 networks and between each pair of networks). At the 2.51 × 10^-11^ (5 × 10^-8^/1,985) significance level, we identified 41 (15 additional) genomic regions for resting fMRI, 14 of which were also associated with task fMRI (**Table S5**). On average, these 41 genetic regions explained 13.5% of the heritability of network-level fMRI traits. Together, the area- and network-level analysis identified 47 genomic regions for resting fMRI, 15 of which were also associated with task fMRI (**Fig. 3A**). The whole brain ICA analysis on largely the same resting fMRI dataset identified 45 associated genomic regions^32,33^ (*P* < 2.81 × 10^-11^, 5 × 10^-8^/1,777). Of the 45 regions, 31 were also identified in our study, with more detailed association patterns being uncovered (**Table S6** and **Fig. S47**). For example, the 19q13.32 region (index variant rs429358) had been found to be associated with the amplitude traits of large ICA-defined regions spanning multiple cognitive and visual networks. In the present study, as shown in **Figures 3B** and **S29-S30,** the area- and network-specific fMRI traits prioritized associations with the visual cortex. Furthermore, we identified 16 new loci that were associated with multiple areas and networks. For example, the 12p11.22 region (index variant rs11049367) was associated with the functional connectivity traits of the frontoparietal and default mode networks, especially the connectivity between the right p9-46v and right TE1m areas in the frontoparietal network (*P* < 2.12 × 10^-11^). Of the 47 fMRI-associated loci, 19 had been linked to brain structural connectivity traits in a recent study of white matter microstructure using diffusion MRI (dMRI)^50^ (**Table S7**). A few of the 19 overlapped loci had wide genetic effects on multiple white matter tracts and functional networks, such as the 16q24.2, 3p11.1, 16q24.2, and 15q14 (**Fig. S48**).

We aimed to replicate the identified genomic loci using independent European and non-European datasets. First, we repeated GWAS on a European dataset with 8,197 subjects, including European individuals in the UKB phases 4 and 5 data (up to 2023 release, removed the relatives of our discovery sample) and individuals of white but non-British ancestry in UKB phases 1 to 3 data. For the 266 independent (LD *r*^2^ < 0.1) network-locus associations in resting fMRI, 93 (34.9%) passed the Bonferroni significance level (1.9 × 10^-4^, 0.05/266) in this validation GWAS, and 200 (75.1%) were significant at the nominal significance level (0.05). All of the 200 significant associations had concordant directions in the two GWAS (**Fig. S49A**). Of the 47 identified genomic loci, at least one association of 21 loci (44.6%) passed the Bonferroni significance level and 39 (82.9%) can be validated at the nominal significance level. For task fMRI, 29.8% (i.e., 17/57) network-locus associations passed the nominal significance level, all of which had the same effect signs in the two GWAS (**Fig. S49B**). The 17 associations were related to eight genomic loci, four of which were significant at the Bonferroni significance level, including the 10q23.33, 16q24.2, 10q26.3, and 15q14.

Next, we performed GWAS on two UKB non-European validation datasets: the UKB Asian (UKBA, *n* = 517) and UKB Black (UKBBL, *n* = 283). Of the 47 genomic loci, 15 (2 also in task) were validated in UKBA and 12 (5 also in task) were significant in UKBBL at the nominal significance level, one of them (2q13 in UKBA) survived the Bonferroni significance level (**Tables S4-S5**). For resting fMRI, we performed further analysis using data from the ABCD study. For the 266 independent (LD *r*^2^ < 0.1) network-locus associations in resting fMRI, 40 (15.0%) were significant at the nominal significance level, most (37/40) had concordant directions in UKB and ABCD European cohort (*n* = 3,821) (**Fig. S49C**). Of the 47 identified genomic loci, at least one association of 16 loci (34.0%) can be validated at the nominal significance level. In addition, 18 associations in 8 loci and 15 associations in 9 loci can be validated in ABCD Hispanic (*n* = 768) and African American (*n* = 1,257) cohorts, respectively.

Several additional analyses were performed to evaluate the robustness of our GWAS results. First, we used the Yeo-7 atlas^51^ to group the 360 cortical areas into 7 functional networks and derived 615 network-level traits (294 for resting fMRI and 321 for task fMRI) using the same dimension reduction procedure as we used for the 12 functional networks^39^. The heritability estimates for these Yeo-7 network-level traits were in a similar range to those based on the 12 networks (**Fig. S50**). At the stringent GWAS significance level 8.13 × 10^-11^ (5 × 10^-8^/615, additionally adjusted for the number of Yeo-7 traits), we identified 28 genomic regions for resting fMRI, 23 of which overlapped with the 41 regions identified by the 12 networks, and 22 overlapped with the 45 regions identified by whole brain ICA analysis^32,33^ (**Fig. S51**). In task fMRI, four regions were identified and all of them were also identified by the network-level traits in our main analyses (**Table S8**). Second, we applied additional quality control procedures for fMRI data and removed more images with potentially poor quality (492 subjects in resting fMRI and 993 in the task fMRI). We rerun our heritability and GWAS analyses and found that the results were consistent before and after these additional quality control steps (**Figs. S52-S53**). Finally, we used a split-half design for our discovery GWAS sample to further examine the replicability of our results. The significant genetic estimates produced by our discovery GWAS were similar in the two independent half-samples for both area-level and network-level traits (**Fig. S54**). Additionally, we used the first half of the sample as a discovery dataset, and the second half as a replication dataset with balanced sample size and homogeneous genetic background. Using the same significance level as for the full discovery GWAS, we identified 17 genomic regions associated with resting fMRI, three of which were associated with task fMRI. All these regions were among the 47 genomic regions reported in **Figure 3A**. The genetic estimates in the two half-samples were highly similar (correlation = 0.981, **Fig. S55**).

### The shared genetic influences with complex brain traits and disorders

To evaluate the shared genetic influences between brain functional organizations and complex brain traits and diseases, we conducted association lookups for independent (LD *r*^2^ < 0.1) significant variants (and variants in LD, *r*^2^ ≥ 0.6) detected in the UKB white British GWAS. In the NHGRI-EBI GWAS catalog^52^, our results tagged variants reported for a wide range of complex traits and diseases, such as neurological disorders, neuropsychiatric disorders, mental health and psychological traits, migraine, cognitive traits, educational attainment, sleep, smoking/drinking, and anthropometric measurements. The top five frequently tagged regions included 19q13.32 (the most frequently reported gene in prior literature was *APOE*), 2q14.1 (*PAX8*), 10q23.33 (*PLCE1*), 17q21.31 (*MAPT*), and 6q16.1 (*FHL5*), which covered 81.66% of all tagged trait-variant associations. To explore the detailed genetic overlap pattern across brain functional areas, we further took the index variants of these complex traits/diseases and performed variant-specific association analysis for all the 64,620 functional connectivity traits (Methods). All the loci highlighted below have been replicated at the nominal significance level in the validation GWAS. These results have been summarized in **Table S9.**

The rs429358 (19q13.32), one of the two variants in the *APOE* ε4 locus, is a well-known risk factor for Alzheimer’s disease. In this locus, we observed the shared genetic influences with Alzheimer’s disease in both resting and task fMRI (**Figs. 4A-B**). We further applied Bayesian colocalization analysis^53^ to test colocalization between fMRI traits and Alzheimer’s disease, which was defined as having a posterior probability of the shared causal variant hypothesis (PPH4) > 0.8^53,54^. Strong colocalizations between fMRI traits and Alzheimer’s disease were identified (PPH4 > 0.946). Furthermore, in both resting and task fMRI, the secondary visual network had the strongest association with rs429358 (**Figs. 4C** and **S56A**). The allele associated with increased risk for Alzheimer’s disease (“C”) was associated with decreased functional connectivity in the visual cortex (**Fig. S57**). Visual deficits were one of the first symptoms of Alzheimer’s disease^55^ and functional connectivity deficits in the visual cortex have been reported in Alzheimer’s disease^56,57^. Decreased eigenvector centrality in the visual cortex was associated with *APOE* ε4 carriership among normal elderly subjects^58^. In addition, the risk allele at rs429358 was associated with decreased default mode activity at the Bonferroni significance level in variant-specific analysis (3.86 × 10^-7^, 0.05/64,620/2), with distinct patterns in resting and task fMRI (**Fig. S56B**). In resting fMRI, the associations were mainly in the visual cluster and the left/right 10d areas (in the superior frontal). Decreased default mode network connectivity in the posterior cingulate cortex/precuneus, orbital and middle frontal cortex, and inferior parietal lobe in *APOE* ε4 carriers has been consistently reported in various studies across adulthood (see Section 3.3.1 and Table 2 of Foo, et al. ^24^). Regardless of *APOE* carrier status, results from previous studies are consistent in finding decreased functional connectivity in the default mode network in subjects with Alzheimer’s disease and mild cognitive impairment, as summarized in Dennis and Thompson ^59^ and Badhwar, et al. ^8^. For task fMRI, most of the significant rs429358 effects were on the interactions between the visual cluster and a few areas in the frontal cluster, including the left/right p32 (in the medial superior frontal), a24 (in the pregenual anterior cingulate cortex), and 8Ad (in the superior frontal) areas. The reduced deactivation of the default mode network during tasks has been observed in different stages of Alzheimer’s disease^60^ and normal carriers of the *APOE* ε4^61^. Biologically, amyloid-β (Aβ) accumulation preferentially starts in several of the core regions of the default mode network, and the earliest Aβ accumulation is further associated with hypoconnectivity within the default mode network and between the default mode and frontoparietal networks^62^. The rs429358 was also strongly associated with the secondary visual and default mode networks in our analysis of the Yeo-7 networks (**Fig. S58**). Furthermore, we found the risk allele at rs429358 decreased the functional connectivity of middle temporal areas in the language network (e.g., the left/right TPOJ and STSdp) and the left IP0 area (in the middle occipital) of the dorsal attention network in task fMRI, but not in resting fMRI (**Fig. S59**). We also tested the association between rs429358 and MRI traits of other imaging modalities, including structural connectivity traits from dMRI^50^ and regional brain volumes from structural MRI (sMRI)^63^. The fMRI traits had much stronger associations (smaller *P*-values) with rs429358 than these structural traits. Together, these results suggest the fMRI traits of brain functions, especially the ones from the visual cortex in the secondary visual and default mode networks, might be more directly related to the genetic pathways of *APOE* ε4 to Alzheimer’s disease than brain morphology. These fMRI traits could be used as imaging biomarkers in etiologic study of Alzheimer’s disease and drug development targeting *APOE* ε4.

**Fig. 4.**
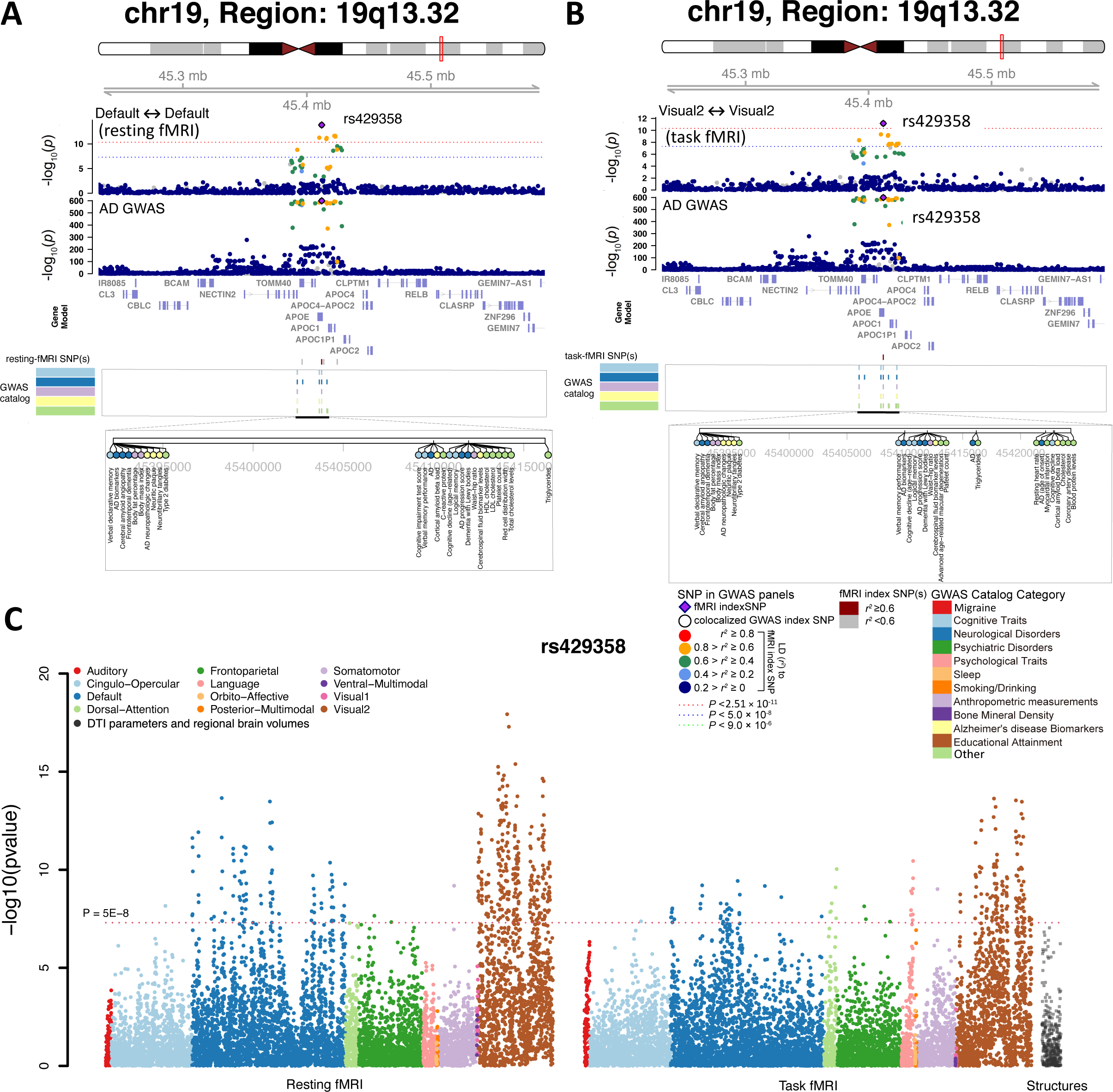
Genetic locus associated with both fMRI traits and Alzheimer’s disease. In the 19q13.32 region, the *APOE* ε4 locus (index variant rs429358) showed associations with both Alzheimer’s disease and functional connectivity measurements. In **(A)**, we illustrate the shared genetic influences for one functional connectivity trait of the default mode network (Default <-> Default) in resting fMRI. In **(B)**, we illustrate the shared genetic influences for one functional connectivity trait of the secondary visual network (Visual 2 <-> Visual 2) in task fMRI. In **(C)**, we illustrate the *P*-value of associations between the rs429358 and various neuroimaging traits, including functional connectivity traits in resting fMRI, functional connectivity traits in task fMRI, as well as structural traits (in gray color), including diffusion tensor imaging (DTI) traits from diffusion MRI and regional brain volumes from structural MRI. The strongest genetic effects were observed in the secondary visual network for both resting and task fMRI.

In the 17p11.2 and 2p16.1 regions, we observed the shared genetic influences between the default mode network and multiple psychiatric disorders, including schizophrenia^64,65^, autism spectrum disorder^66^, MDD^67^, and epilepsy^68^. For example, we tagged rs4273100 and rs1518395, which have been implicated with schizophrenia (**Fig. S60**). Rs1518395 was also a risk variant for MDD and the rs2947349 in 2p16.1 was associated with epilepsy. In resting fMRI, all three index variants exhibited the strongest associations with the default mode network, especially the frontal cluster (**Figs. S61-S62**). In addition, we observed the shared genetic influences with cognitive ability and education^69^ in the 17p11.2 region and with psychological traits (e.g., neuroticism^70^ and subjective well-being^71^) in the 2p16.1 region, which were also mainly related to the default mode network (**Fig. S63**). In the 10q23.33 and 6q16.1 regions, our identified variants tagged those that have been implicated with migraine^72,73^ (**Fig. S64**). These genetic effects affected multiple networks and had the strongest associations in the auditory and cingulo-opercular networks (**Figs. S65**-**S66**). We also tagged risk variants of brain aneurysm^74^ and cerebral blood flow^75^ in these regions. Migraine is a heterogeneous disorder and no neuroimaging biomarker has been well established in previous small sample fMRI studies^76^. Our results may help identify whether risk variants of migraine predispose to migraine in particular brain areas or networks. Specifically, our findings suggest the genetic overlaps among migraine, cerebrovascular traits, and brain functions across multiple networks, especially the auditory and cingulo-opercular networks.

The shared genetic influences with cognitive ability^69^, intelligence^77^, and education^69^ were observed in the 10q26.13, 5q15, and 3p11.1 regions (**Fig. S67**). The index variants of these cognitive traits were associated with a few specific functional areas in the temporal and parietal lobes. For example, in the 10q26.13 locus, our tagged variant (rs2629540) was associated with math ability, education^69^, and cocaine dependence^78^. Rs2629540 was particularly associated with the precuneus-related areas in different networks, such as the right PCV in the posterior multimodal network, the left/right 7Pm in the frontoparietal network, and the right 31pd in the default mode network (**Fig. S68**). The precuneus is involved in a variety of complex functions and responds to a wide variety of cognitive processes^79^. In the 5q15 region, we observed shared genetic effects with cognitive performance, math ability, and education^69^, most of which were related to the left/right TPOJ2 areas (in the middle temporal) of the posterior multimodal network (**Fig. S69A**). The *NR2F1* is well studied in the arealization of the cerebral cortex^80^. In the 3p11.1 region, we tagged variants associated with intelligence^77^ (rs7652296, *EPHA3*). The *EPHA3* is involved in axon guidance^81^ and the rs7652296 was mainly associated with between-network connectivity of a few temporal and parietal areas (**Fig. S69B**). These findings partially support the parieto-frontal integration theory of intelligence^82,83^, uncovering the genetic overlaps between cognitive functions and specific temporal and parietal functional areas.

In addition, we found colocalized genetic effects with psychological traits (e.g., risk-taking behaviors^84^) in the 3q24 (rs2279829) and 3p12.1 (rs6762267) regions (**Fig. S70**). Rs2279829 and rs6762267 were mainly related to the interactions among the frontoparietal, cingulo-opercular, and default mode networks, with the strongest genetic effects being on a few frontal areas (**Fig. S71**). Increasing evidence suggests the frontal lobe plays an important role in risk-taking and risk behaviors^85^. In summary, brain functions measured in fMRI have substantial area-specific genetic overlaps with complex brain traits and clinical outcomes. Uncovering the detailed genetic colocalized patterns may help understand how alterations in specific brain functions lead to risk for brain conditions and disorders.

### Genetic correlations with complex traits

To further explore the genetic links, we examined the genetic correlations (GC)^49^ between fMRI traits and 50 complex traits, most of which had genetic overplays with fMRI traits, as well as additional mental health traits and major brain disorders. First, we examined the genetic correlations with 4 global functional connectivity and amplitude traits (2 traits for resting and 2 for task). At the FDR 5% level (4 × 50 tests), we found the global fMRI traits were significantly associated with hypertension, neuroticism (e.g., feeling nervous and worry), sleep traits, and task-taking behaviors (e.g., automobile speeding) (**Table S10**). For example, resting functional connectivity was negatively correlated with neuroticism (feeling nervous, GC = −0.181, *P* < 1.14 × 10^-4^) and sleep duration (GC = −0.173, *P* < 1.58 × 10^-4^). Hypertension was negatively correlated with the global amplitude in task fMRI (GC = −0.282, *P* < 7.34 × 10^-6^).

Next, we explored the spatial patterns of genetic overlaps by evaluating the genetic correlations between complex traits and 8,531 functional connectivity traits. Area- and network-specific genetic overlaps were widely observed in resting fMRI. For example, at the FDR 5% level (8,531 tests), cognitive function^86^ had genetic correlations with cognitive networks (the cingulo-opercular, default mode, frontoparietal, and dorsal attention), such as the right IFSa area (in the triangular part of inferior frontal) (**Fig. 5A**). Most of the significant genetic correlations were negative, which suggest genetic effects predispose to less resting connectivity is associated with increased intelligence. For schizophrenia^87^ and cross-disorder^88^, we found consistent positive genetic correlations with the default mode network (e.g., the left/right 47s in the posterior orbital and the left/right 8BL in the medial superior frontal) and negative genetic correlations with the secondary visual network (e.g., the left/right LIPv in the superior parietal) (**Figs. 5B** and **S72**). Similarly, we found neuroticism^89^ (feeling nervous) had positive genetic correlations with the default mode network (e.g., the right PGi in the angular) and negative genetic correlations with the secondary visual network (e.g., the right FST in the middle temporal and the right VMV3 in the fusiform) (**Fig. S73**).

**Fig. 5.**
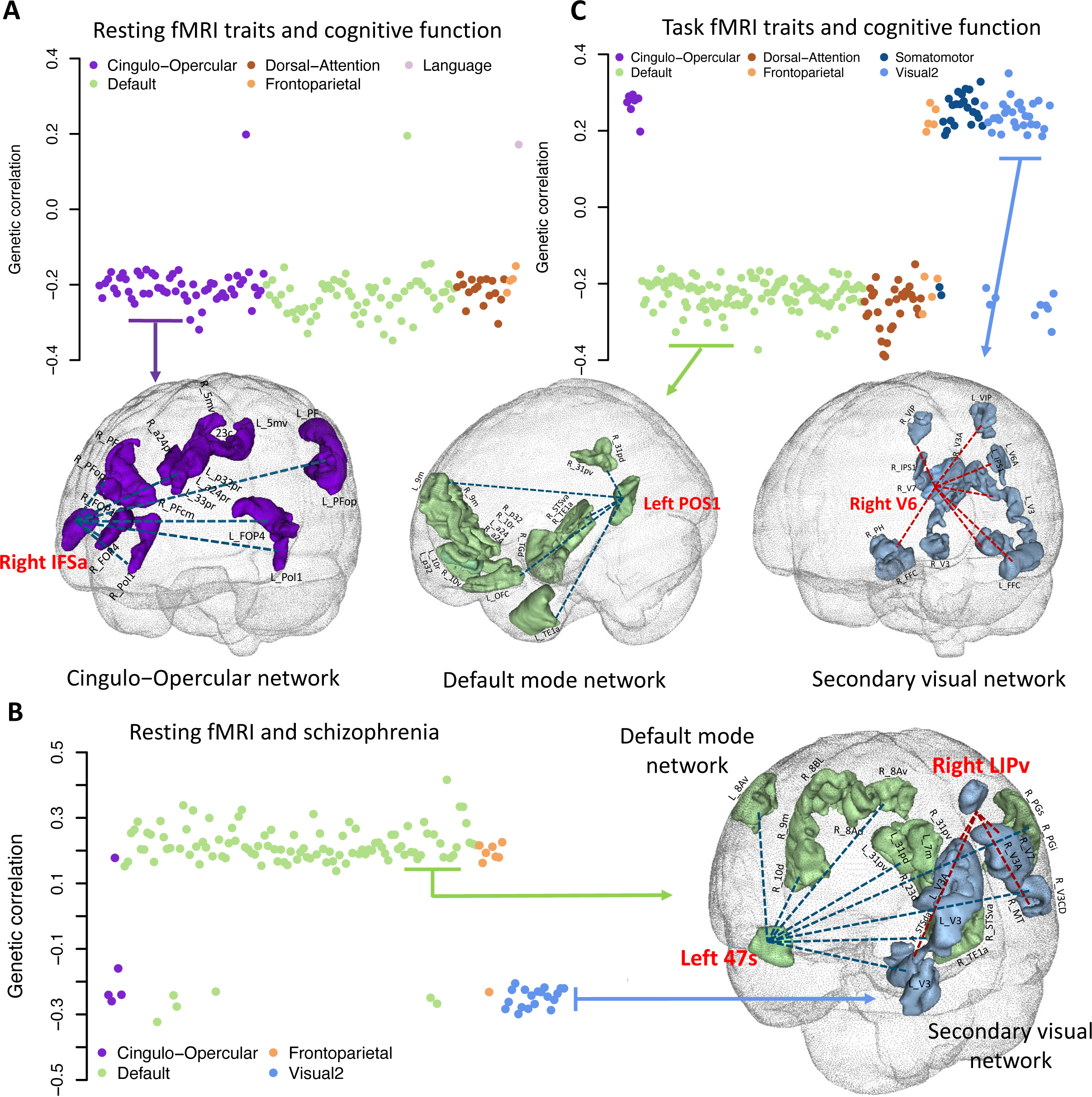
Selected pairwise genetic correlations between fMRI traits and cognitive function and schizophrenia. **(A)** We illustrate significant genetic correlations between cognitive function and functional connectivity across different networks in resting fMRI at the FDR 5% level. These significant genetic correlations were particularly related to functional connectivity traits of specific areas, including the right IFSa in the cingulo−opercular network. Most of the genetic correlations were negative. See Table S10 for the full list of estimates. **(B)** We illustrate significant genetic correlations between schizophrenia and functional connectivity across different networks in resting fMRI at the FDR 5% level. These significant genetic correlations were particularly related to the functional connectivity traits of specific areas, including the left 47s in the default mode network and the right LIPv in the secondary visual network. **(C)** We illustrate significant genetic correlations between cognitive function and functional connectivity across different networks in task fMRI at the FDR 5% level. These significant genetic correlations were particularly related to the functional connectivity traits of specific areas, including the left POS1 in the default mode network and the right V6 in the secondary visual network. Similar to the resting fMRI results in (A), genetic correlations of cognitive function with the default mode and dorsal attention networks were negative in task fMRI. However, the genetic correlations of cognitive function with the cingulo−opercular network became positive, and the somatomotor and secondary visual networks also had positive genetic correlations in task fMRI.

Task fMRI provided additional insights into the genetic correlations with cognitive function (**Fig. 5C**). Similar to resting fMRI, the default mode network had negative genetic correlations with cognitive function. The correlations were enriched in connectivity traits between the visual cluster and the frontal cluster (**Fig. S74A**). Moreover, the secondary visual and somatomotor networks (e.g., the right V6 in the cuneus, the left VMV2 in the lingual, the left VIP in the superior parietal, and the right OP2-3 in Rolandic operculum and insula) had positive genetic correlations with cognitive function (**Fig. S74B**). In summary, these results show the default mode network has negative genetic correlations with cognition and positive genetic correlations with brain disorders and neuroticism. The genetic correlations with cognition had opposite directions in the default mode network and the secondary visual network in task fMRI, pointing to genetic influences on task-specific brain activity. Patterns of other complex traits (e.g., snoring, sleep duration, hypertension, general risk tolerance, and education) were summarized in the **Supplementary Note** and **Figures S75-S76**. In summary, discovering genetic co-variations with complex traits and diseases in specific brain areas and networks might improve our understanding of how brain function is affected by genetic risk factors and aid early detection and timely treatment of brain diseases.

### Gene-level analysis and biological annotations

Using GWAS summary statistics of network-level fMRI traits, MAGMA^90^ detected 67 significant genes with 352 associations (*P* < 1.34 × 10^-9^, adjusted for 1,985 phenotypes). The area-level traits additionally yielded 7 associated genes for resting fMRI and 1 for task fMRI (*P* < 1.55 × 10^-10^, adjusted for 8,531 × 2 phenotypes, **Table 11**). Among the 75 genes, 30 had not been identified by the whole brain ICA analysis^32^ (**Fig. S77**). We performed functional lookups for these fMRI-associated genes. First, nine genes (such as *SSH2* and *KANSL1*) showed a high probability of being loss-of-function (LoF) intolerant^91^ (pLI > 0.98), indicating extreme intolerance of LoF variation. Second, 24 (such as *FAM53B* and *METTL10*) of the 67 genes were identified by a recent eQTL study of developing human brain^92^, and 15 were in previously constructed transcriptional networks^93^, such as the *FAT3, MEF2C, CRHR1, NR2F1,* and *VRK2,* which were in the *adult neurons, synaptic transmission, and neuron projection development* function module. Our findings showed substantial overlaps with genes reported in previous studies on brain genetics. For example, *CRHR1* plays an important molecular role in regulating amygdala function and anxiety^94–97^ and was significantly associated with the functional connectivity of the frontoparietal network (*P* = 2.34 × 10^-11^). The *MEF2C* was associated with frontoparietal, default mode, and language networks in our analyses (*P* < 2.25 × 10^-10^) and is known to cause multiple Mendelian disorders characterized by intellectual disability^98,99^. In addition, we applied FUMA^100^ to map significant variants (*P* < 2.51 × 10^-11^) to genes via physical position, eQTL association, and 3D chromatin (Hi-C) interaction. FUMA yielded 226 associated genes, 166 of which were not discovered in MAGMA (**Table S12**). For example, we found associations between the default mode network and *SLC6A4*, which is involved in the epigenetic mechanism of increased risk for mental illness and abnormal brain function^101–104^. In addition, 5 of the fMRI-associated genes (*CALY*, *SLC47A1, SLC6A4, CYP2C8,* and *CYP2C9*) were targets for 51 nervous system drugs^105^ (anatomical therapeutic chemical (ATC) codes starting with “N”), which was a higher probability of overlap than by chance (*P* = 0.027). Specifically, there were 29 anti-depressants (N06A) to treat MDD and related conditions, 13 anti-psychotics drugs (N05A) to manage psychosis, 4 psychostimulants (N06B) for attention-deficit/hyperactivity disorder (ADHD) and nootropics, and 2 anti-migraine (N02C) (**Table S13**).

To identify brain cell types where genetic variation leads to changes in brain function, we performed partitioned heritability analyses^106^ for cell type-specific regulatory elements. Specifically, we estimated partitioned heritability enrichment within differentially accessible chromatin of neurons (NeuN+, including two subtypes GABAergic [NeuN+/Sox6+] and glutamatergic neurons [NeuN+/Sox6-]) and glial cells (NeuN-, including oligodendrocyte [NeuN-/Sox10+], microglia and astrocyte [NeuN-/Sox10-])^107^. To identify global enrichment across the whole brain, we performed partitioned heritability using global functional connectivity and amplitude traits in resting fMRI. For both functional connectivity and amplitude traits, there was a much stronger enrichment in neuronal regulatory elements than in glia (**Fig. 6A**). For further resolution across brain networks, we also performed enrichment analysis for the mean amplitude of the 12 networks. In most functional networks, neurons were more enriched than glia, and the strongest neuronal enrichment was found in the posterior multimodal network (**Fig. 6B**). These findings may indicate that common variants associated with brain functional activity primarily affect the function of regulatory elements in neurons, which are expected to influence brain functional interactions.

**Fig. 6.**
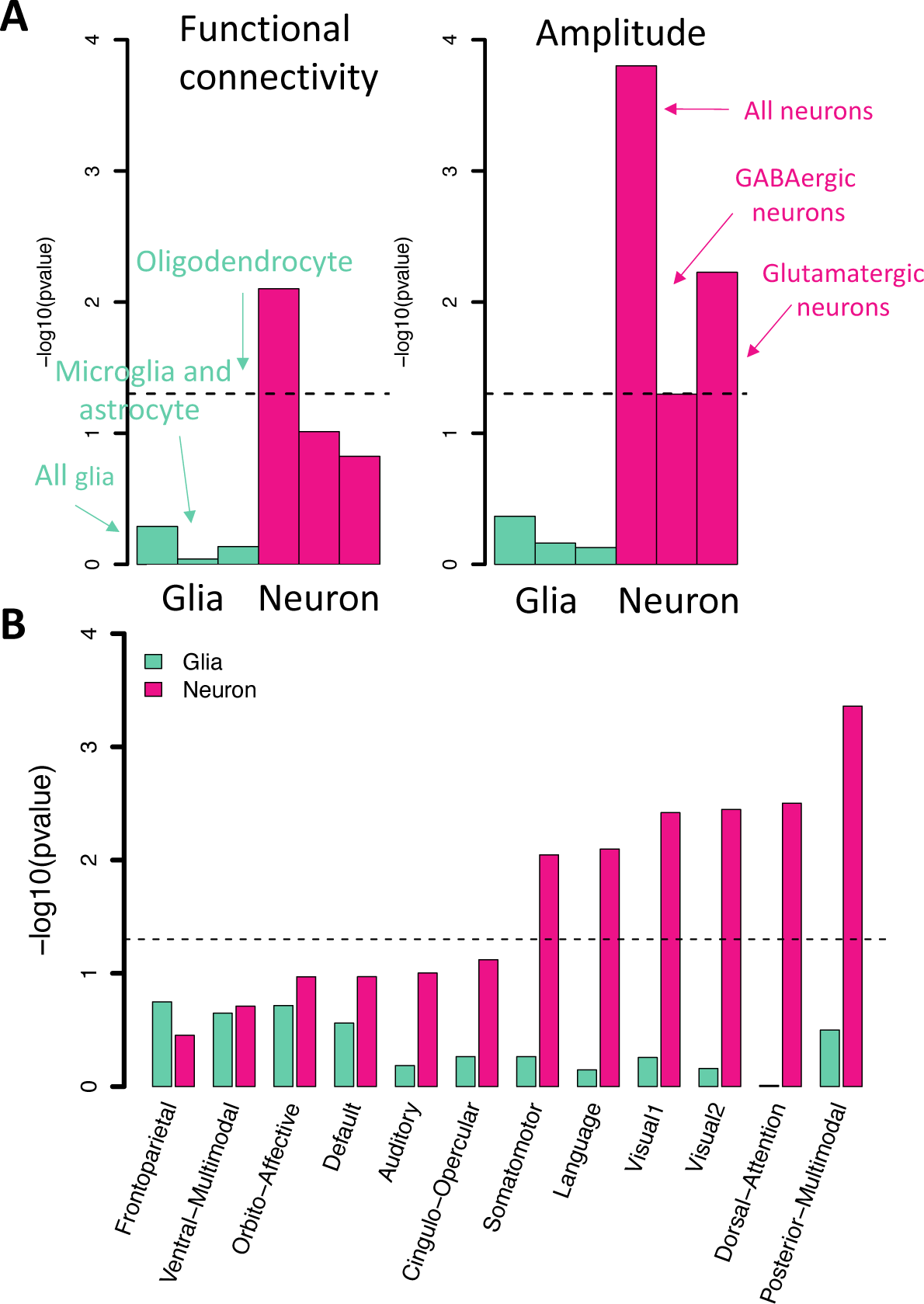
Partitioned heritability enrichment analysis. **(A)** Heritability enrichment of global functional connectivity and global amplitude of resting fMRI in regulatory elements of glial cells (glia, including all glial cells, oligodendrocyte subtype, and microglia/astrocyte subtype) and neuronal cells (neurons, including all neurons, GABAergic subtype, and glutamatergic subtype). The dashed lines indicate the nominal significance level. **(B)** Heritability enrichment of mean amplitude of the 12 functional networks in regulatory elements of glial and neuronal cell types.

## DISCUSSION

Most previous large-scale genetic studies of brain imaging data have focused on brain structures. Nevertheless, brain functional traits could also connect genetic variations to mechanisms underlying behavioral differences^108^. Using resting and task fMRI data from the UKB study, we provided fine details of genetic influences on cerebral cortex functional architectures through a parcellation-based approach. We showed the similarities and differences in the genetic architecture of intrinsic and extrinsic functional organizations. Genetic colocalization and correlation analyses uncovered important brain functional areas and networks that were genetically implicated in specific diseases and traits. Our results may help guide future clinical research and applications in the field of neurodegenerative and neuropsychiatric disorders. For example, structural MRI traits^109^ (such as gray matter volumes) are frequently used as imaging biomarkers in practical applications of Alzheimer’s disease. However, we found substantial genetic links between the visual cortex function and Alzheimer’s disease, and there were stronger associations with *APOE* ε4 in fMRI traits than in structural MRI traits. These findings suggest that fMRI traits of the visual cortex may be used as endophenotypes of Alzheimer’s disease.

At the group mean level, prior literature has demonstrated that the intrinsic and extrinsic functional architectures are highly similar with small but consistent differences^44,110–113^. These task-related changes are essential for the human brain to adaptively alter its functionality via rapid changes in inter-regional functional connectivity^44^. Using large-scale individual-level data, we showed the overall genetic similarity between resting and task fMRI (e.g., mean genetic correlation = 0.7). Although the genetic differences between resting and task fMRI may be small, several lines of evidence suggest such differences could be important and are genetically related to cognition and brain diseases. For example, cognitive function had genetic correlations with the secondary visual and somatomotor networks during task performance, but not at rest.

Although many efforts have been made to understand the functional organizations of the human brain, there is no one widely-accepted standard pipeline for functional connectivity analysis in fMRI^114^. Our study is one of the first attempts to study the genetic architecture of brain functions using a parcellation-based approach in a biobank-scale dataset. The major difference between the whole brain ICA and parcellation-based approaches is that the ICA regions are typically distributed across multiple regions and different networks by combining their major variations. Compared to the recent whole brain ICA-based studies^31,33^, our parcellation-based approach was able to uncover high-resolution fine details on the genetic effect patterns and enable the comparison between intrinsic and extrinsic functional architectures. More importantly, our results suggest the usefulness of specific fMRI phenotypes for different brain disorders from a genetic perspective.

There are a few limitations in the present study. First, the UKB participants were mostly middle-aged to elderly Europeans. Nonlinear aging effects on brain functional connectivity have been widely observed^115^. We have used the ABCD study as a replication dataset, it is of great interest to further study the gene-age interactions and evaluate the generalizability of UKB results across the lifespan^116^. It is also interesting to investigate the population-specific genetic components when more large-scale fMRI data from global populations become available. Due to potentially different environmental influences and population-specific genetic effects, fMRI traits could have different heritability across ancestry groups. Second, the UKB task fMRI data were from a single emotion processing task^36,37^. Although previous studies have shown that the functional architectures of different tasks were highly similar^44,110,112^, multi-task fMRI data may provide new insights in genetic studies. It might be possible to impute/predict multi-task fMRI data for UKB using the multi-task HCP data as training reference panels. To allow comparison between resting and task fMRI, our study focused on functional connectivity traits. It is also of great interest to study genetics influences on task activation measures/maps for task fMRI. Finally, this study partitioned the cerebral cortex using the Glasser360 atlas. By applying more brain parcellations^3^ to the large-scale UKB dataset, it will be possible to establish more useful imaging phenotypes for specific brain disorders.

## METHODS

Methods are available in the ***Methods*** section.

*Note: One supplementary information pdf file, one supplementary figure pdf file, and one supplementary table zip file are available*.

## ACKNOWLEDGEMENTS

We thank Fidel Alfaro-Almagro and Doug Crabill for their helpful conversations. The study has been supported by start-up funds from Purdue Statistics Department and funding from the Analytics at Wharton. This research was partially supported by U.S. NIH grants MH086633 (HT.Z.) and MH116527 (TF.L. and HP.Z.). Assistance for this project was also provided by the UNC Intellectual and Developmental Disabilities Research Center (NICHD; P50 HD103573; Y.L.). We thank the individuals represented in the UK Biobank study for their participation and the research teams for their work in collecting, processing, and disseminating these datasets for analysis. We would like to thank the research computing groups at the University of North Carolina at Chapel Hill, Purdue University, and the Wharton School of the University of Pennsylvania for providing computational resources and supports that have contributed to these research results. We gratefully acknowledge all the studies and databases that made GWAS summary data available. This research has been conducted using the UK Biobank resource (application number 22783), subject to a data transfer agreement.

## AUTHOR CONTRIBUTIONS

B.Z., HT.Z., J.L.S., and S.M.S. designed the study. B.Z., TF.L., Y.Y., X.Y., YL.Y., D.X., X.W., Z.Z., TY. L, Z.W., YJ.L., and Z.F. analyzed the data. TF. L., Z.Z., and Y.S. downloaded the datasets, processed fMRI data, and undertook quantity controls. Y.L. and HP.Z. provided feedback on the study design and results interpretation. B.Z. wrote the manuscript with feedback from all authors.

**CORRESPONDENCE AND REQUESTS FOR MATERIALS** should be addressed to HT.Z.

## COMPETING FINANCIAL INTERESTS

The authors declare no competing financial interests.

## METHODS

### Imaging datasets

We used the raw resting and task fMRI data from the UKB study, as well as raw resting fMRI data from the ABCD study. The UKB study obtained ethics approval from the North West Multicentre Research Ethics Committee (approval number: 11/NW/0382). All procedures in the ABCD study were approved by the institutional review boards at ABCD collection sites (approval numbers: 201708123 and 160091). The image acquisition and preprocessing procedures were detailed in the **Supplementary Note**. The UKB task fMRI study implemented the emotion processing task^36,37^. In this study, the time series were extracted from the whole scan of the task fMRI data (including blocks of both Shape and Face activations, Data-Category 106). Thus, all contrasts were analyzed together when we calculated the functional connectivity measurements in downstream analysis. We used a parcellation-based approach based on the Glasser360 atlas^1^. Briefly, for each subject, we projected the resting and task fMRI data onto the Glasser360 atlas and obtained the 360 × 360 functional connectivity matrices. The original Glasser360 atlas is a surface-based parcellation for the cerebral cortex^117^ and it has been transformed into a volumetric atlas that is compatible with the UKB volume-based data (**Supplementary Note**). The 360 functional areas were grouped into 12 functional networks^39^ (**Table S1**). To aid interpretation, the 360 functional areas were labeled using the automated anatomical labeling atlas^118^. We examined the spatial overlaps between the Glasser360 functional areas and the whole brain ICA-defined brain regions generated from the previous 25-dimension and 100-dimensions ICA analyses^30^ (**Figs. 1B** and **S2**).

We mainly studied two sets of fMRI traits: area-level traits and network-level traits. Area-level traits were the 8,531 functional connectivity measurements among all area pairs within each of the 12 networks, which provided fine details on cerebral cortex functional organizations and enabled the comparison between intrinsic and extrinsic functional architectures. Network-level traits were expected to aggregate the major information from the area-level traits. We input area-level functional connectivity traits and extracted their low-rank representations/phenotypes via a combined PCA and ICA^29^ dimension reduction approach in a training-validation design^31^. The low-rank imaging phenotypes were linear combinations of area-level traits and were independent of each other. We used a training-validation design (across two sets of independent subjects) to optimize both the initial dimensionality in the PCA step and the final number of ICA components (with the reproducibility of ICA weight vectors > 0.9). Specifically, the Glasser360 atlas classified 360 functional areas (nodes) into 12 networks, resulting in 360 × 359/2 = 64,620 functional connectivity traits between all pairs of functional areas (edges). The 12 networks allowed for categorizing all edges into 78 network-based groups, including 12 within-network groups, such as the default mode group, where both nodes of the edge were in the default mode network. There were 66 between-network groups, such as the sensorimotor-default mode group, where the two nodes of an edge came from sensorimotor and default mode networks, respectively. The top ICA components were extracted from each of the 78 network-based groups, which were interpreted as the network-level functional connectivity traits. These ICA components have been mapped back to the 360 functional areas of the Glasser360 atlas and visualizations have been provided to facilitate biological interpretations (**Fig. S78**). Additionally, we considered the mean amplitude of each network (12 networks in total), which was a measure of brain activity^40^. The network-level feature extraction was performed separately for resting and task fMRI. Together, there were 1,985 network-level traits, 1,066 of which were from resting fMRI and 919 from task fMRI. Detailed steps of our parcellation-based dimension reduction procedure can be found in the **Supplementary Note** and the overview of the procedure and study design can be found in **Figure S1**. Overall, the network-level amplitude traits described the average brain activity across all brain functional areas within the network; and the network-level functional connectivity traits quantified the strength of functional connectivity within each network or between two different networks.

We analyzed the following datasets separately: 1) the white British discovery GWAS, which used data of individuals of white British ancestry in UKB phases 1 to 3 data (*n* = 34,641 for resting and 32,144 for task, released up through 2020); 2) European validation GWAS: UKB white but non-British individuals in phases 1 to 3 data and all White individuals in newly released UKB phases 4 and 5 data (UKBW, *n* = 8,197 for resting and 7,684 for task); 3) two non-European UKB validation GWAS: UKB Asian (UKBA, *n* = 517 for resting and 429 for task) and UKB Black (UKBBL, *n* = 283 for resting and 231 for task); and 4) the UKB first revisit data (*n* = 1,491 for resting and 1,362 for task). The average age (at imaging) of all subjects was 64.16 (standard error = 7.73), 51.6% were females. The assignment of ancestry in UKB was based on self-reported ethnicity (Data-Field 21000), which was verified in Bycroft, et al. ^119^. We have also used the resting fMRI data from the ABCD study as non-UKB replication datasets (*n* = 3,821 for European, 768 for Hispanic, and 1,257 for African American cohorts). Details of ABCD data processing can be found in **Supplementary Note**.

### Heritability and GWAS analysis

We downloaded the imputed data from UKB data resources^119^ (Data-Category 263). We performed the following quality controls on subjects with both imaging and genetics data: 1) excluded subjects with more than 10% missing genotypes; 2) excluded variants with minor allele frequency less than 0.01; 3) excluded variants with missing genotype rate larger than 10%; 4) excluded variants that failed the Hardy-Weinberg test at 1 × 10^-7^ level; and 5) removed variants with imputation INFO score less than 0.8. SNP heritability was estimated by GCTA^41^ using all autosomal SNPs in the white British discovery GWAS. We adjusted the effects of age (at imaging), age-squared, sex, age-sex interaction, age-squared-sex interaction, imaging site, the top 40 genetic principal components (PCs)^119^, as well as the head motion, head motion-squared, brain position, brain position-squared, and volumetric scaling. We also examined the genetic variance estimates from the GCTA. Genome-wide association analysis was performed in linear mixed effect models using fastGWA^120^, while adjusting the same set of covariates as in GCTA. GWAS were also separately performed via Plink^121^ in validation datasets, where we adjusted for the top ten genetic PCs instead of the top 40. We removed values greater than five times the median absolute deviation from the median for each continuous phenotype or covariate variable. For area-level traits, the independent lead variants were clumped by Plink (LD *r*^2^ < 0.1,--clump-r2 0.1--clump-kb 250). The genomic loci associated with network-level traits were defined using FUMA^100^ (version 1.3.5e). Specifically, to define the LD boundaries, FUMA identified independent significant variants, which were defined as variants with a *P*-value smaller than the predefined threshold and were independent of other significant variants (LD *r*^2^ < 0.6). FUMA then constructed LD blocks for these independent significant variants by tagging all variants in LD (*r*^2^ ≥ 0.6) with at least one independent significant variant and had a MAF ≥ 0.0005. These variants included those from the 1000 Genomes reference panel that may not have been included in the GWAS. Moreover, within these significant variants, independent lead variants were identified as those that were independent of each other (LD *r*^2^ < 0.1). If LD blocks of independent significant variants were close (< 250 kb based on the closest boundary variants of LD blocks), they were merged into a single genomic locus. Thus, each genomic locus could contain multiple significant variants and lead variants. Independent significant variants and all the variants in LD with them (*r*^2^ ≥ 0.6) were searched on the NHGRI-EBI GWAS catalog (version 2019-09-24) to look for previously reported associations (*P* < 9 × 10^-6^) with any traits. We performed association analysis to illustrate association patterns for selected colocalized index variants across all 64,620 functional connectivity traits in resting and task fMRI. The significance threshold was set to be 3.86 × 10^-7^ (0.05/(64,620 × 2)). The same set of covariates used in the above GWAS analysis was adjusted in this analysis. LDSC^49^ (version 1.0.1) was used to estimate and test genetic correlations. We used the pre-calculated LD scores provided by LDSC, which were computed using 1000 Genomes European data. We used HapMap3 variants and removed all variants in the major histocompatibility complex (MHC) region.

We performed several additional heritability and GWAS analyses. First, we used the Yeo-7 atlas^51^ to categorize the 360 Glasser areas into 7 functional networks. We applied the same dimension reduction procedure as we used for the 12 functional networks^39^, and generated 615 network-level traits (294 for resting and 321 for task). Using the same set of covariates as we used in the main analysis, we performed SNP heritability and GWAS analyses for these Yeo-7 network-level traits. Second, we implemented additional quality control procedures to remove potentially poor-quality images. After these additional quality controls, we rerun our heritability and GWAS analyses to examine the robustness of the results reported in our main analyses. Details of the Yeo-7 traits generation and additional quality controls can be found in the **Supplementary Note**. Furthermore, we split our discovery GWAS sample into two independent parts and performed GWAS separately to evaluate genetic effect estimates in two datasets with equal sample sizes and homogenous genetic backgrounds.

### Gene-level analysis and biological annotation

Gene-based association analysis was performed in UKB white British discovery GWAS for 18,796 protein-coding genes using MAGMA^90^ (version 1.08). Default MAGMA settings were used with zero window size around each gene. We then carried out FUMA functional annotation and mapping analysis, in which variants were annotated with their biological functionality and then were linked to 35,808 candidate genes by a combination of positional, eQTL, and 3D chromatin interaction mappings. Brain-related tissues/cells were selected in all options and the default values were used for all other parameters in FUMA. We also explored the overlaps with drug target genes using a nervous system drug target database^105^, which contained 241 target genes for 273 nervous system drugs. We tested for whether the number of overlapping genes was greater than the expected number by chance using the Chi-squared test with the resampling-based *P*-value. We performed heritability enrichment analysis via partitioned LDSC^106^. Baseline models were adjusted when estimating and testing the enrichment scores for our brain cell type-specific annotations.

## Code availability

We made use of publicly available software and tools. The codes used in fMRI preprocessing and the parcellation-based network-level feature extraction pipeline will be shared on Zenodo.

## Data availability

The individual-level data used in the present study can be applied from the UK Biobank (https://www.ukbiobank.ac.uk/) and ABCD (https://abcdstudy.org/) studies. Our GWAS summary statistics will be shared on Zenodo and BIG-KP https://bigkp.org/. 3D visualizations of ICA components can be downloaded at: https://www.dropbox.com/s/np0rjlzvfa4fgae/Resting_fMRI_ICA.zip?dl=0 (for resting fMRI) and https://www.dropbox.com/s/rg053royl62z12v/Task_fMRI_ICA.zip?dl=0 (for task fMRI).

## Notes

### Competing Interest Statement

The authors have declared no competing interest.

### Author Declarations

The UKB study obtained ethics approval from the North West Multicentre Research Ethics Committee (approval number: 11/NW/0382). All procedures in the ABCD study were approved by the institutional review boards at ABCD collection sites (approval numbers: 201708123 and 160091).

### Summary of Updates

Update the data analysis to include more data from the UK Biobank and ABCD studies.

