## Supplementary material for "Genetic influences on the intrinsic and extrinsic functional organizations of the cerebral cortex": supp_information

### Supplementary Information for:
### Genetic influences on the intrinsic and extrinsic functional organizations of the cerebral cortex

October 30, 2023

**This PDF file includes:**

Supplementary Note
Captions for Table S1 to S13

**Other Supplementary Information for this manuscript includes the following:**

Figs. S1 to S78 (available in a PDF file)
Tables S1 to S13 (.xlsx, available in a ZIP file)

### Supplementary Note

#### Image acquisition and preprocessing

This work made use of resting-state and task-evoked functional magnetic resonance imaging (fMRI) data from the UK Biobank (UKB) study. The image acquisition and preprocessing procedures were detailed in the UKB Brain Imaging Documentation (`https://biobank.ctsu.ox.ac.uk/crystal/crystal/docs/brain_mri.pdf`). Below we briefly introduced the image acquisition and preprocessing procedures.

**UKB fMRI acquisition** All UKB brain imaging datasets, including the resting-state fMRI (rsfMRI), the task-evoked fMRI (tfMRI), and the T1-weighted structural MRI (sMRI) data, were acquired from standard Siemens Skyra 3T scanners. The rsfMRI data were acquired at 490 time points and a duration of 6 minutes, each with a 2.4×2.4×2.4 mm spatial resolution at a dimension of 88×88×64. The gradient-echo echo-planar imaging (GE-EPI) was adopted with a multiband factor of 8, no iPAT, flip angle 52°, and fat saturation. The echo time (TE) and repetition time (TR) were 39 ms and 735 ms, respectively. As implemented in the CMRR multiband acquisition (Moeller et al., 2010), a separate "single-band reference scan" was also acquired. This had the same geometry (including EPI distortion) as the time series data, but had higher between-tissue contrast to noise, and was used as the reference scan in head motion correction and alignment to other modalities (Alfaro-Almagro et al., 2018). The tfMRI data adopted the Hariri faces/shapes "emotion" task which has been implemented in the Human Connectome Project (HCP) tasks but with shorter overall duration and hence fewer total stimulus block repeats (`https://biobank.ndph.ox.ac.uk/showcase/label.cgi?id=106`). Participants were presented with blocks of face or shape trials and asked to decide which of two faces (or shapes) presented on the bottom of the screen match the face (or shape) at the top of the screen. The tfMRI data were acquired at 332 time points within a duration of 4 minutes. Other scanning parameters including the spatial resolution and dimension, multiband factor, TR and TE were the same as the UKB rsfMRIs. The ePrime stimulus script is available at `http://biobank.ctsu.ox.ac.uk/crystal/refer.cgi?id=1462`. In addition, the T1-weighted sMRI scans were acquired at the isotropic resolution of 1 mm and a dimension of 208×256×256 matrix, with a TR of 2000 ms, an inversion time of 880 ms, a TE of 2 ms, an in-plane acceleration of 2, and a scan time of 5 min using straight sagittal orientation.

**UKB image preprocessing** The UKB rsfMRI and tfMRI data were preprocessed by the UKB brain imaging team (Alfaro-Almagro et al., 2018). The full pipeline can be found in Section 3 of the UKB Brain Imaging Documentation. Overall, the pipeline included three parts: image cleaning, image registration, and representative time series generation. The source codes have been shared by the UKB brain imaging team at `https://git.fmrib.ox.ac.uk/falmagro/UK_biobank_pipeline_v_1`.

For rsfMRI, the image-cleaning workflow in the UKB preprocessing pipeline included the following steps: motion correction using MCFLIRT (Jenkinson et al., 2002); grand-mean intensity normalization of the entire 4D dataset by a single multiplicative factor; highpass temporal filtering (Gaussian-weighted least-squares straight line fitting, with a sigma of 50.0 s); EPI unwarping; and

GDC unwarping. Finally, structured artifacts were removed by the ICA+FIX processing (i.e., independent component analysis (ICA) followed by FMRIB's ICA-based X-noiseifier (Beckmann and Smith, 2004; Salimi-Khorshidi et al., 2014; Griffanti et al., 2014)). FIX was hand-trained on 40 UKB rsfMRI subjects by the UKB brain imaging team. The image registration part had the following steps. First, they aligned the GDC unwarped rsfMRI data from the previous step with the high-resolution T1 MRI image. The EPI unwarping in the last step already included an alignment to the T1, though the unwarped data was written out in native (unwarped) fMRI space (and the transform to T1 space was written out separately). This T1 alignment was carried out by FLIRT, with a final BBR cost function (Greve and Fischl, 2009). After the fMRI GDC unwarping, a final FLIRT realignment to T1 was applied, which toke into account any shifts resulting from the GDC unwarping. Second, they registered the T1 MR image for each individual to the standard MNI152 2×2×2 mm space. Third, they combined the two image warping together, conducted a transformation from the GDC unwarped fMRI space to the MNI standard space, and registered the cleaned fMRI data from the previous step to the MNI standard space by applying the combined image warping. The above three steps were completed in the fMRI expert analysis tool (FEAT) from the software FSL.

For tfMRI, the preprocessing steps included cleaning and registration of tfMRIs to the standard space (similar to the rsfMRI) and applying spatial smoothing with a Gaussian kernel of 5 mm FWHM and intensity normalization. Task-induced activation was then modeled using FEAT (FMRI Expert Analysis Tool) and analyzed using FILM with local autocorrelation correction (Woolrich et al., 2001). Z-statistics of the group-level fixed effect for three activation contrasts (Shapes, Faces, and Faces-Shapes) were generated and the Z-statistics were thresholded at $Z >$ 120. Therefore, the activation maps for the Shapes, Faces, and Faces-Shapes activation contrast were the three thresholded Z-statistics: 1) Shapes group-level fixed effect Z-statistics, thresholded at $Z > 120$; 2) Faces group-level fixed-effect Z-statistics, thresholded at $Z > 120$; 3) Faces-Shapes group-level fixed-effect Z-statistics, thresholded at $Z > 120$. More details can be found in the UKB Brain Imaging Documentation.

**Additional quality controls in sensitivity analysis** In our main analyses, we followed the above fMRI processing and basic quality control steps used by the UKB brain imaging team and adjusted for the covariates they released and recommended. Briefly, 1) the UKB brain imaging team estimated the mean framewise displacement (averaged across the brain) for each consecutive pair of time points in both rsfMRI and tfMRI data. These estimates were then averaged across all time points to obtain the head motion parameters, which were included as covariates in our analyses. 2) In consideration of the signal dropouts, FMRIB's ICA-based X-noiseifier (Beckmann and Smith, 2004; Salimi-Khorshidi et al., 2014; Griffanti et al., 2014)) was trained and applied. 3) The UKB brain imaging team has identified unusable MRI scans with semi-automated quality control methods, and these images were not included in our analyses.

In our sensitive analyses, we have further implemented an additional set of quality control steps and examined their influences on our genetic analyses. First, we calculated the framewise displacement by ourselves and applied filters to remove outliers. Specifically, the rfMRI and tfMRI framewise displacement at each time point were first calculated following (Power et al., 2012), using the translation and rotation parameters at each consecutive pair of time points generated by MCFLIRT (Jenkinson et al., 2002). If the framewise displacement of 30% of the scans from the fMRI image were larger than 0.5 mm (Power et al., 2014; Mehler et al., 2020), or the mean framewise displacement was larger than 0.5 mm, we considered this image had bad quality and removed it from the analyses. Second, to detect anatomical outliers, we have used T1-to-standard nonlinear alignment discrepancy (UKB data field: 25732), the discrepancy between rfMRI brain image and T1 brain image (UKB data field: 25739), and the discrepancy between tfMRI brain image and T1 brain image (UKB data field: 25740) as additional quality control measures. In addition, we added additional quality control steps based on the temporal signal-to-noise ratio (TSNR) for both the rfMRI (UKB data field: 25744) and tfMRI (UKB data field: 25745). If the discrepancy or the inverted TSNR from one image exceeded 5 times the median absolute deviation from the median, it was considered an outlier and was excluded from downstream analyses.

In summary, the additional QC procedures included framewise displacement checking, T1-to-standard nonlinear alignment discrepancy, rfMRI-T1 and tfMRI-T1 alignment discrepancy, as well as the rfMRI TSNR and tfMRI TSNR. Subjects were excluded if they met any of the following criteria: 1) identified as unusable by the UKB, 2) 30% of the scans from the fMRI image with a framewise displacement larger than 0.5 mm, 3) the mean framewise displacement was larger than 0.5 mm, 4) the T1-to-standard nonlinear alignment discrepancy or fMRI-T1 alignment discrepancy exceeded 5 times the median absolute deviation from the median, or 5) the inverted rfMRI TSNR or the inverted tfMRI TSNR exceeded 5 times the median absolute deviation from the median. We removed 492 subjects from rfMRI and 993 subjects from tfMRI for sensitivity analysis after all these steps were completed.

**The Glasser360 atlas and Ji-12 networks** The Glasser360 atlas (Glasser et al., 2016) is a detailed group-based parcellation constructed using both high-resolution functional and anatomical MRI data from subjects of the HCP database. This atlas included 360 cortical areas with 180-area per hemisphere. We used the Glasser360 volumetric parcellation (`https://identifiers.org/neurovault.collection:1549`), which was converted from the original surface-based atlas to a volumetric atlas by a series of conversion and transformation steps using FreeSurfer (Andreas, 2016). After the preprocessing steps in the above sections, this atlas was mapped onto the UKB registered and cleaned rsfMRI and tfMRI datasets to extract the 360 regional time series. The amplitude traits and the full connectivity matrices were extracted from the time series standard deviation and the Gaussianized temporal correlation between every node time series pairs, respectively. We then incorporated the networks defined in Ji et al. (2019), which split the 360 areas into 12 networks. Based on this Ji-12 network atlas, we generated network-level traits. First, the $360 \times 359/2$ area-leve pairs were separated into 78 network-level groups, including 12 within-network groups and $12 \times 11/2 = 66$ between-network groups. For example, the auditory and default networks had 15 and 77 areas, respectively, whereas the auditory default class was one of the between-network classes that included $15 \times 77$ area-level pairs. Based on the network-level groups, we extracted the ICA components, which were described in detail in the next section.

#### Parcellation-based network level feature extraction

According to the above section, there were 360 areas (nodes) classified into 12 networks in the Glasser360 atlas; and there were $360 \times 359/2 = 64,620$ connectivity traits (edges) between pairs of areas. These connectivity traits can be classified into 78 network-based groups, including 12 within-network clusters, such as the default-mode group where both nodes in an edge belonged to the default-mode network, and 66 between-network groups, such as the sensorimotor-default mode group where nodes in an edge belonged to the sensorimotor and default-mode networks, respectively. To mitigate high dimensional data and aggregate the regional functional connectivity information, we applied ICA-based dimension reduction (Shlens, 2014; Hyvarinen, 1999; Elliott et al., 2018) for each of the 78 network-based groups. Specifically, we assumed that the functional connectivities within each cluster can be represented as a linear mixture of $q$ independent signal sources, as expressed in the equation $\boldsymbol{Z} = \boldsymbol{AS}$, where $\boldsymbol{Z}$ is a connectivity matrix with rows representing subjects and columns representing connectivity traits from the group, $\boldsymbol{S} = (\boldsymbol{S}_1, \ldots, \boldsymbol{S}_q)^T$ is an orthogonal matrix representing $q$ ICA bases, and $\boldsymbol{A}$ is a source signal matrix representing the ICA scores (linear weights on the signals). The model was solved using the combination of Principal Component Analysis (PCA) pre-whitening and the fast ICA algorithm. The use of PCA pre-whitening provides several benefits, including reduced computational complexity, data decorrelation, and an improvement in the performance of ICA through the elimination of insignificant trailing eigenvalues (Draper et al., 2003). This PCA+ICA dimension reduction approach has been widely applied in fMRI data analysis Calhoun et al. (2015); Hyvarinen (1999); Elliott et al. (2018) and the Melodic ICA tool of the FSL software. The reproducibility of each independent component was assessed across multiple datasets. Specifically, we trained the ICA bases in the UKB phases 1 and 2 data (data released up through 2018), and validated the ICA bases in the UKB phase 3 data (data released in early 2020). Independent components with poor reproducibility across different datasets were excluded from the final derived features. Briefly, the overarching process involved dividing the $360 \times 360$ functional connectivity matrix into 78 submatrices based on network affiliations of the 360 area for each subject. The top triangle of each within-network submatrix, or all elements of each between-network submatrix, were extracted and compiled into a person $\times$ edge matrix. This matrix was cleaned by excluding columns with low reproducibility, resulting in a refined person $\times$ edge matrix. Dimension reduction was then performed on this refined matrix to derive the ICA scores for each submatrix. More details on the ICA procedure were presented in the following steps. Importantly, as shown in later sections, each ICA component can be projected back onto the original 360 Glasser areas for visualization and interpretation purposes.

- Step 1. Image preprocessing.
  We preprocessed the raw fMRI and T1 data following the UKB pipeline mentioned in the above sections and generated the cleaned and aligned fMRI data for UKB phase 1, 2, 3, 4, 5, and re-visit datasets.

- Step 2. Generating the area-level connectivity traits.
  For each subject, each of the 360 areas of the brain, regional time series were extracted by taking the average from the voxelwise time series over each region. The correlation between

each two regional time series was Gaussianized from Pearson correlations (r-values) into z-statistics, including empirical correction for temporal auto-correlation. Then we obtained $360 \times 360$ functional connectivity matrix for each subject included in the above datasets, representing $64,620$ connectivity traits. This was carried out by the FSLNets toolbox (`http://fsl.fmrib.ox.ac.uk/fsl/fslwiki/FSLNets`).

- Step 3. Cleaning of area-level connectivity traits.
  The reproducibility of each of the $64,620$ regional connectivity traits was calculated based on Pearson's correlation between the connectivity measures from the first and second visits of the revisiting subjects in the UKB retest dataset. RfMRI connectivity traits with reproducibility below 0.3 and tfMRI connectivities with reproducibility below 0.25 were excluded from subsequent analyses.

- Step 4. ICA decomposition.
  For each network-level group, we constructed the population connectivity matrix $\boldsymbol{Z}$, with rows representing subjects and columns representing the cleaned connectivity traits from this group. We applied the singular value decomposition on the functional connectivity matrix $\boldsymbol{Z} = \boldsymbol{U}\boldsymbol{\Lambda}\boldsymbol{V}^T$. We considered the largest $r$ eigenvalues of $\boldsymbol{\Lambda}$ and extracted the corresponding $r$ columns of $\boldsymbol{V}$ as matrix $\boldsymbol{V}_r$, representing the top $r$ PCA eigenvectors. The $\boldsymbol{V}_r$ was then decomposed into the ICA score and basis matrices $\boldsymbol{V}_r^T = \boldsymbol{W}_r{}^T\boldsymbol{S}$ using the fast ICA algorithm (Koldovsky et al., 2006), resulting in $\boldsymbol{Z} \approx \boldsymbol{U}\boldsymbol{\Lambda}_r\boldsymbol{W}_r{}^T\boldsymbol{S}$. This step was carried out for all 78 network-level groups across a list of tuning parameters $r$s.

- Step 5. ICA validation.
  For each network-level group, Step 4 was trained using UKB phases 1 & 2 and UKB phase 3 datasets, separately. For each ICA basis $\boldsymbol{S}_i, i = 1, 2, \ldots, q$, reproducibility was calculated using the correlation between the ICA basis calculated from the UKB phases 1 & 2 dataset and phase 3 dataset. Those ICA bases with reproducibility less than 0.9 were considered unstable. Tuning parameter $r$ (Step 4) was selected to maximize the number of stable bases.

- Step 6. ICA projection.
  For each network-level group, we projected the regional fMRI feature matrices $\boldsymbol{Z}$ onto the ICA basis matrix $\boldsymbol{S}$ generated from UKB phases 1 and 2 data (with unstable components excluded) and obtained the final network level ICA features $\boldsymbol{F} = \boldsymbol{X}\boldsymbol{S}^T$.

- Step 7. Within each of the 12 networks, the mean functional activity was calculated across all functional connectivity traits. An additional feature, the mean amplitude, was calculated as the standard deviation of the brain functional time series, which was a measure of brain activity. All the above steps were performed for rsfMRI and tfMRI, respectively.

- Step 8. Replacing the Ji-12 networks (Ji et al., 2019) with Yeo-7 networks (Yeo et al., 2011), we repeated the above Step 4 $\sim$ Step 7 to generate network-level traits for the Yeo-7 networks.

**Mapping the ICA components back to the 360 functional areas of the Glasser360 atlas** For the network-level ICA components derived in the above section, we have mapped them back to the 360 functional areas of the Glasser360 atlas by aggregating the weights of both nodes on each edge. Specifically, the matrix $\boldsymbol{S}$ represents the final mapping from ICA scores $\boldsymbol{U\Lambda W_r}^T$ to the original person × edge matrix $\boldsymbol{Z}$. We can draw $\boldsymbol{S}$ as a 360 × 360 matrix $\tilde{\boldsymbol{S}}$ (or network-based sub-matrices), to show how the ICA score weights were distributed at each edge of the 360 × 360 matrix. We then calculated the node-level weight for each functional area by summing up each column of $\tilde{\boldsymbol{S}}$. Examples of the mappings of ICA components were shown in Figure S78. We have also identified the top-ranking areas with the highest contributions to the ICA components, which cumulatively accounted for 30% of the total contribution. These results are summarized in Table S1 and were used to map ICA traits back to specific areas.

**Overlaps between Glasser360 atlas and whole brain-ICA maps** The UKB brain imaging team generated whole brain group ICA spatial maps by executing spatial ICA at two dimensionalities of 25 and 100, which were publicly accessible at `http://www.fmrib.ox.ac.uk/ukbiobank`. After manual examination, 49 artifact ICA components were excluded (4 out of 25 and 45 out of 100), while 76 usable ICA components were retained (Alfaro-Almagro et al., 2018). The ICA maps consisted of continuous ICA weights rather than binary masks, and they may contain multiple spatially separate peaks/regions. The Glasser360 atlas was aligned with these whole brain ICA spatial maps through the following steps: 1) The binary mask for each ICA component was created by extracting ICA weights greater than the 80th percentile of the robust intensity range (the range of intensities between the 2nd and 98th percentiles of the positive ICA weights). All voxels with intensities below the threshold were set to zero. 2) The overlap between each of the 360 functional areas from the Glasser360 atlas and the ICA mask for each ICA component was quantified by counting the number of voxels. Areas with $>$ 10% voxels of overlap with the mask of an ICA component were considered as regions with substantial overlaps with that ICA component.

**ABCD data analysis** We also used resting fMRI data from the ABCD study as additional replication data. The image acquisition and preprocessing procedures were detailed in Casey et al. (2018). We downloaded the minimally processed resting fMRI data and performed the same preprocessing steps as in Zhao et al. (2022). We then applied the above Glasser360 atlas and UKB-derived ICA components to ABCD datasets to generate the area-level and network-level resting fMRI traits. Following Zhao et al. (2022), we used the Michigan Imputation Server (`https://imputationserver.sph.umich.edu/`) to perform genotype imputation and standard quality controls on the ABCD study. We removed relatives and adjusted for effects of age, age-squared, sex, head motion, age-sex interaction, age-squared-sex interaction, and top ten genetic PCs.

### Extended results of genetic correlation patterns

Snoring had negative genetic correlations with the cluster 7 of the default mode network (e.g., left/right TE2a in inferior temporal) and positive genetic correlations with the left 13l area (in posterior orbital) of the frontoparietal network (Fig. S75A). The sleep duration had negative genetic correlations with multiple networks (e.g., the somatomotor, secondary visual, and cingulo-opercular), most of the correlations were negative (Fig. S75B). General risk tolerance had positive genetic correlations with the right 5L area (in postcentral) of the somatomotor network (Fig. S75C). High blood pressure had negative genetic correlations with a few areas in the default model network, including the left/right s32 (in the medial orbital of superior frontal) and left/right 25 (in olfactory) (Fig. S76A). Education had genetic correlations with multiple networks, especially the default mode network (e.g., left/right POS1 in calcarine and right 7m in Precuneus) and the cingulo-opercular network (e.g., the right SCEF in supplementary motor area and left/right 6r areas in opercular part of inferior frontal) (Fig. S76B).

### Legends for Tables S1 to S13 (All tables can be found in a zip file).

**Table S1: The 360 functional areas defined in the Glasser360 atlas.** More details can be found in Glasser et al. 2016 (https://doi.org/10.1038/nature18933). The first 180 areas are for the left brain, and the second 180 areas are for the right brain.

**Table S2: SNP heritability estimates of 8,531 area-level within-network connectivity in resting-state fMRI and task-evoked fMRI.** We used UKB individuals of British ancestry (n = 34,641 for resting and 32,144 for task).

**Table S3: SNP heritability estimates of 1,985 network-level traits.** We used UKB individuals of British ancestry (n = 34,641 for resting and 32,144 for task).

**Table S4: Independent (LD $r^2 < 0.1$) significant associations for resting-state and task-evoked fMRI.** We used UKB individuals of British ancestry (n = 34,641 for resting and 32,144 for task).

**Table S5: Independent (LD $r^2 < 0.1$) significant associations for resting-state and task-evoked fMRI using network-level traits.** We used UKB individuals of British ancestry (n = 34,641 for resting and 32,144 for task).

**Table S6: Summary of loci identified by parcellation-based traits and whole brain ICA traits in resting fMRI.**

**Table S7: Summary of loci identified by diffusion MRI traits and fMRI traits.**

**Table S8: Summary of GWAS results in additional analyses.** We show the results in Yeo-7 network analysis, sensitivity analysis, and split-half analysis.

**Table S9: Independent significant variants and their correlated variants for fMRI traits that have previously been identified in GWAS of any traits listed in the GWAS catalog.** We listed all previously reported GWAS signals with P-value $< 9 \times 10^{-6}$ (the default threshold in FUMA).

**Table S10: Genetic correlation between fMRI traits and other complex traits.**

**Table S11: List of significant gene-level associations identified by MAGMA for network-level fMRI traits.** We reported the associations passing the Bonferonni correction ($P < 1.34 \times 10^{-9}$ for 1,985 network-level traits and $P < 1.55 \times 10^{-10}$ for $8,531 \times 2$ area-level traits).

**Table S12: List of mapped genes identifiedby FUMA at 2.51e-11 significance level.** We mapped the significant variants ($P < 2.51 \times 10^{-11}$) to genes via physical position, eQTL association, and 3D chromatin (Hi-C) interaction.

**Table S13: Nervous system drug-target genes overlapped with fMRI-associated genes.**
