## Supplementary material for "Genetic influences on the intrinsic and extrinsic functional organizations of the cerebral cortex": supp_figure

This PDF file includes:

Supplementary Figures S1 to S78

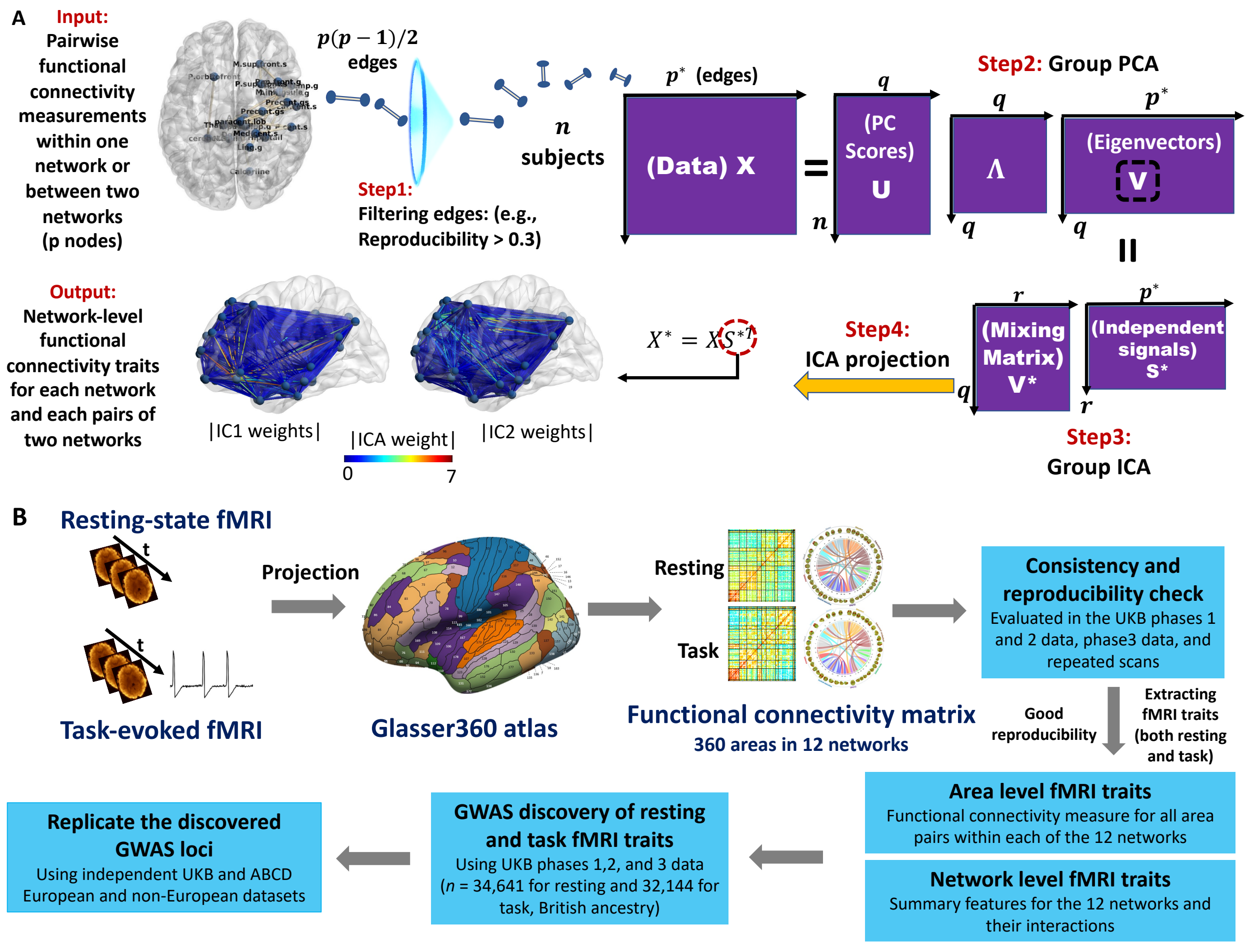


**Fig. S1 Visualization of the parcellation-based network-specific dimension reduction procedure (A) and the overview of study design (B).**

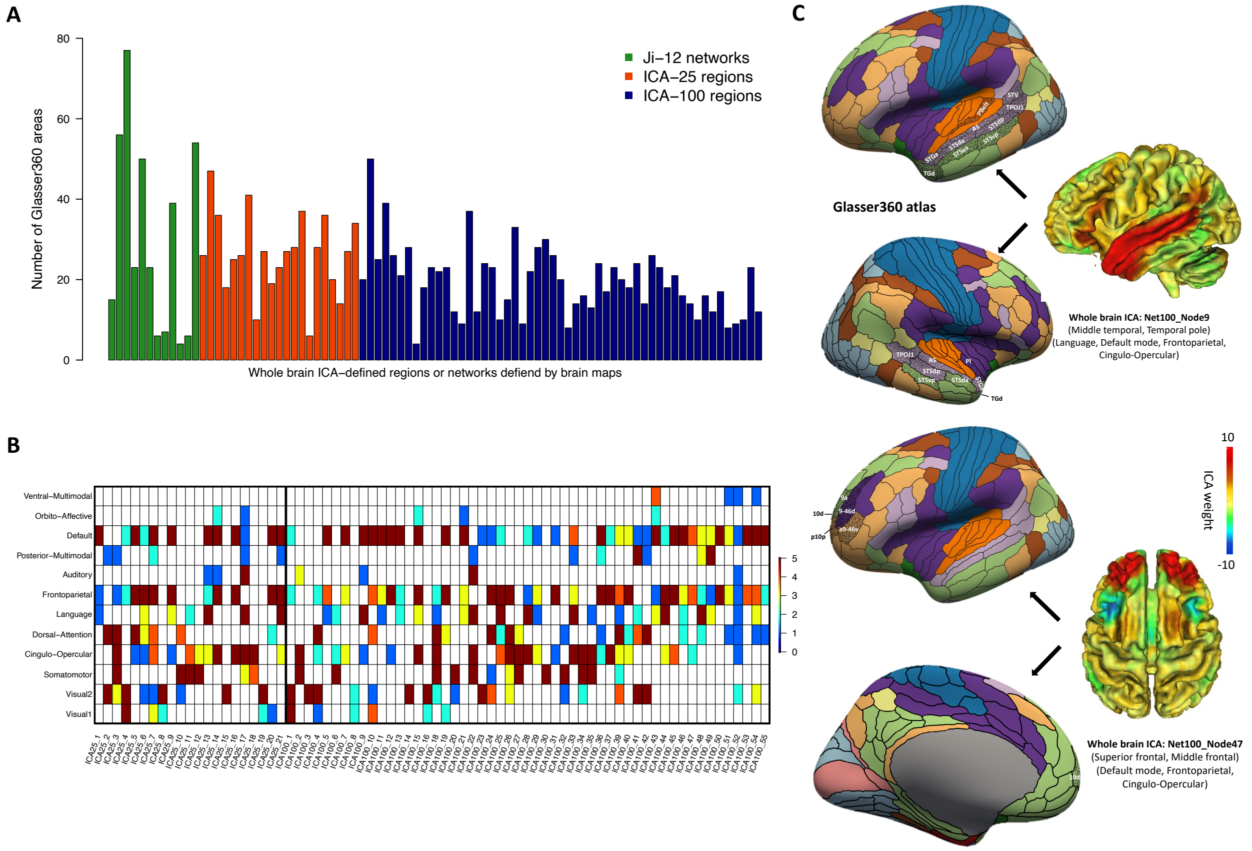


**Fig. S2 Comparison between the parcellation-based brain areas in the Glasser360 atlas and the selected whole brain ICA-defined brain regions.**

**A:** For each of the 12 networks in the present study (Ji-12-networks) and the 76 whole brain ICA-defined brain regions in previous GWAS (https://www.fmrib.ox.ac.uk/ukbiobank/), we illustrate the number of its overlapped Glasser360 areas. There are 21 ICA regions from the 25-dimensions ICA analysis (ICA-25) and 55 regions from the 100-dimensions ICA analysis (ICA-100). The mean is 30 for the 12 networks, 26.43 for the 21 ICA-25 regions, 18.87 for the 55 ICA-100 regions, and 20.97 for all 76 ICA regions. **B:** Number of Glasser360 areas overlapped between the 12 networks (y-axis) and ICA-defined brain regions (the x-axis). These results suggest most of these whole brain ICA-defined brain regions are distributed across multiple brain networks. **C:** We illustrate the spatial overlaps between the functional areas defined in the Glasser360 atlas and 2 selected the whole brain ICA-defined brain regions (Net100_Node9 and Net100_Node47, Glasser360 areas highlighted by dots with names being labeled). Specifically, the Net100_Node9 region (mainly in the middle temporal and temporal pole) overlapped with 18 areas in the language, default mode, frontoparietal, and cingulo-opercular networks, and the Net100_Node47 region (mainly in the superior frontal and middle frontal) overlapped with 6 areas in the default mode, frontoparietal, and cingulo-opercular networks.

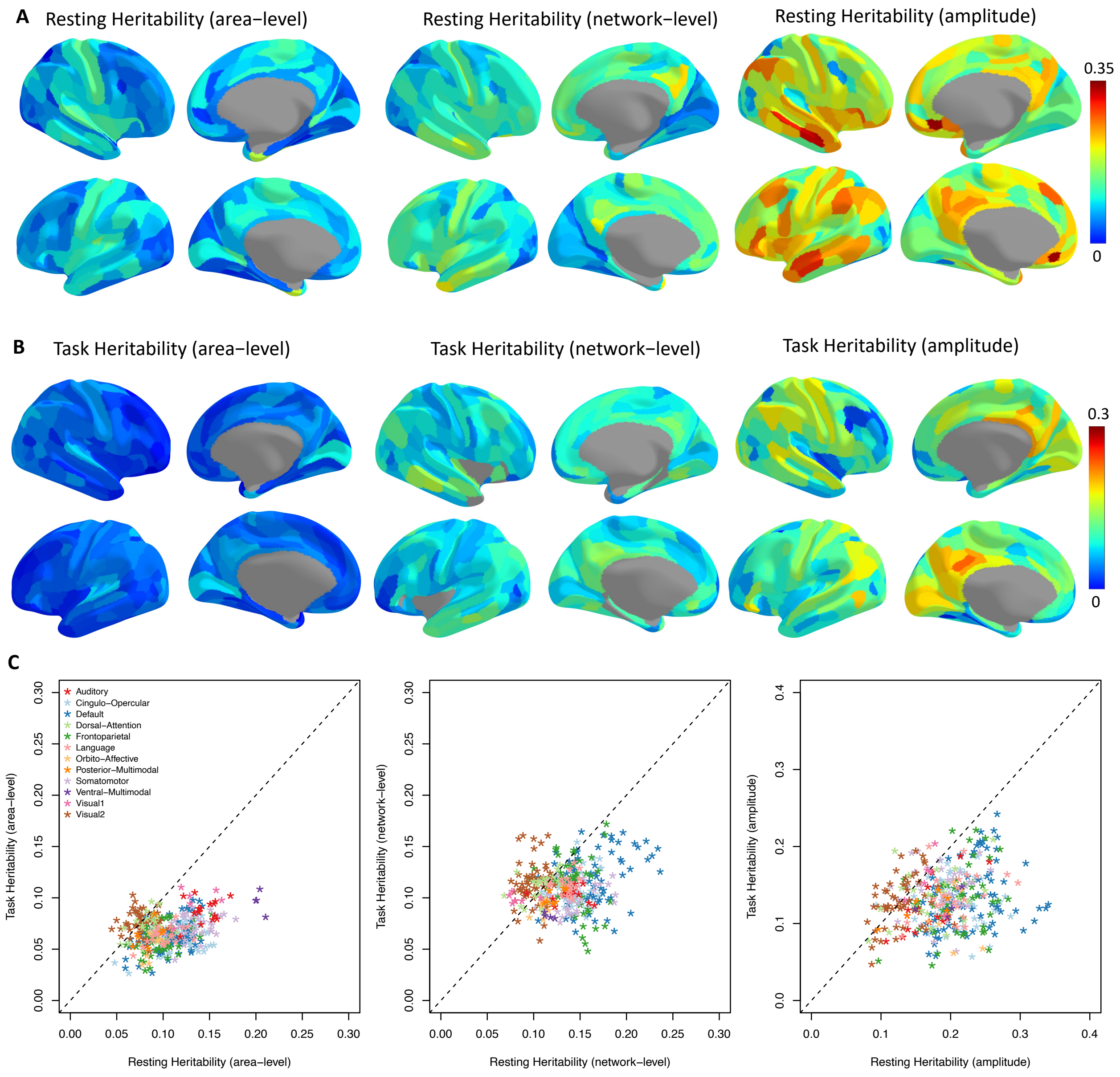


**Fig. S3: Surface maps of the mean heritability for each of the 360 cortical areas.**

The three top panels (**A**) illustrate the heritability maps for area-level traits (left), network-level traits (middle), and amplitude traits (right) in resting fMRI. The three middle panels (**B**) illustrate the maps for task fMRI. The three bottom panels (**C**) compare the resting fMRI and task fMRI results**.**

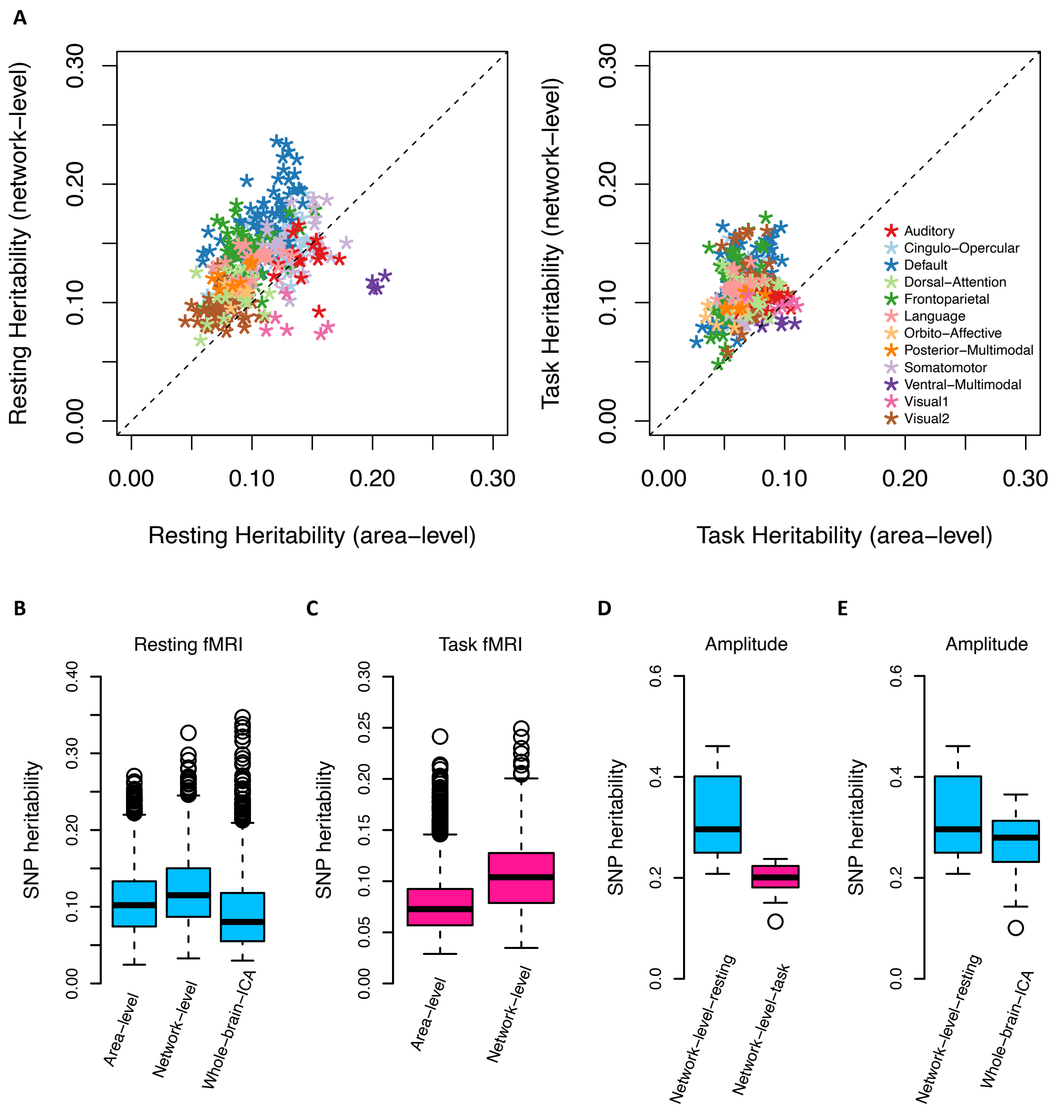


**Fig. S4 Summary of SNP heritability estimates.**

Heritability was estimated in both resting-state fMRI and task-evoked fMRI using UKB individuals of British ancestry (*n* = 34,641 for resting and 32,144 for task). **A:** Comparing the results of area-level and network-level traits after mapping them back to the 360 cortical areas. **B:** Comparison of the heritability estimates among area-level functional connectivity traits, network-level functional connectivity traits, and the whole brain ICA-based functional connectivity traits in resting fMRI. **C:** Comparison of the heritability estimates between area-level functional connectivity traits and network-level functional connectivity traits in task fMRI. **D:** Comparison of the heritability estimates between the network-level amplitude traits in resting and task fMRI. **E:** Comparison of the heritability estimates between network-level amplitude traits and the whole brain ICA-based amplitude traits in resting fMRI.

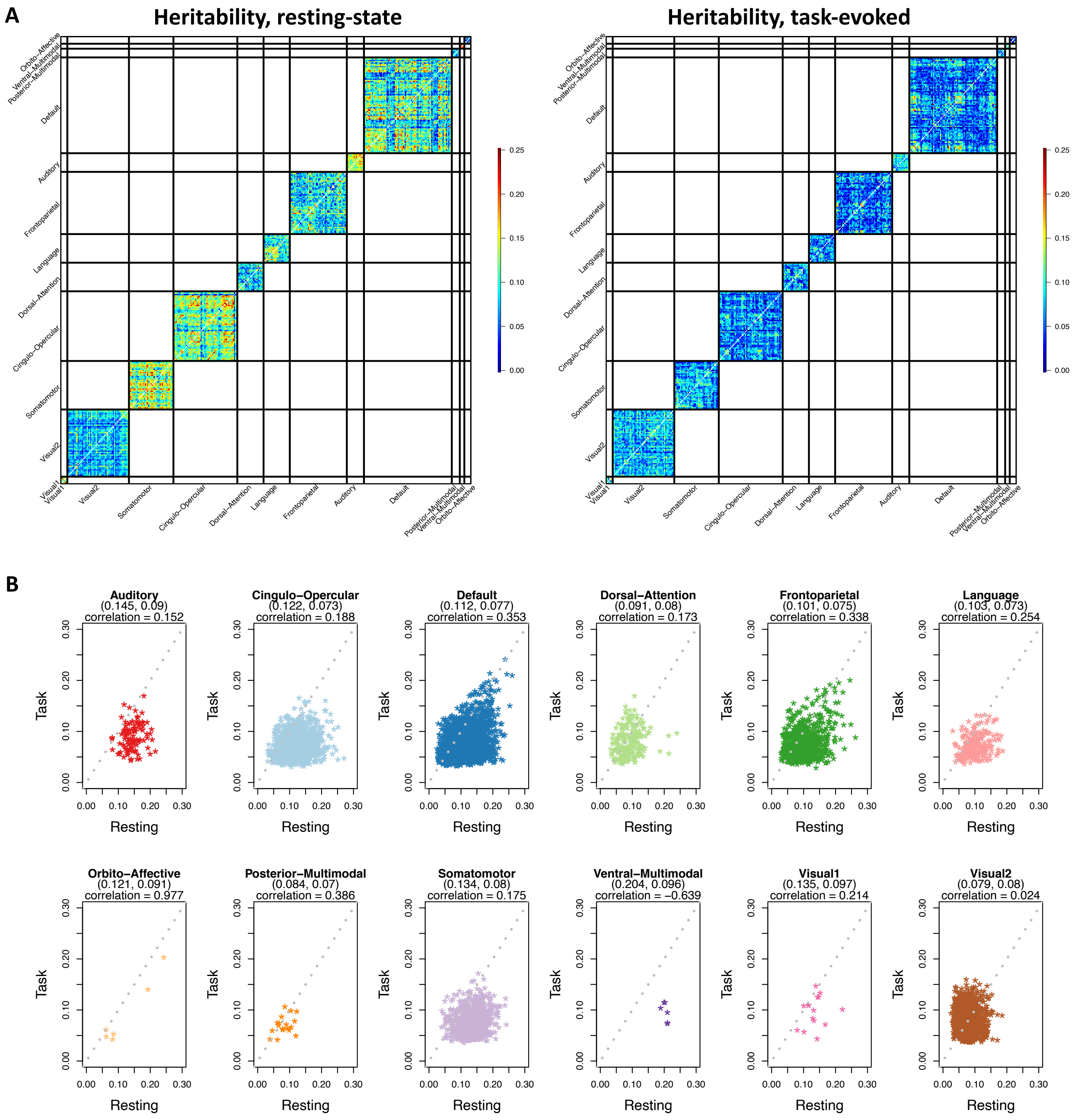


**Fig. S5 Heritability pattern of the area-level within-network functional connectivity traits.**

Heritability was estimated for the area-level within-network connectivity traits of each network in both resting-state fMRI and task-evoked fMRI using UKB individuals of British ancestry (*n* = 34,641 for resting and 32,144 for task). **A:** Heritability spatial map in resting fMRI (left) and task fMRI (right). **B:** Comparison of the resting and task heritability in each network. We reported the mean of resting heritability estimates and task heritability estimates as well as their correlation.

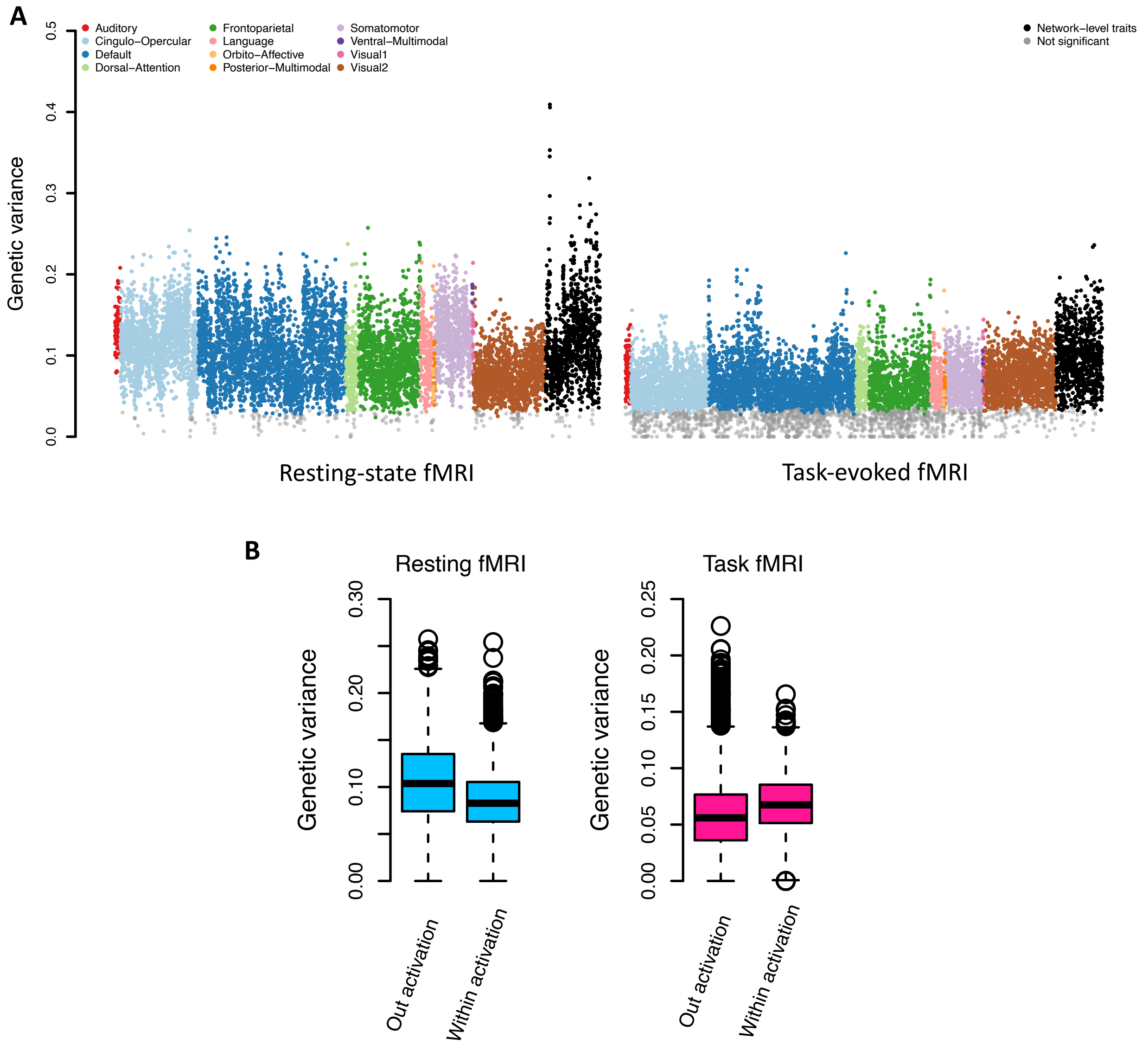


**Fig. S6 Genetic variance pattern in resting and task fMRI.**

**A**: The dots represent the genetic variance estimates of fMRI traits, including 8,531 area-level and 1,066 network-level traits in resting fMRI (left panel), and 8,531 area-level traits and 919 network-level traits in task fMRI (right panel). The dots of non-significant genetic variance estimates (after controlling false discovery rate at 5% level) had gray color. **B**: Comparison of genetic variance between the activated areas (within activation, defined by the shape contrast in task fMRI) and the nonactivated areas (out activation) in resting fMRI (left panel) and task fMRI (right panel).

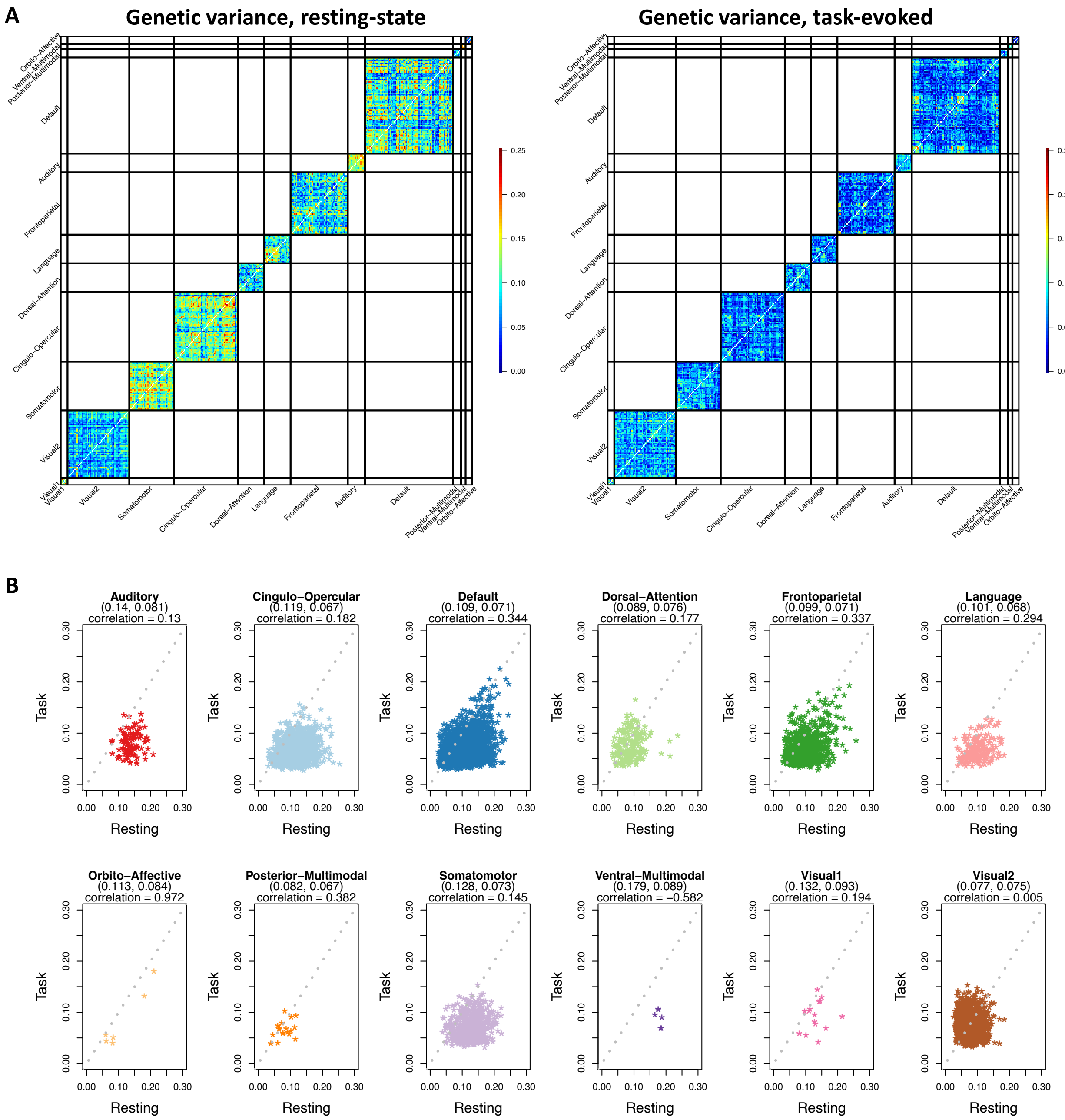


**Fig. S7 Genetic variance pattern of the area-level within-network functional connectivity traits.**

Genetic variance was estimated for the area-level within-network connectivity traits of each network in both resting-state fMRI and task-evoked fMRI using UKB individuals of British ancestry (*n* = 34,641 for resting and 32,144 for task). **A:** Genetic variance spatial map in resting fMRI (left) and task fMRI (right). **B:** Comparison of the genetic variance in resting fMRI and task fMRI for each network. We reported the mean of genetic variance estimates in resting fMRI and task fMRI as well as their correlation.

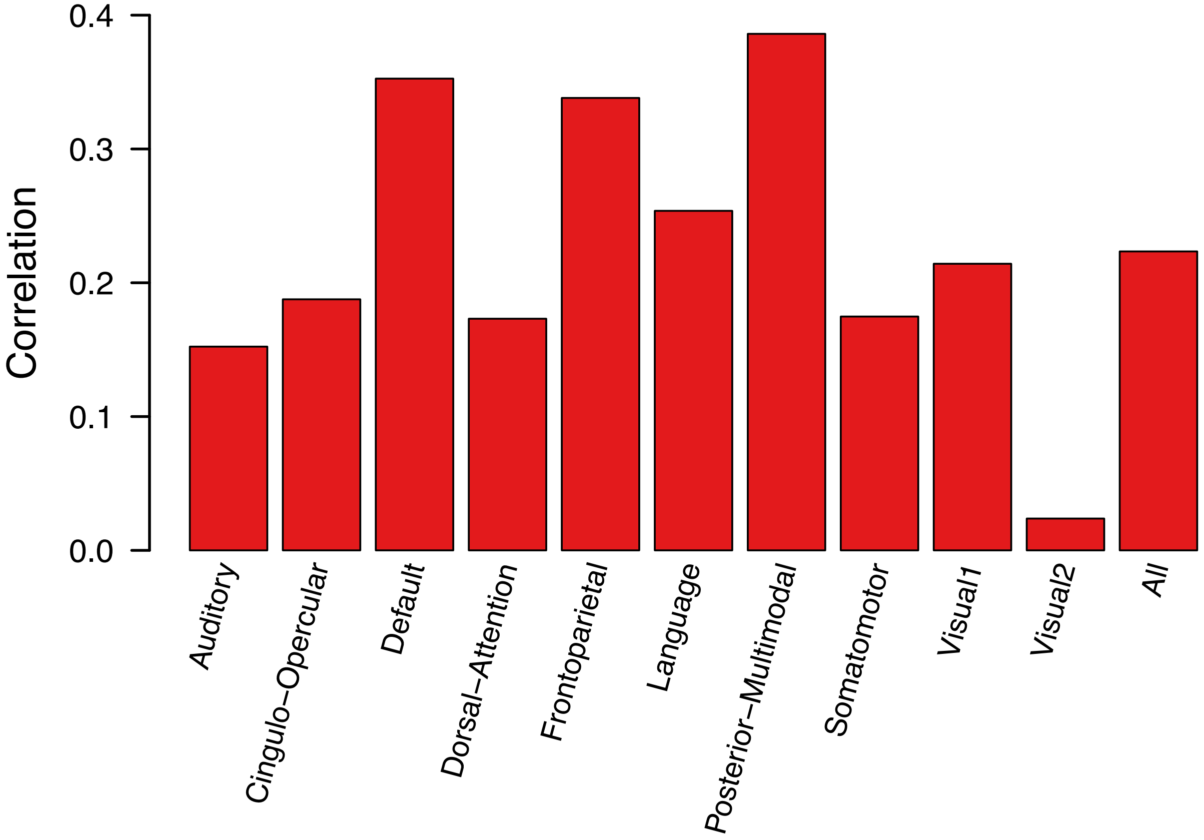


**Fig. S8 Correlation of heritability estimates between resting and task fMRI traits.**

We show the results for all area-level traits and separately for the networks with more than 10 significant heritability estimates.

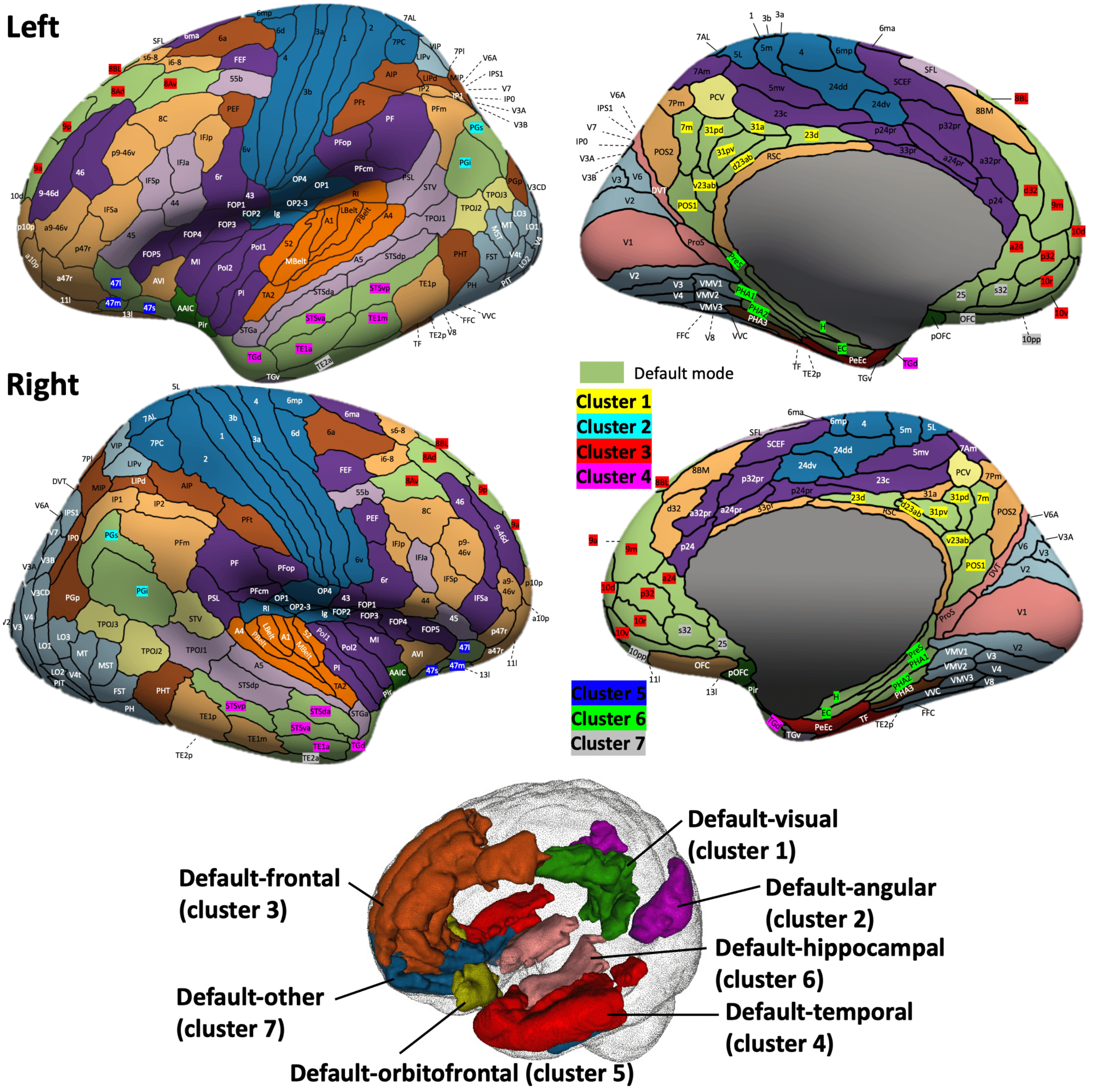


**Fig. S9 Location of the 7 clusters in the default mode network.**

We divided the functional areas in the default mode network into 7 clusters, mainly based on their locations in the brain. The cluster 1 was mainly in the visual cortex (precuneus, calcarine, and cingulate), the cluster 2 was mainly in the angular, the cluster 3 was mainly in the frontal, the cluster 4 was mainly in the temporal, the cluster 5 was mainly in the orbitofrontal, the cluster 6 was mainly in the hippocampus, and the cluster 7 included other areas.

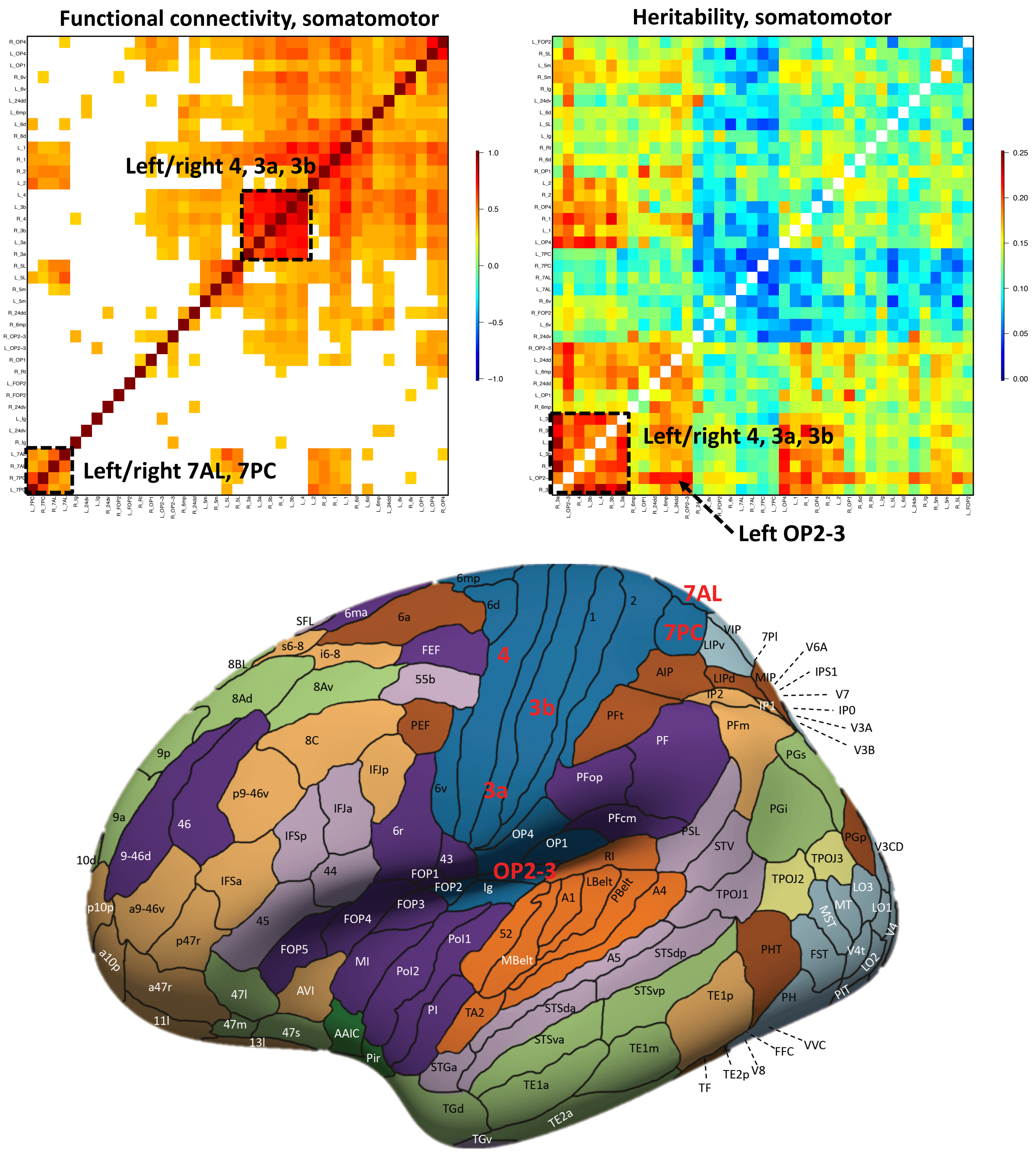


**Fig. S10 Heritability of the somatomotor network in resting-state fMRI.**

Top left: the functional connectivity pattern of the somatomotor network in resting-state fMRI. Top right: significant heritability estimates of the functional connectivity traits in the somatomotor network. Bottom: location of the highlighted areas that had the highest heritability (left hemisphere). Background colors indicate different functional networks, whose names can be found in Figure S1.

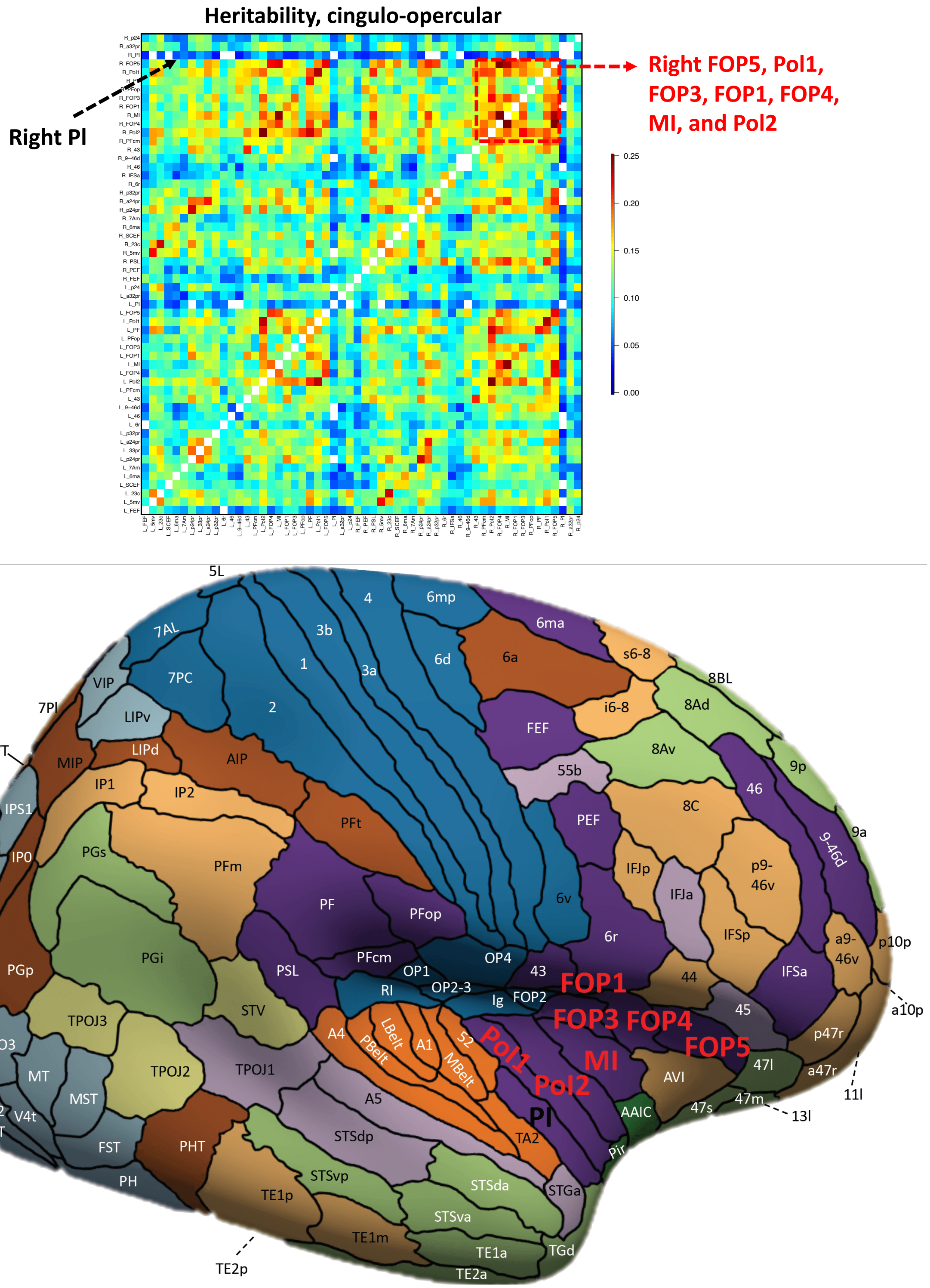


**Fig. S11 Heritability of the cingulo-opercular network in resting-state fMRI.**

Top: significant heritability estimates of the cingulo-opercular network in resting-state fMRI. Bottom: location of the highlighted areas that had the highest (in red) and lowest (in black) heritability (right hemisphere). Background colors indicate different functional networks, whose names can be found in Figure S1.

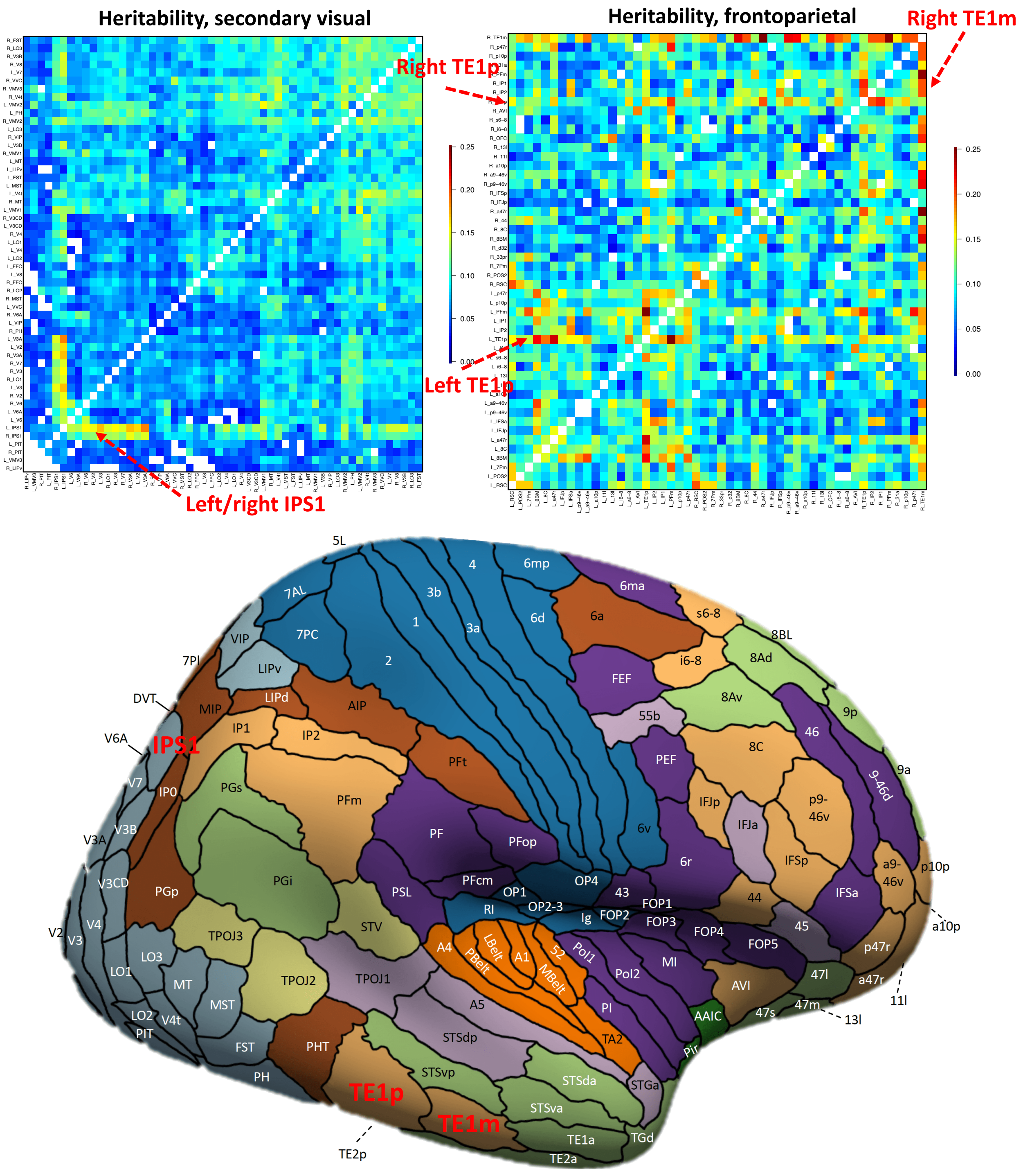


**Fig. S12 Heritability of the secondary visual and frontoparietal networks in resting-state fMRI.**

Top left: significant heritability estimates of the secondary visual network in resting-state fMRI. Top right: significant heritability estimates of the frontoparietal network in resting-state fMRI. Bottom: location of the highlighted areas that had the highest heritability (right hemisphere). Background colors indicate different functional networks, whose names can be found in Figure S1.

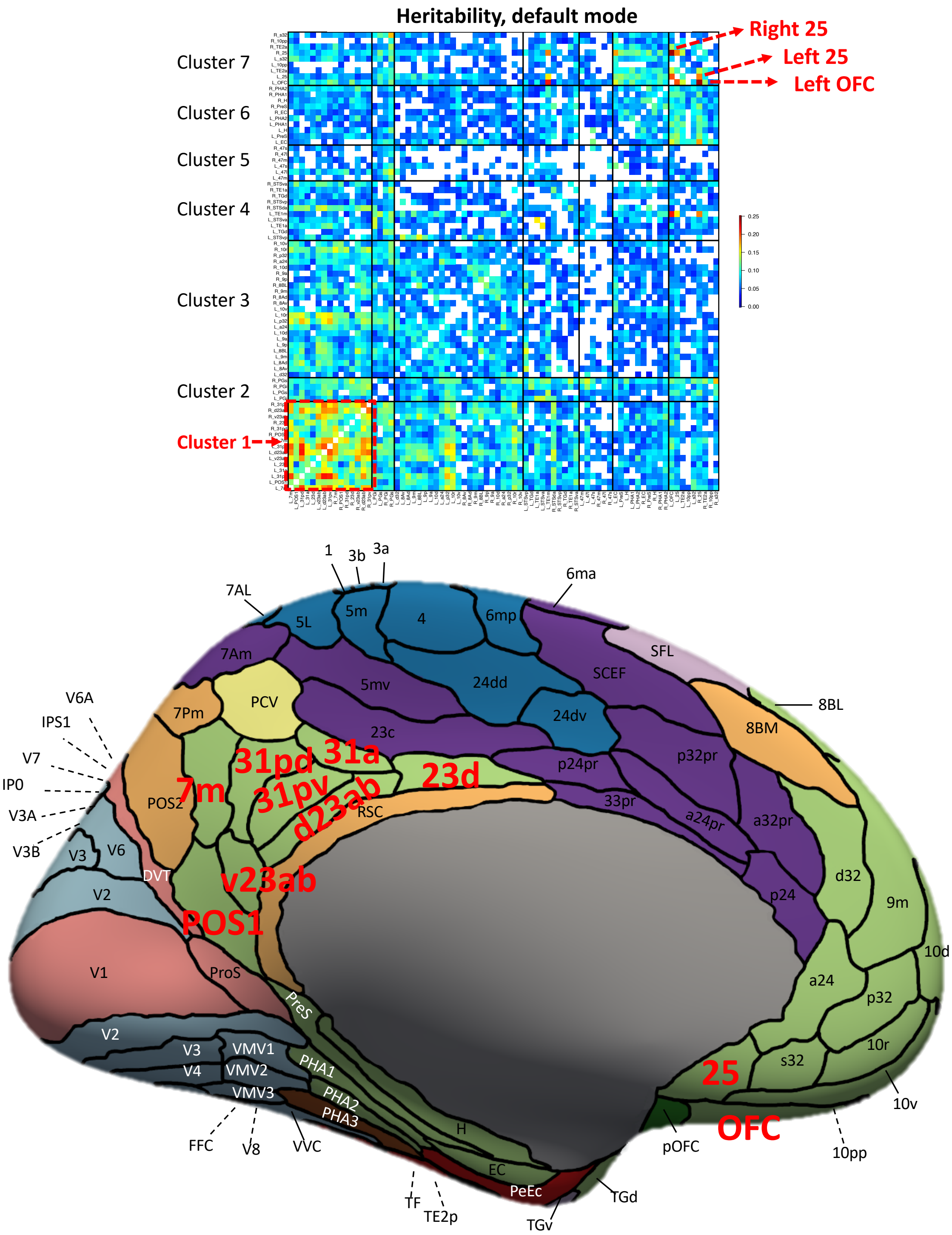


**Fig. S13 Heritability of the default mode network in task-evoked fMRI.**

Top: significant heritability estimates of the default mode network in task-evoked fMRI. The seven clusters in the default mode network can be found in Figure S9. Bottom: location of the highlighted areas that had the highest heritability (left hemisphere). Background colors indicate different functional networks, whose names can be found in Figure S1.

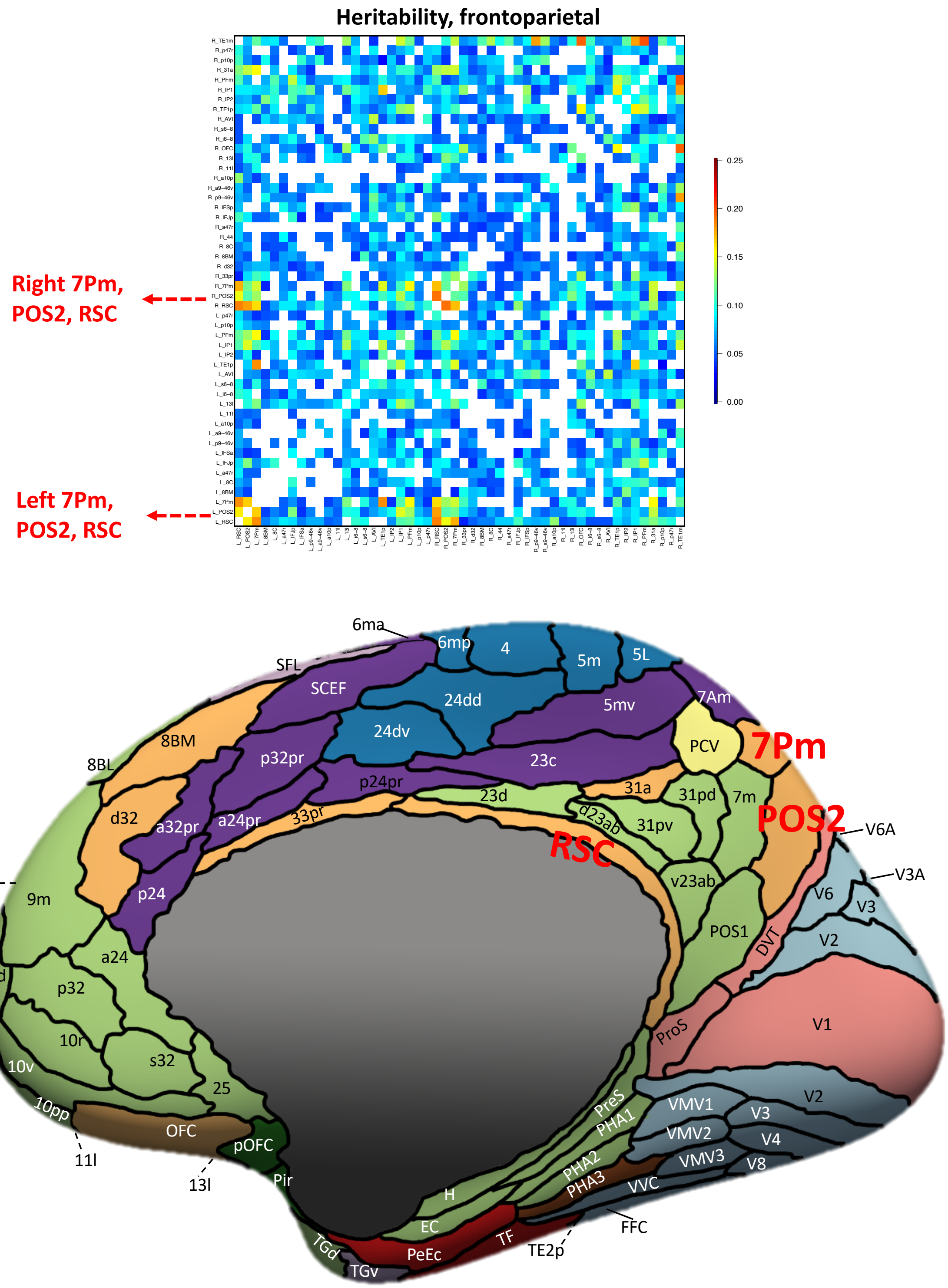


**Fig. S14 Heritability of the frontoparietal network in task-evoked fMRI.**

Top: significant heritability estimates of the frontoparietal network in task-evoked fMRI. Bottom: location of the highlighted areas that had the highest heritability (right hemisphere). Background colors indicate different functional networks, whose names can be found in Figure S1.

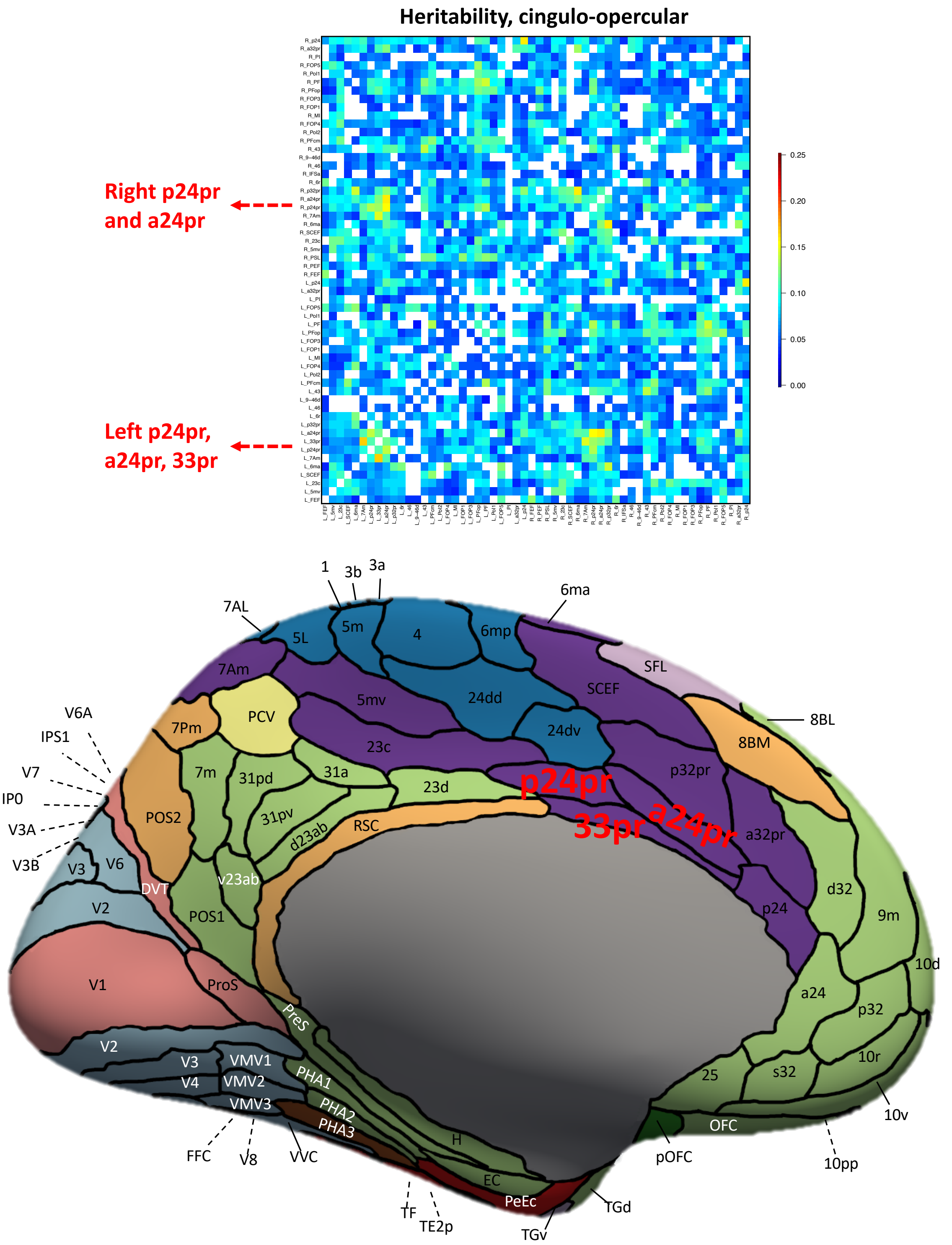


**Fig. S15 Heritability of the cingulo-opercular network in task-evoked fMRI.**

Top: significant heritability estimates of the cingulo-opercular network in task-evoked fMRI. Bottom: location of the highlighted areas that had the highest heritability (right hemisphere). Background colors indicate different functional networks, whose names can be found in Figure S1.

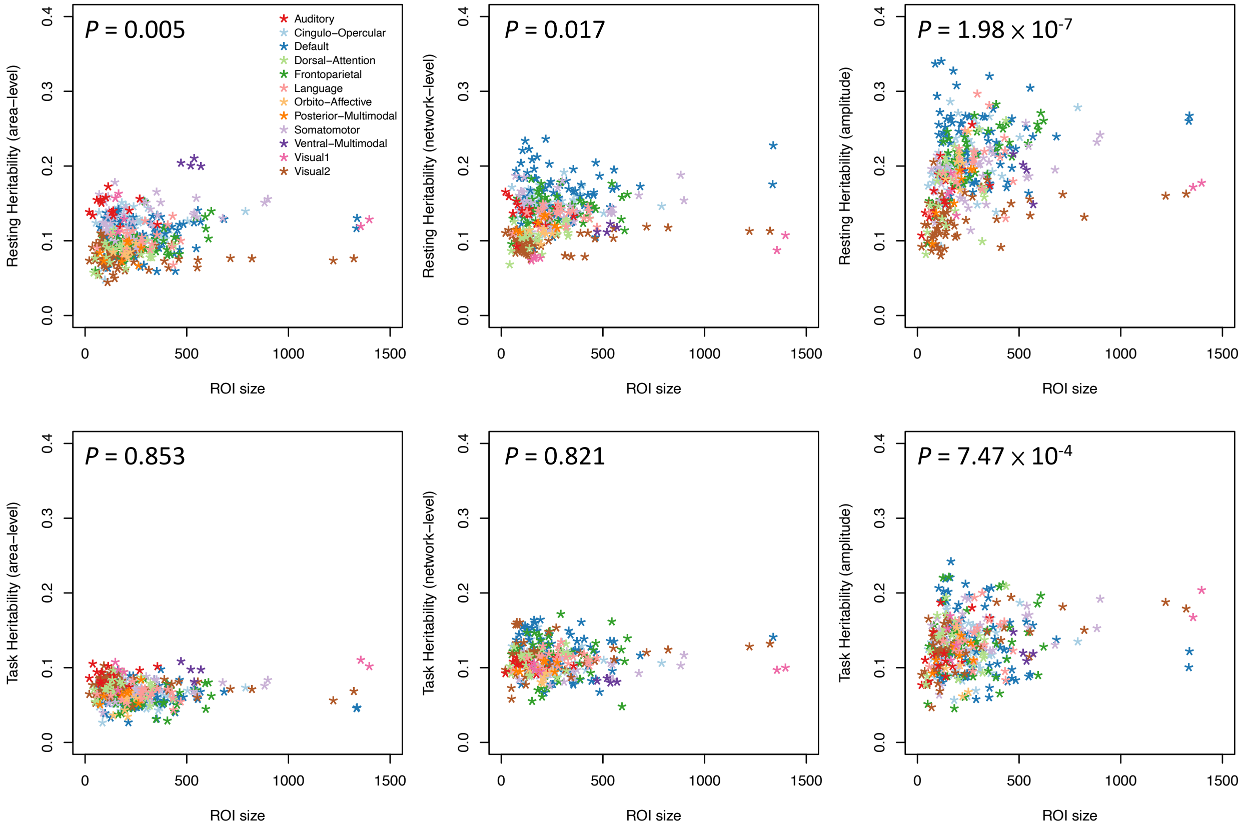


**Fig. S16 Relationship between mean heritability estimates and the size of cortical areas.**

From left to right, the three top panels illustrate the relationships for area-level traits, network-level traits, and amplitude traits in resting fMRI, respectively. The three bottom panels illustrate the maps for task fMRI. We labeled the *P*-values from the t-test in linear models.

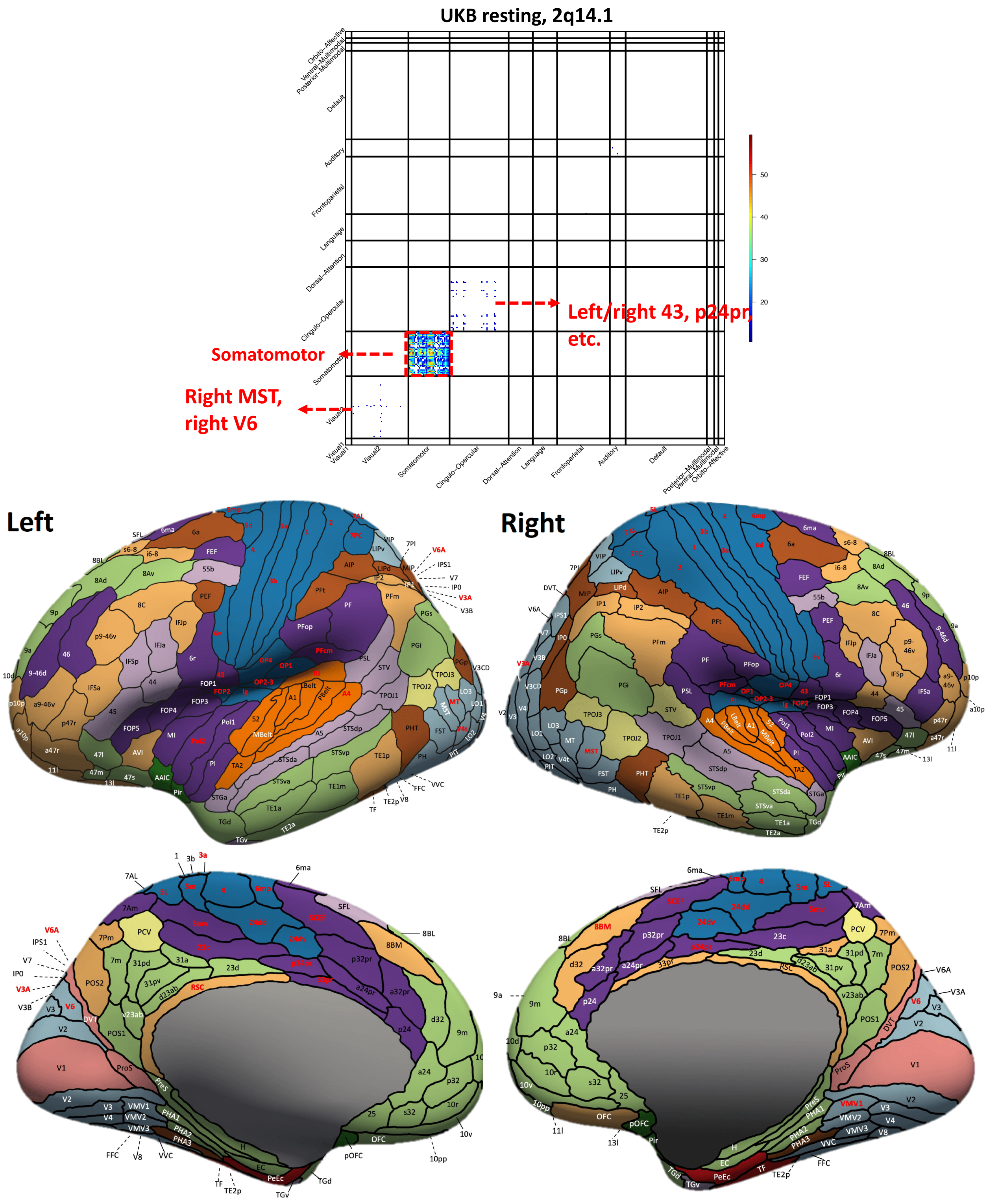


**Fig. S17 Functional areas and networks associated with 2q14.1 in resting-state fMRI.**

Top: map of -$\text{Log}_{10}$(Pvalue) for significant associations ($P < 2.93 \times 10^{-12}$). Bottom: location of the functional areas that having significant associations (names highlighted in red). Background colors indicate different functional networks, whose names can be found in Figure S1.

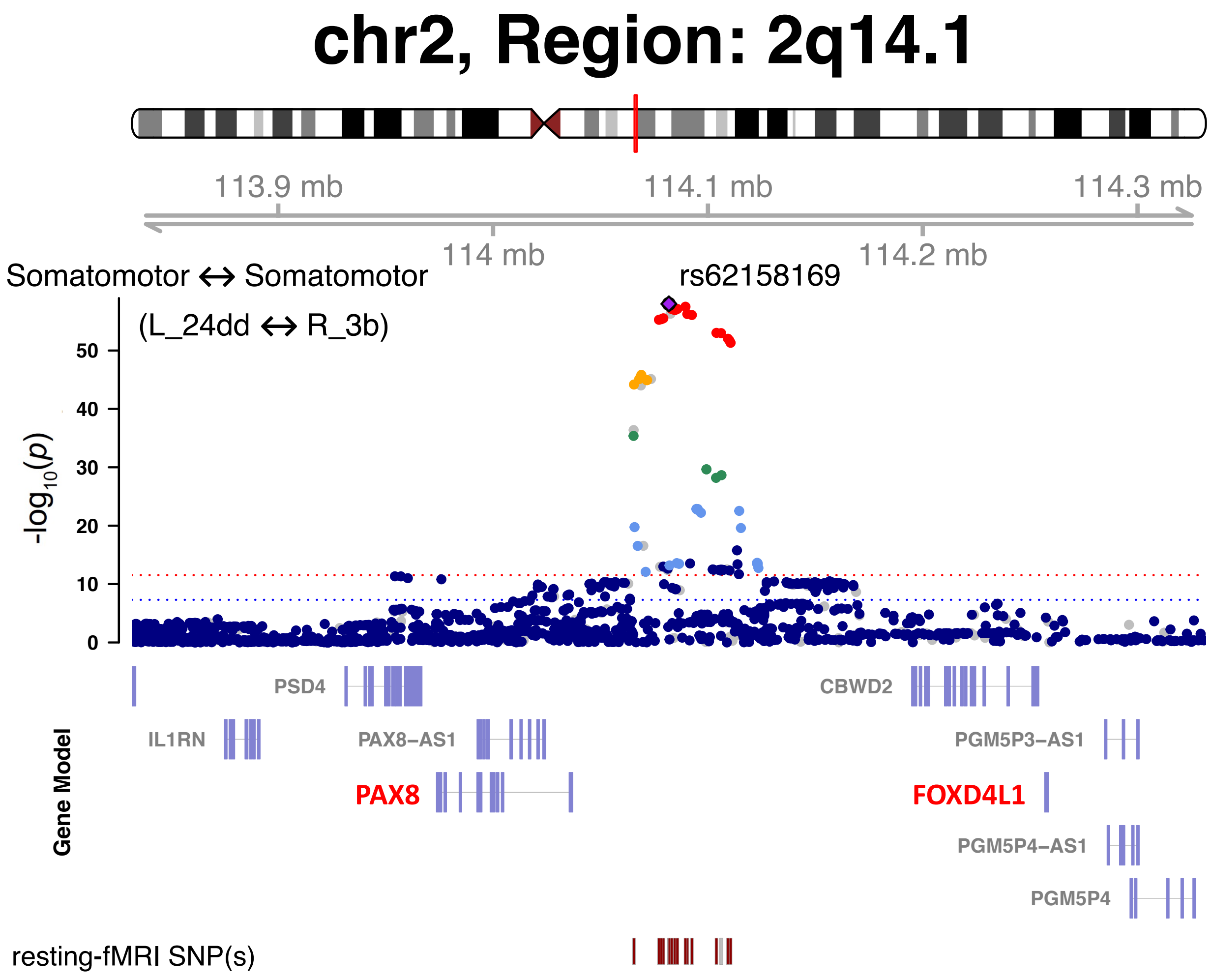


**Fig. S18 Selected functional areas and networks associated with 2q14.1 in resting-state fMRI.**

We illustrate one functional connectivity trait (between the L_24dd and R_3b areas within the somatomotor network) that is associated with the 2q14.1 locus (index variant rs62158160). The index variant and its proxy variants (LD $r^2 \geq 0.8$) are known brain eQTLs (https://metabrain.nl/) for the genes *PAX8* and *FOXD4L1*(bolded in red).

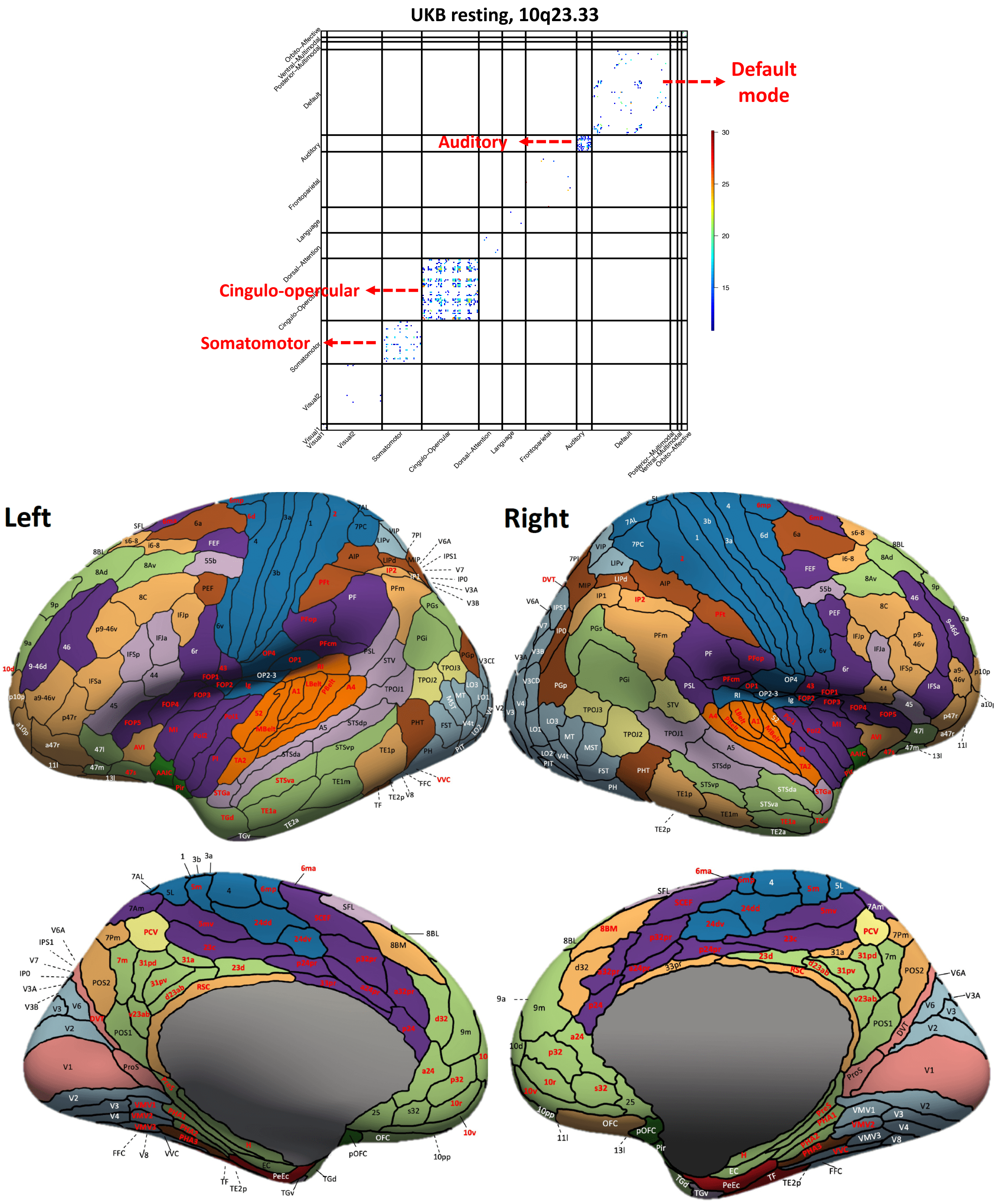


**Fig. S19 Functional areas and networks associated with 10q23.33 in resting-state fMRI.**

Top: map of -Log$_{10}$(Pvalue) for significant associations ($P < 2.93 \times 10^{-12}$). Bottom: location of the functional areas that having significant associations (names highlighted in red). Background colors indicate different functional networks, whose names can be found in Figure S1.

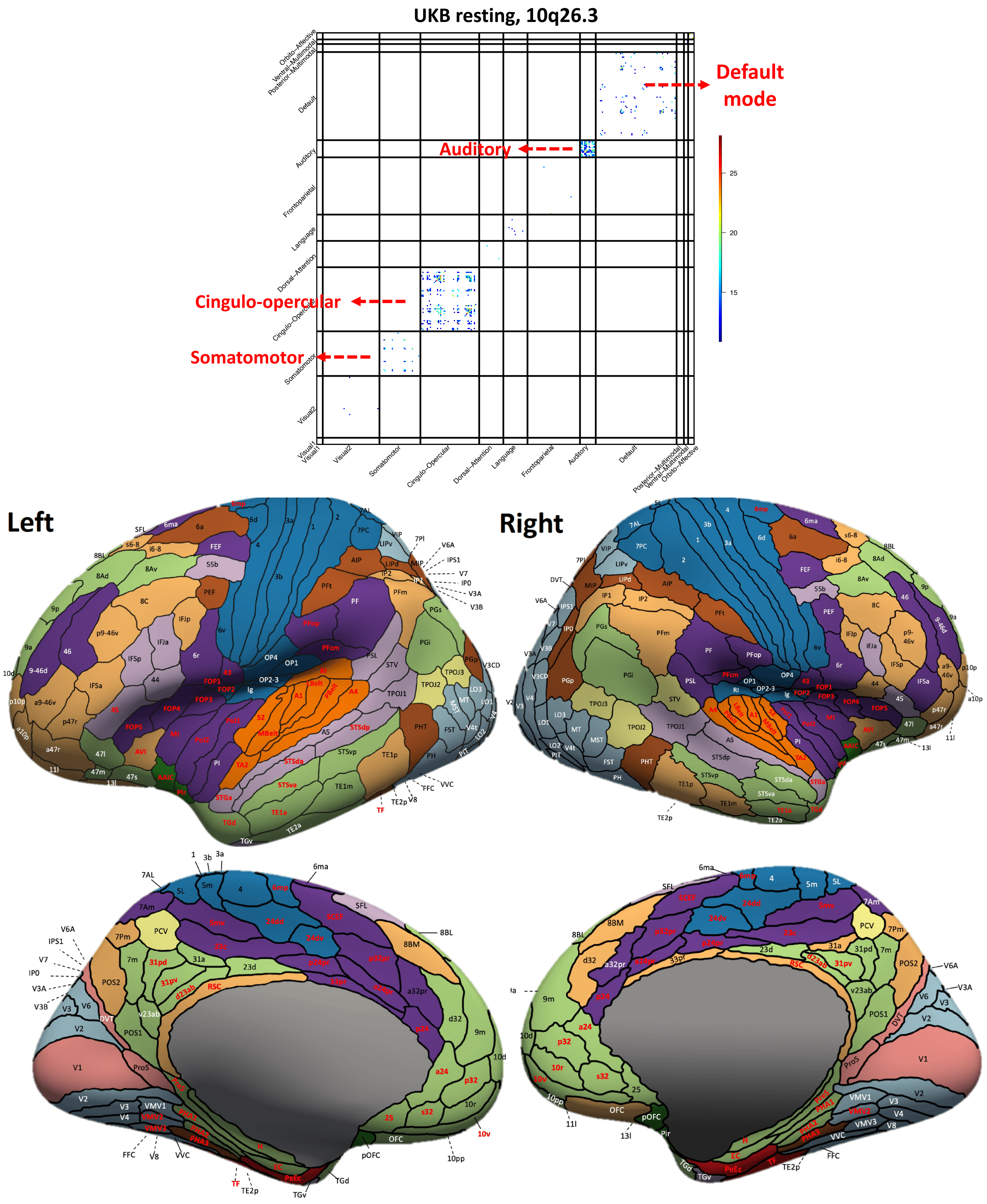


**Fig. S20 Functional areas and networks associated with 10q26.3 in resting-state fMRI.**

Top: map of -Log$_{10}$(Pvalue) for significant associations ($P < 2.93 \times 10^{-12}$). Bottom: location of the functional areas that having significant associations (names highlighted in red). Background colors indicate different functional networks, whose names can be found in Figure S1.

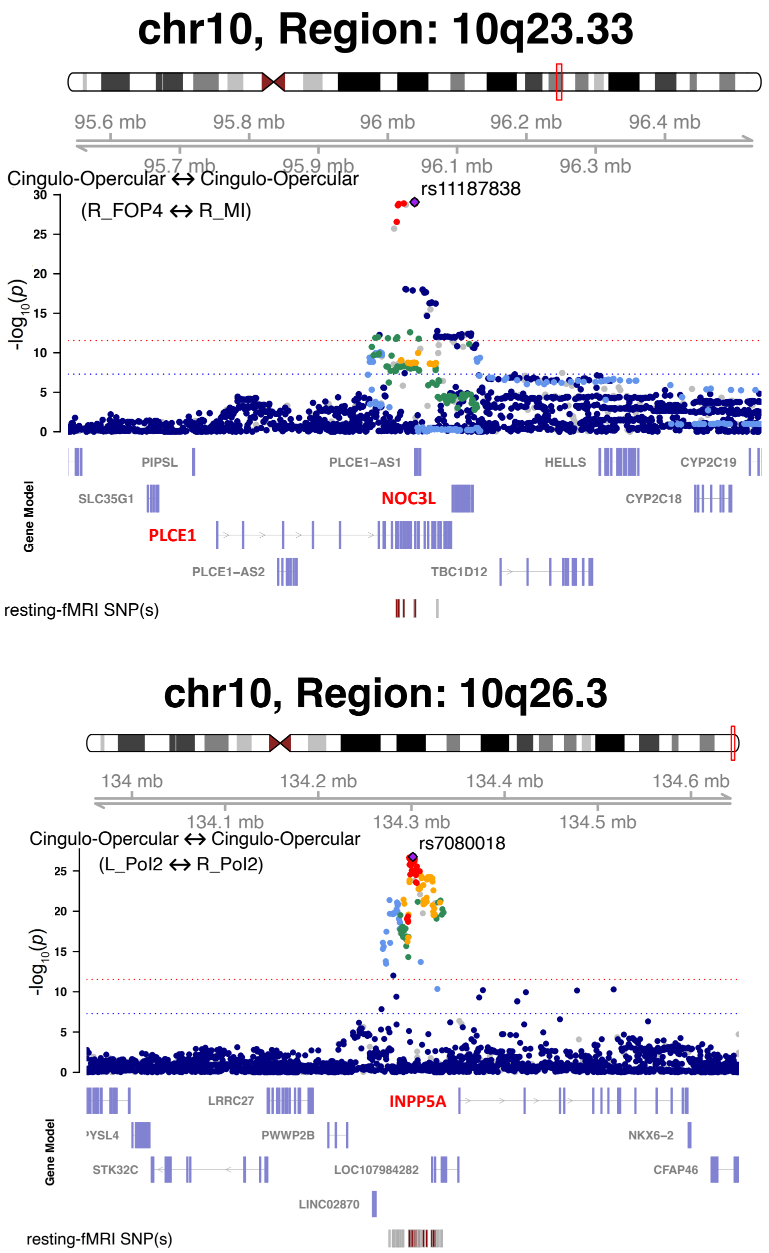


**Fig. S21 Selected functional areas and networks associated with 10q23.33 and 10q26.3 in resting-state fMRI.**

We illustrate one functional connectivity trait (top, between the R_FOP4 and R_MI areas within the cingulo-opercular network) associated with the 10q23.33 locus (index variant rs11187838) and one functional connectivity trait (bottom, between the L_Pol2 and R_Pol2 areas within the cingulo-opercular network) associated with the 10q26.3 locus (index variant rs7080018). The index variant and its proxy variants (LD $r^2 \geq 0.8$) are known brain eQTLs (https://metabrain.nl/) for the genes *NOC3L, PLCE1,* and *INPP5A* (bolded in red).

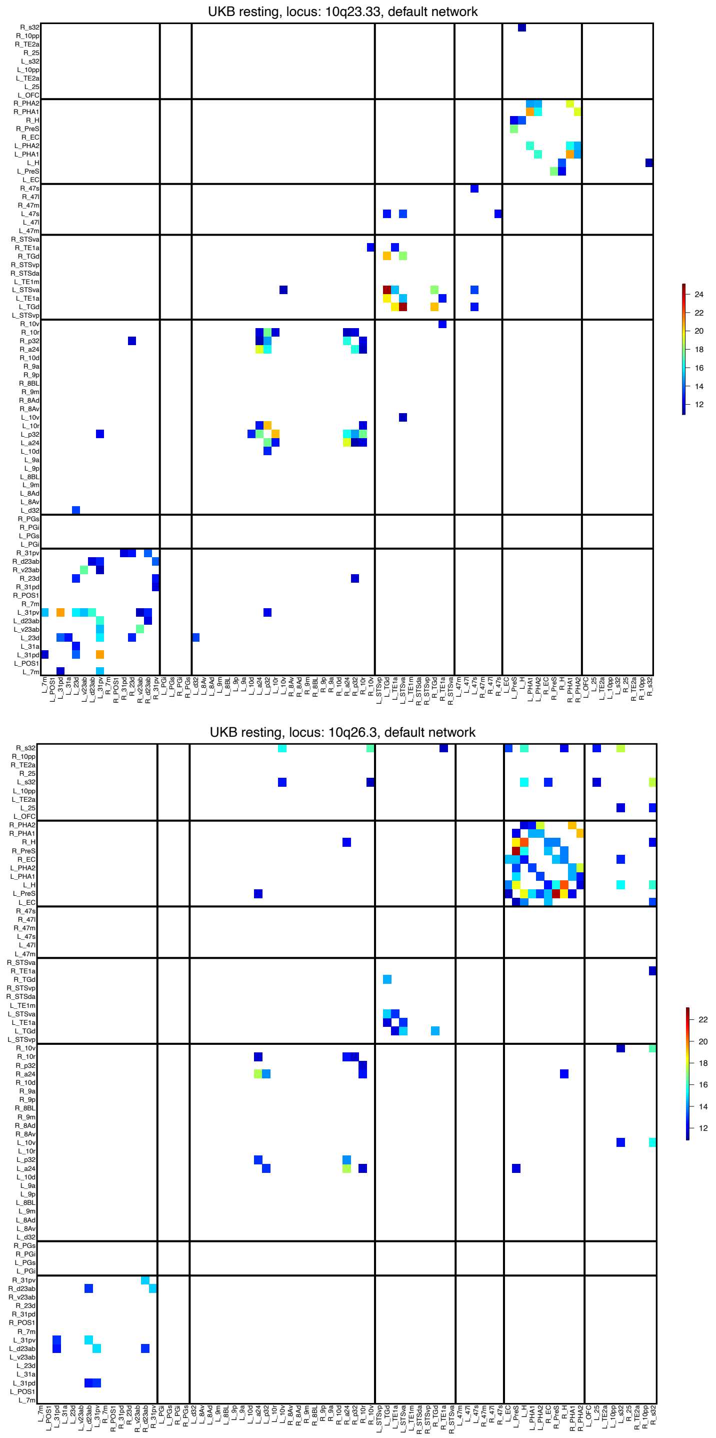


**Fig. S22 Functional areas associated with 10q23.33 and 10q26.3 in resting-state fMRI of the default mode network.**

We show the -Log$_{10}$(Pvalue) map for significant associations of 10q23.33 (top) and 10q26.3 (bottom) ($P < 2.93 \times 10^{-12}$). The location of the functional areas and subclusters of the default mode network can be found in Figure S9.

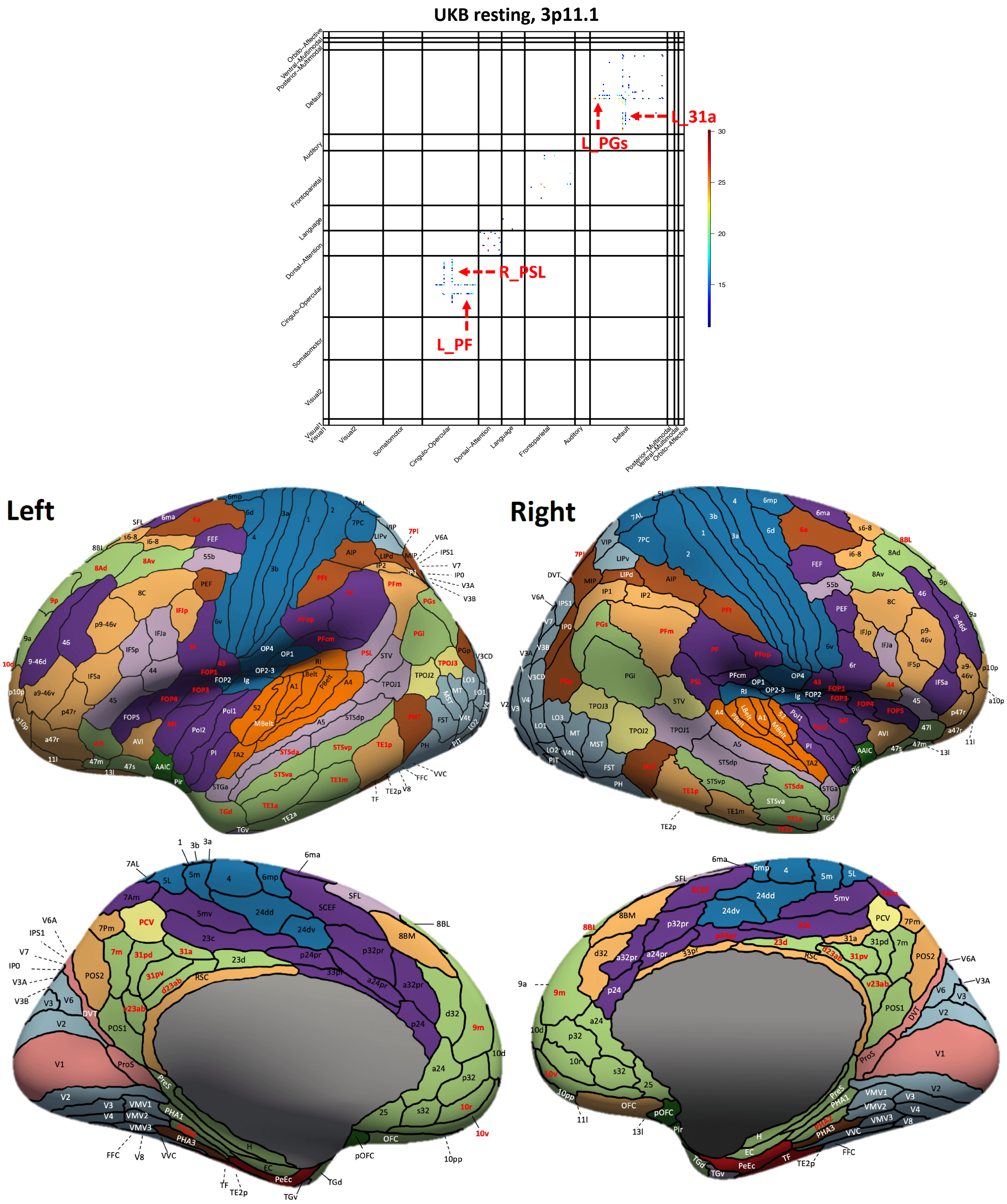


**Fig. S23 Functional areas and networks associated with 3p11.1 in resting-state fMRI.**

Top: map of -Log$_{10}$(Pvalue) for significant associations ($P$ < 2.93 × 10$^{-12}$). Bottom: location of the functional areas that having significant associations (names highlighted in red). Background colors indicate different functional networks, whose names can be found in Figure S1.

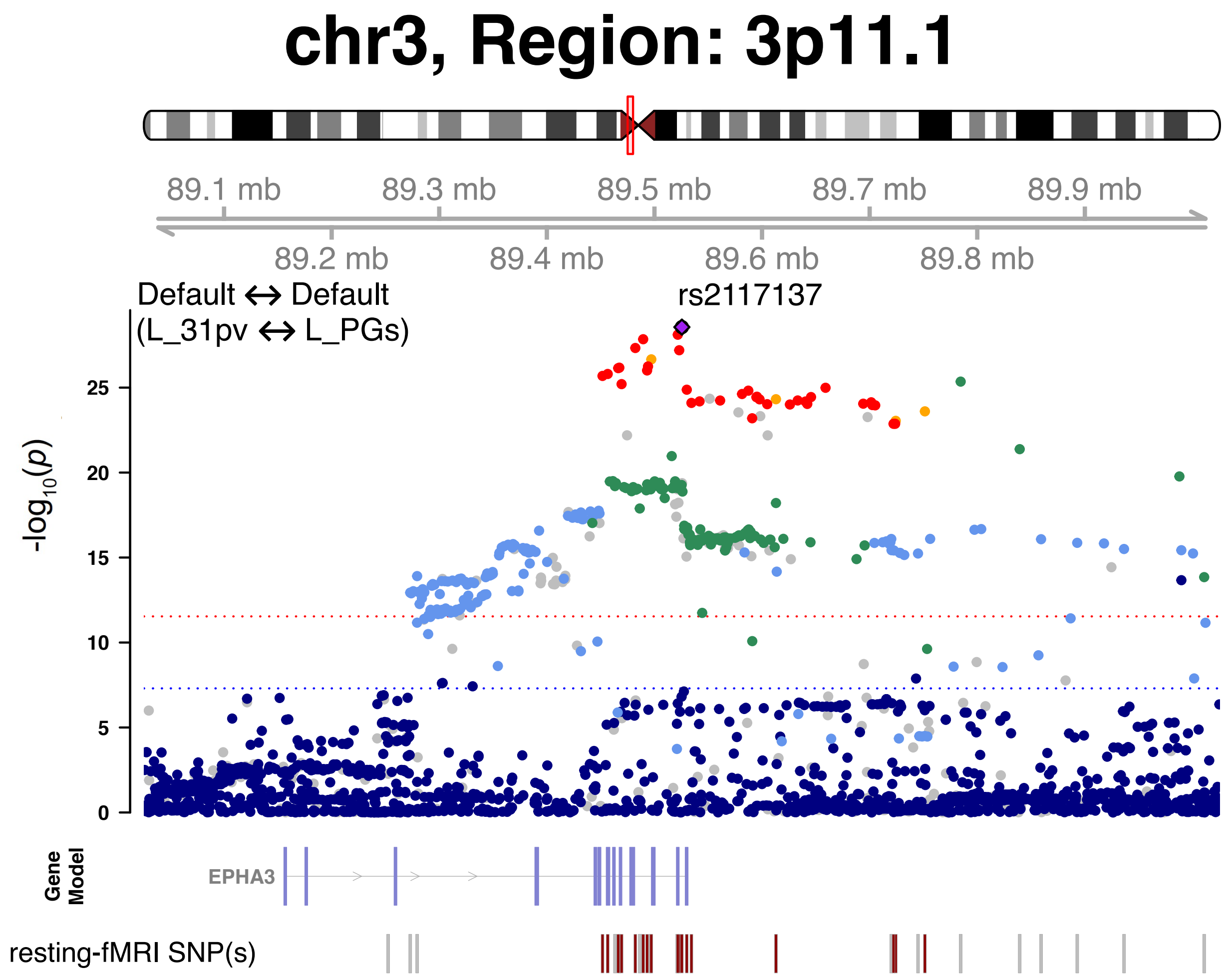


**Fig. S24 Selected functional areas and networks associated with 3p11.1 in resting-state fMRI.**

We illustrate one functional connectivity trait (between the L_31pv and L_PGs areas within the default mode network) associated with the 3p11.1 locus (index variant rs2117137).

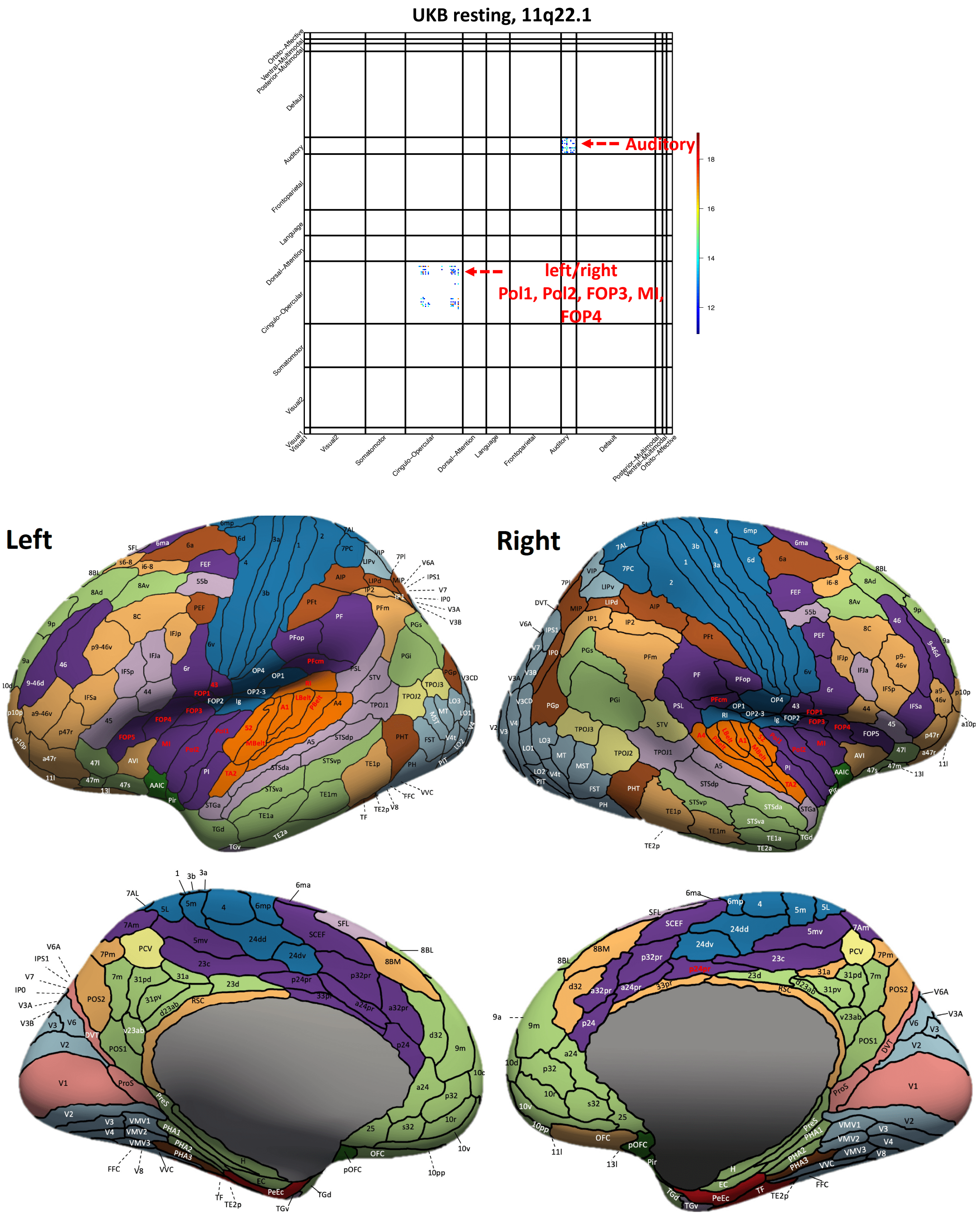


**Fig. S25 Functional areas and networks associated with 11q22.1 in resting-state fMRI.**

Top: map of -Log$_{10}$(Pvalue) for significant associations ($P < 2.93 \times 10^{-12}$). Bottom: location of the functional areas that having significant associations (names highlighted in red). Background colors indicate different functional networks, whose names can be found in Figure S1.

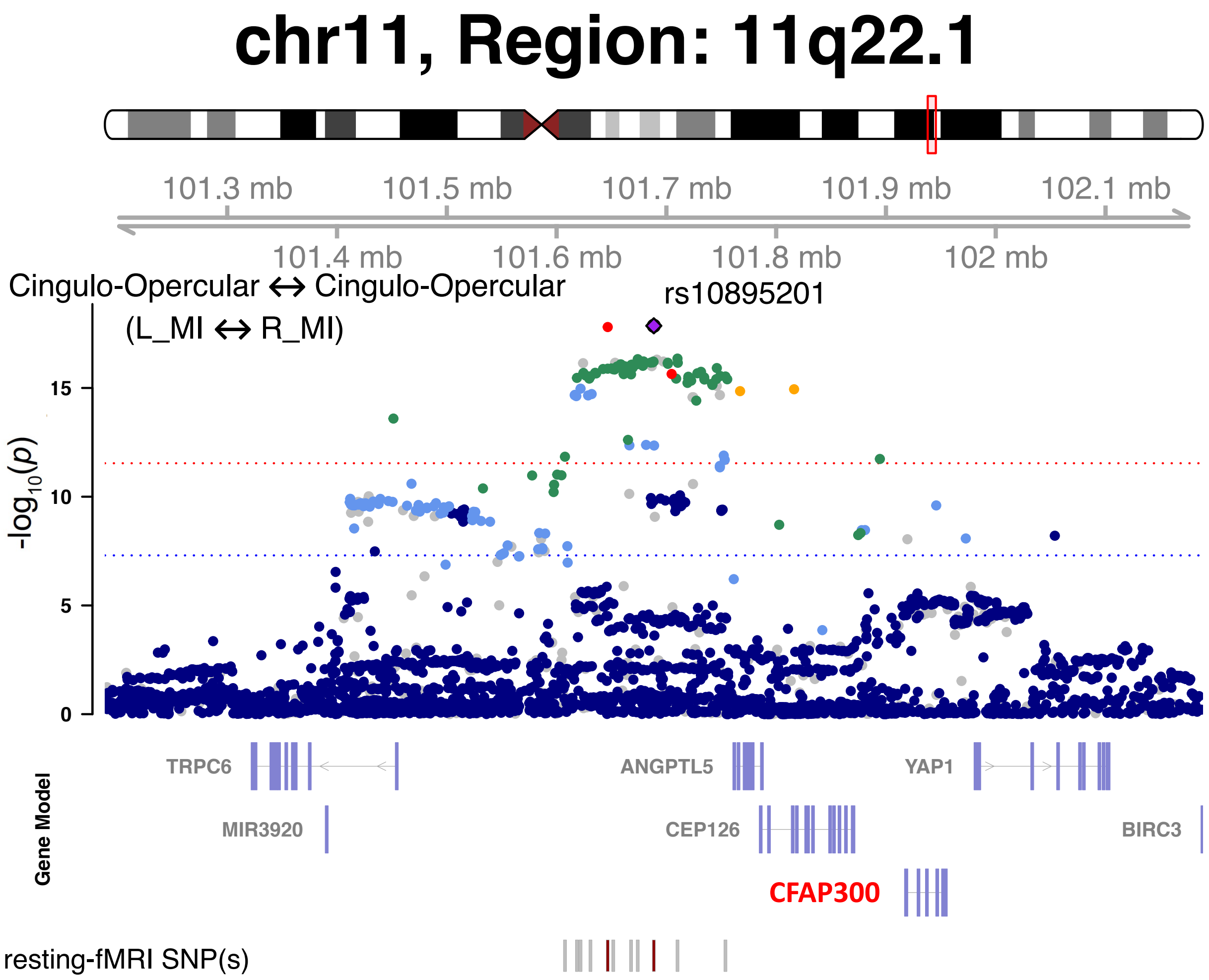


**Fig. S26 Selected functional areas and networks associated with 11q22.1 in resting-state fMRI.**

We illustrate one functional connectivity trait (between the L_MI and R_MI areas within the cingulo-opercular network) associated with the 11q22.1 locus (index variant rs10895201). The index variant and its proxy variants (LD $r^2 \geq 0.8$) are known brain eQTLs (https://metabrain.nl/) for the gene *CFAP300* (bolded in red).

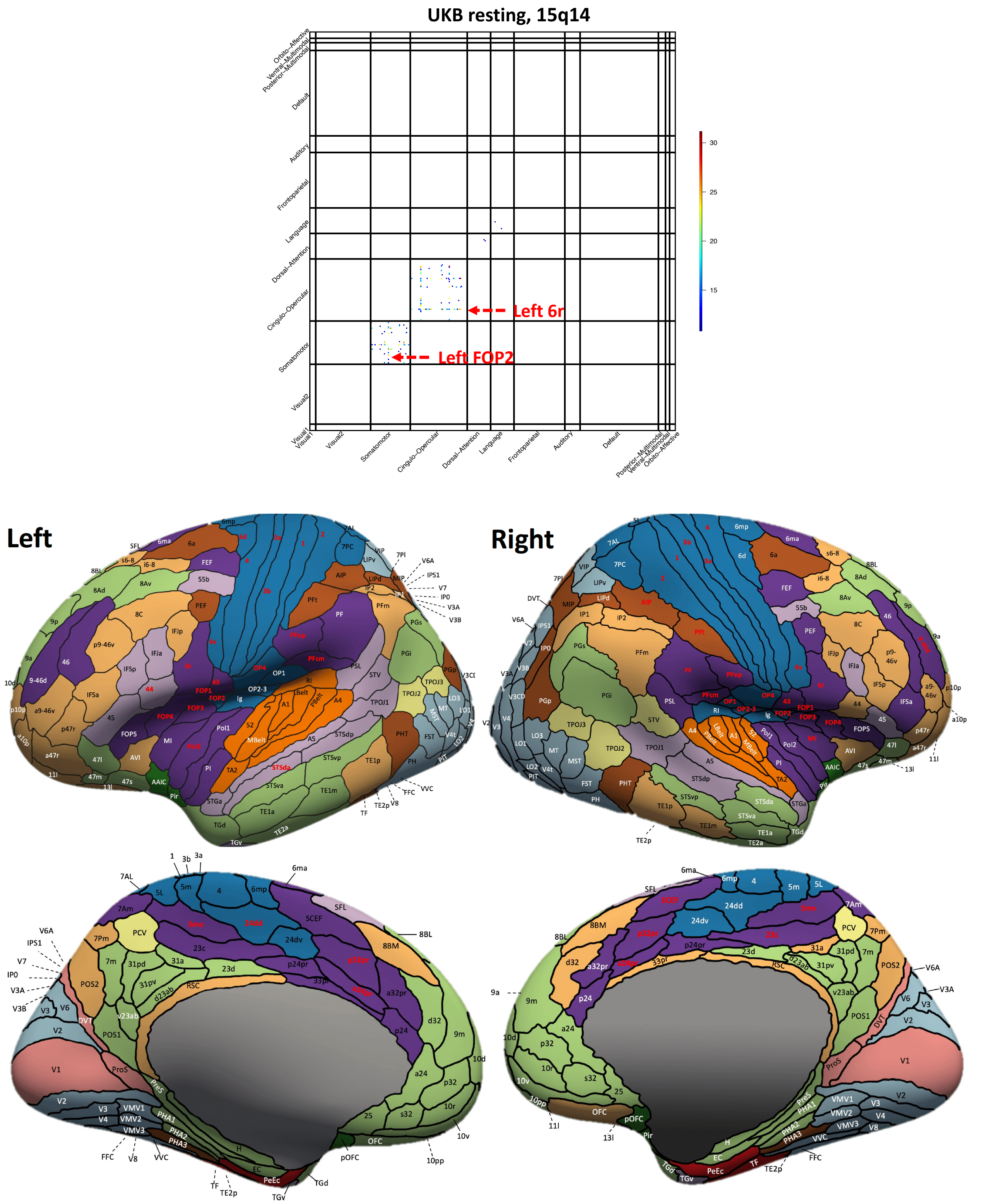


**Fig. S27 Functional areas and networks associated with 15q14 in resting-state fMRI.**

Top: map of -Log10(Pvalue) for significant associations ($P < 2.93 \times 10^{-12}$). Bottom: location of the functional areas that having significant associations (names highlighted in red). Background colors indicate different functional networks, whose names can be found in Figure S1.

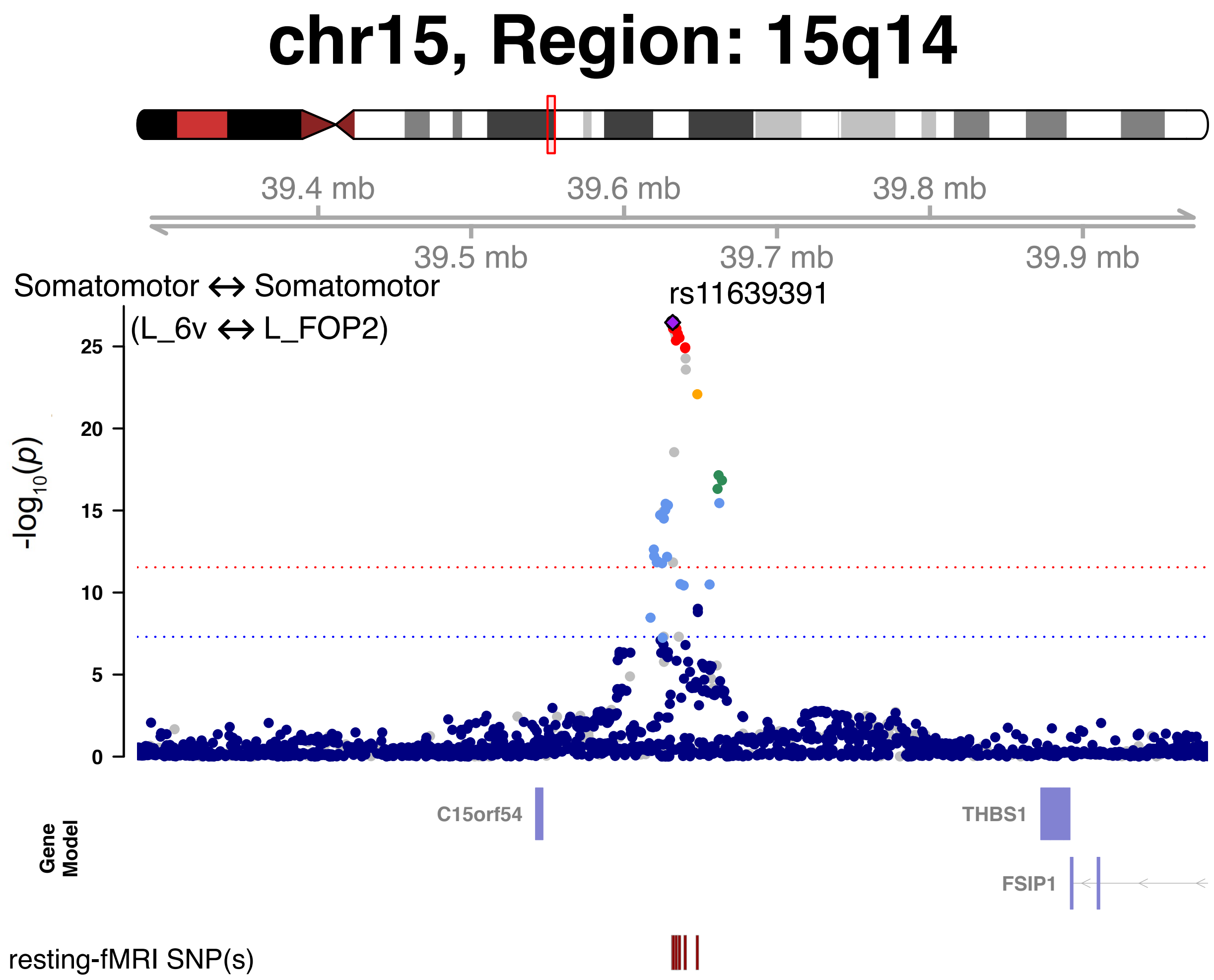


**Fig. S28 Selected functional areas and networks associated with 15q14 in resting-state fMRI.**

We illustrate one functional connectivity trait (between the L_6v and L_FOP2 areas within the somatomotor network) associated with the 15q14 locus (index variant rs11639391).

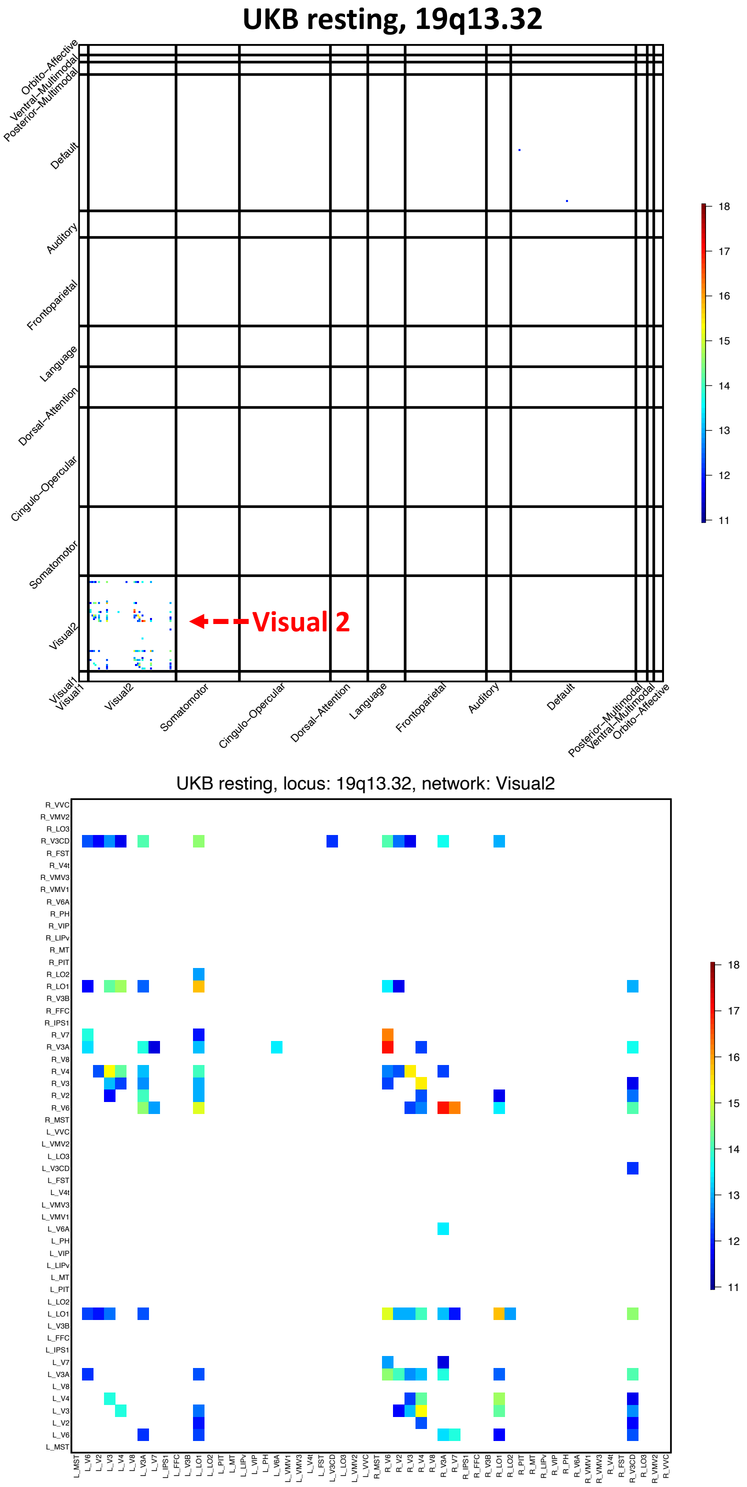


**Fig. S29 Functional areas and networks associated with 19q13.32 in resting-state fMRI.**

Top: map of -$Log_{10}$(Pvalue) for significant associations across all networks ($P < 2.93 \times 10^{-12}$). Bottom: map of -$Log_{10}$(Pvalue) for significant associations in the secondary visual network.

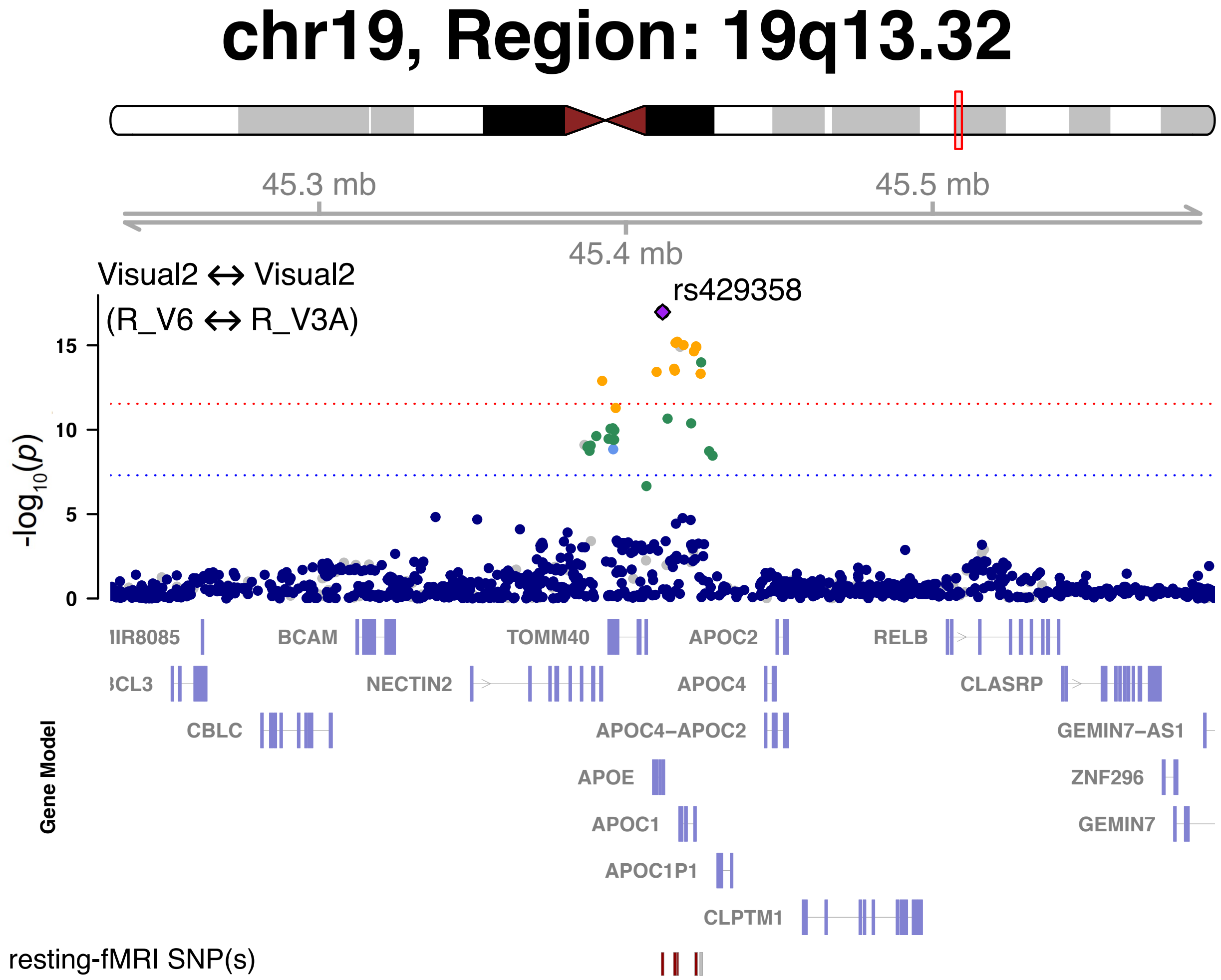


**Fig. S30 Selected functional areas and networks associated with 19q13.32 in resting-state fMRI.**

We illustrate one functional connectivity trait (between the R_6v and R_V3A areas within the secondary visual network) associated with the 19q13.32 locus (index variant rs429358).

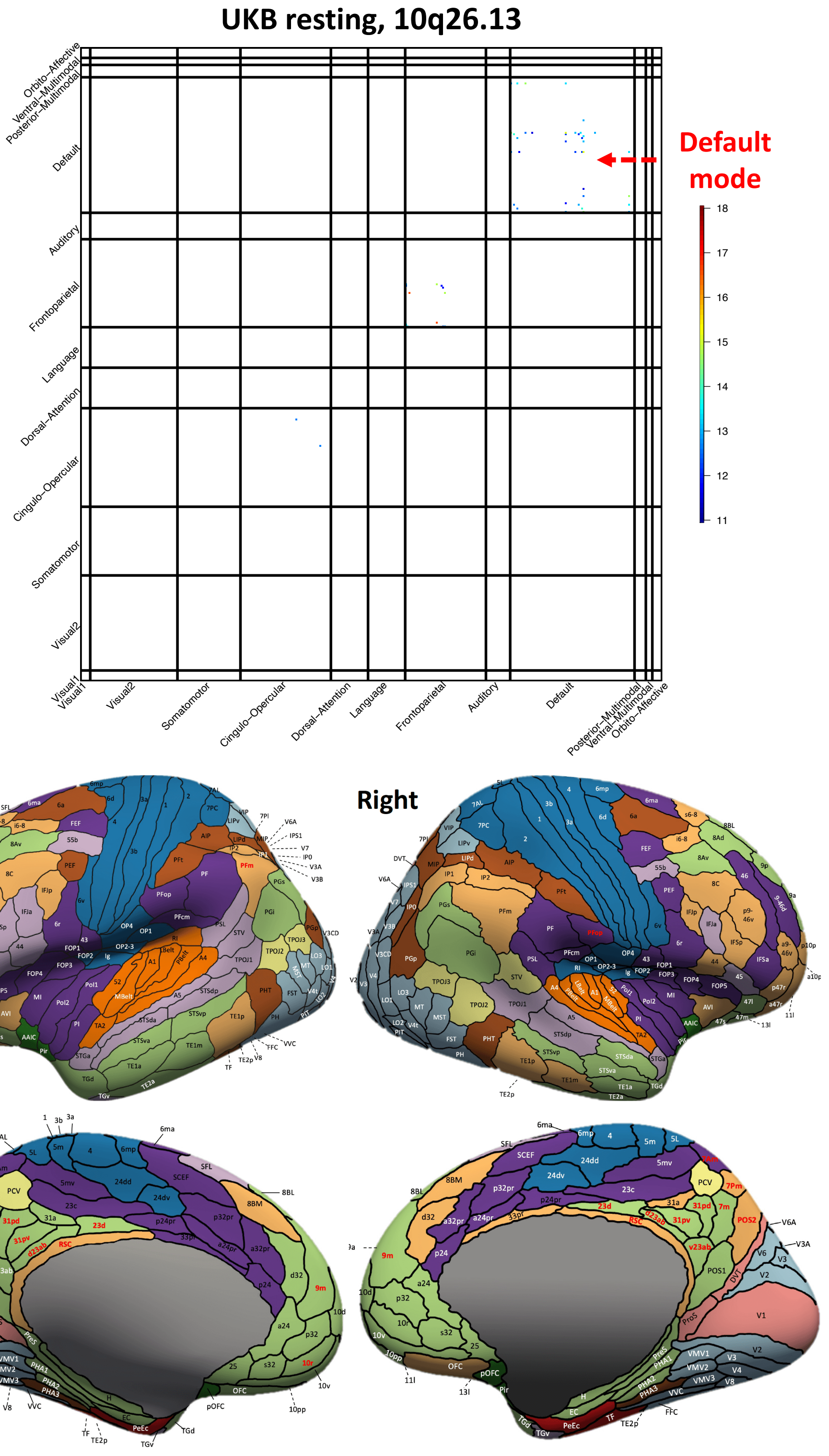


**Fig. S31 Functional areas and networks associated with 10q26.13 in resting-state fMRI.**

Top: map of -Log$_{10}$(Pvalue) for significant associations ($P < 2.93 \times 10^{-12}$). Bottom: location of the functional areas that having significant associations (names highlighted in red). Background colors indicate different functional networks, whose names can be found in Figure S1.

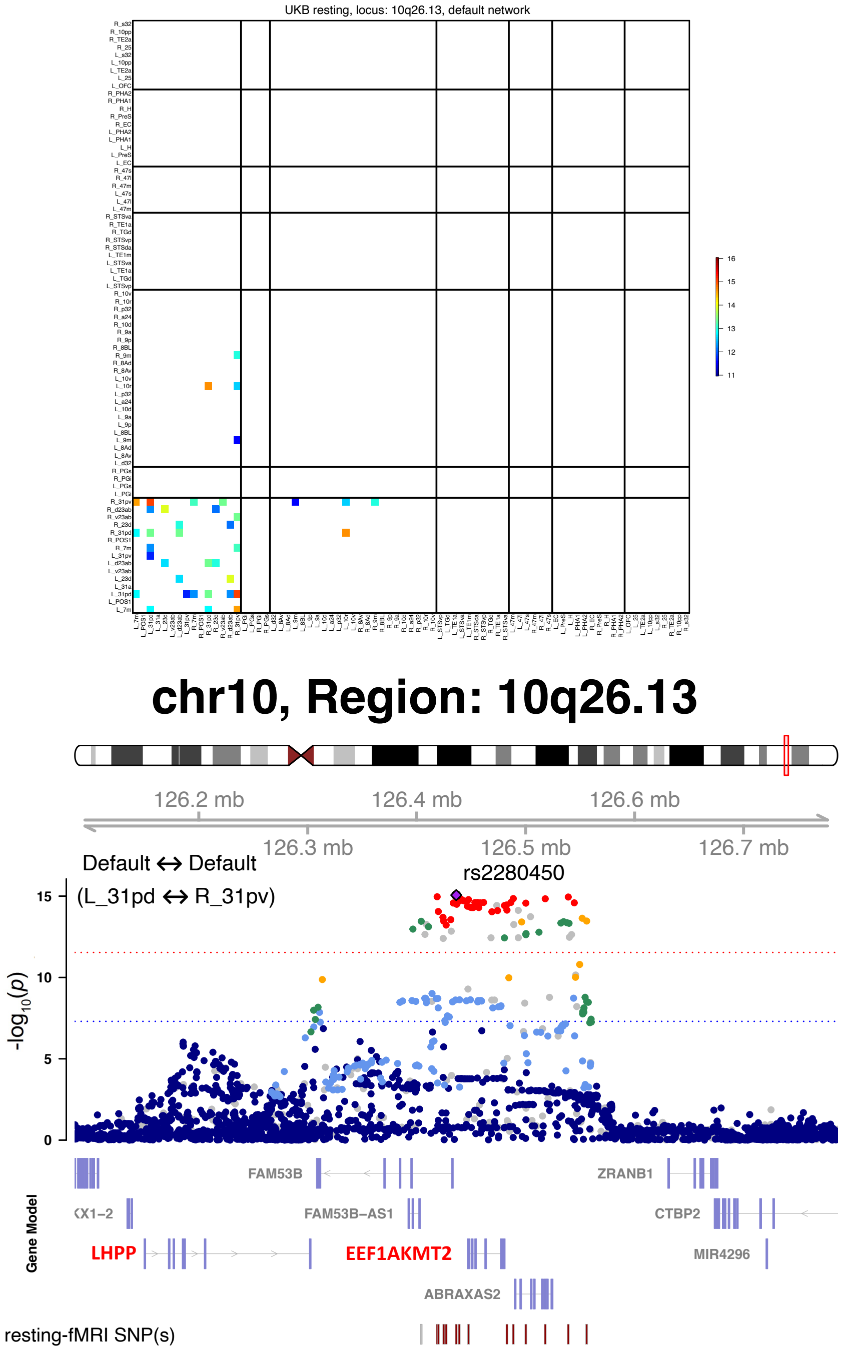


**Fig. S32 Functional areas associated with 10q26.13 in the default mode network of resting-state fMRI.**

Top: we show the -Log$_{10}$(Pvalue) map for significant associations of 10q26.13 ($P < 2.93 \times 10^{-12}$). Location of the functional areas and subclusters of default mode network can be found in Figure S11. Bottom: We illustrate one functional connectivity trait (between the L_31pd and R_31pv areas within the default mode network) associated with the 10q26.13 locus (index variant rs2280450). The index variant and its proxy variants (LD $r^2 \geq 0.8$) are known brain eQTLs ([https://metabrain.nl/](https://metabrain.nl/)) for the genes *LHPP* and *EEF1AKMT2* (bolded in red).

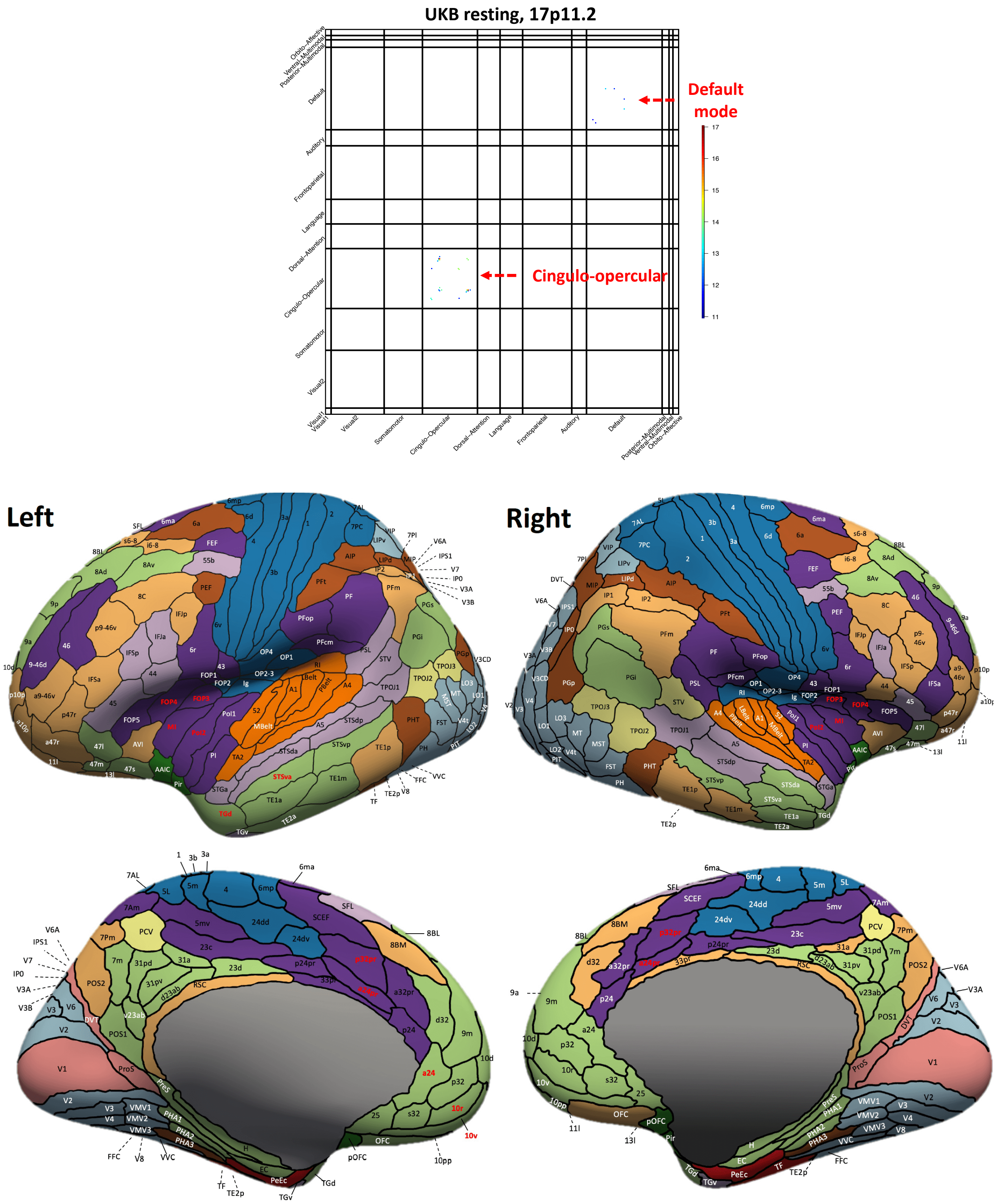


**Fig. S33 Functional areas and networks associated with 17p11.2 in resting-state fMRI.**

Top: map of -Log$_{10}$(Pvalue) for significant associations ($P < 2.93 \times 10^{-12}$). Bottom: location of the functional areas that having significant associations (names highlighted in red). Background colors indicate different functional networks, whose names can be found in Figure S1.

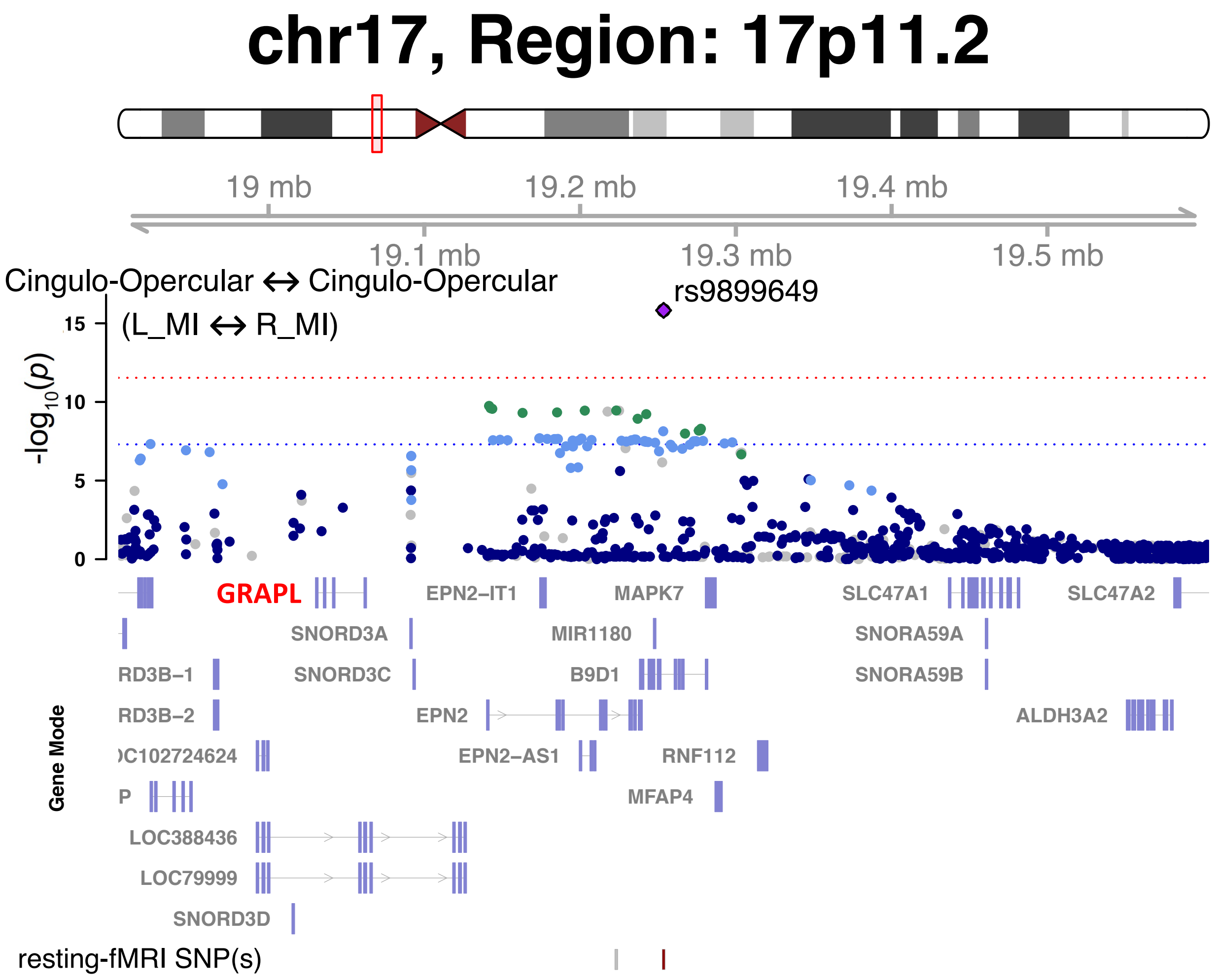


**Fig. S34 Selected Functional areas and networks associated with 17p11.2 in resting-state fMRI.**

We illustrate one functional connectivity trait (between the L_MI and R_MI areas within the cingulo-opercular network) associated with the 17p11.2 locus (index variant rs9899649). The index variant and its proxy variants (LD $r^2 \geq 0.8$) are known brain eQTLs (https://metabrain.nl/) for the gene *GRAPL* (bolded in red).

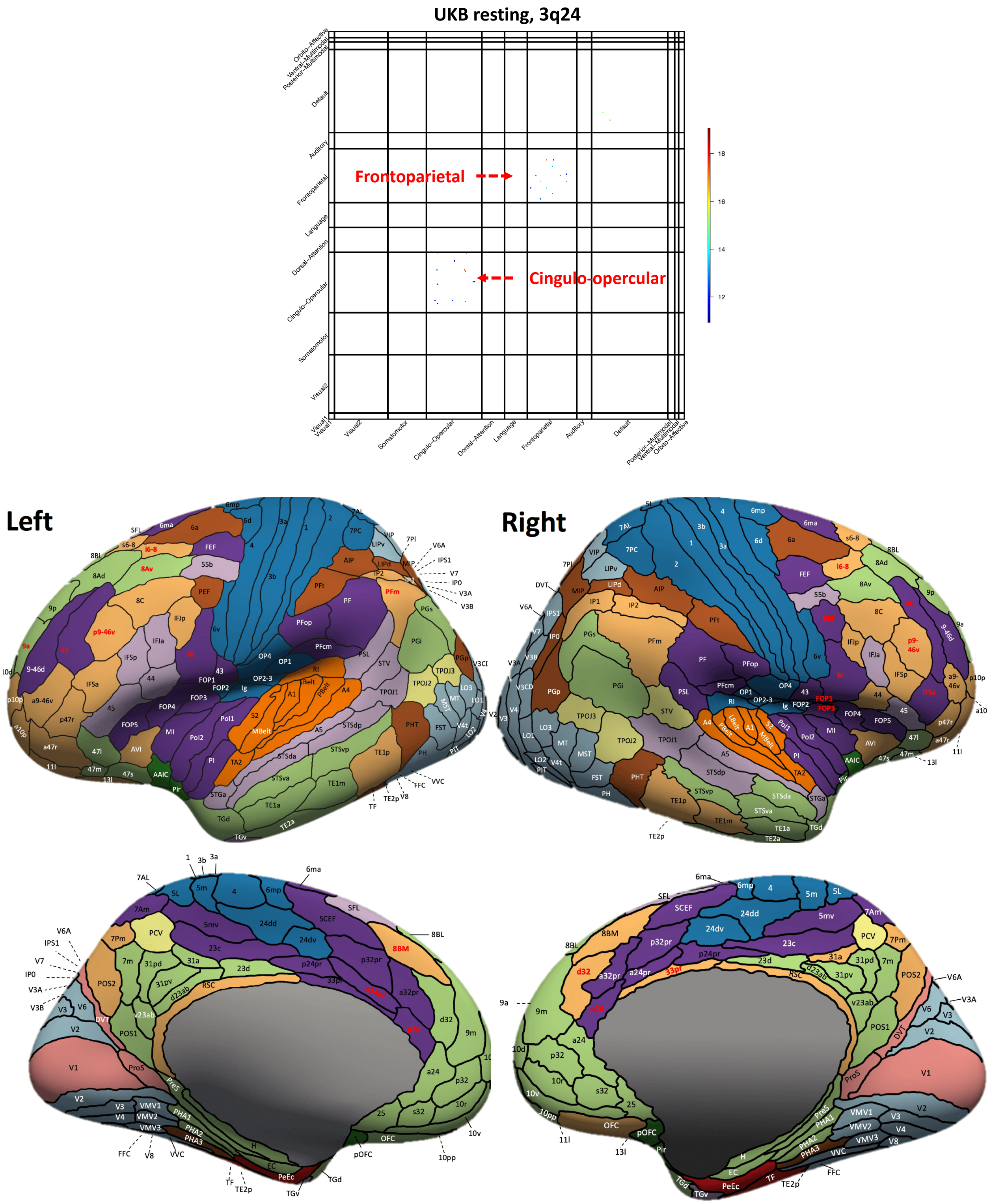


**Fig. S35 Functional areas and networks associated with 3q24 in resting-state fMRI.**

Top: map of -Log$_{10}$(Pvalue) for significant associations ($P < 2.93 \times 10^{-12}$). Bottom: location of the functional areas that having significant associations (names highlighted in red). Background colors indicate different functional networks, whose names can be found in Figure S1.

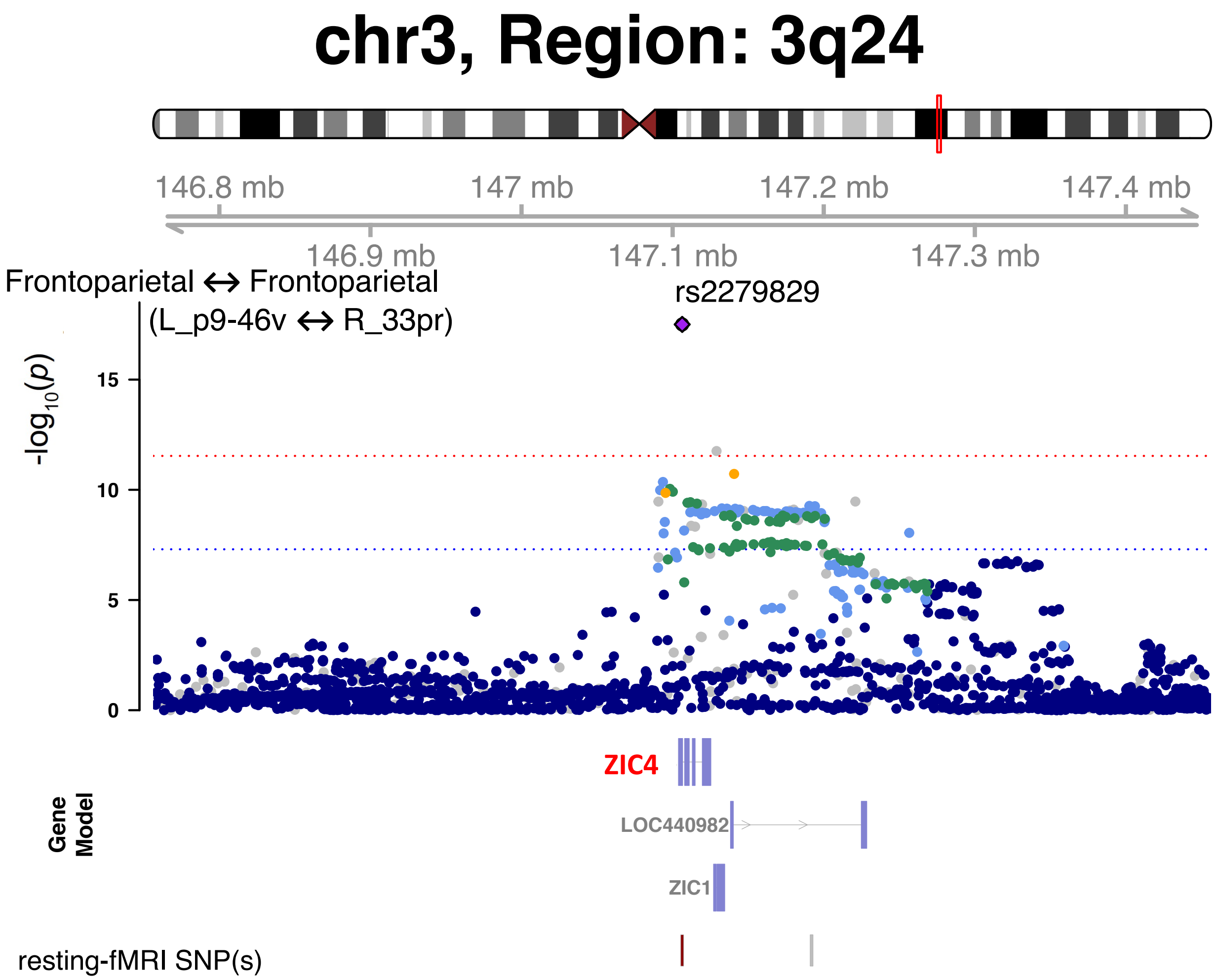


**Fig. S36 Selected functional areas and networks associated with 3q24 in resting-state fMRI.**

We illustrate one functional connectivity trait (between the L_p9-46v and R_33pr areas within the frontoparietal network) associated with the 3q24 locus (index variant rs2279829). The index variant and its proxy variants (LD $r^2 \geq 0.8$) are known brain eQTLs (https://metabrain.nl/) for the gene *ZIC4* (bolded in red).

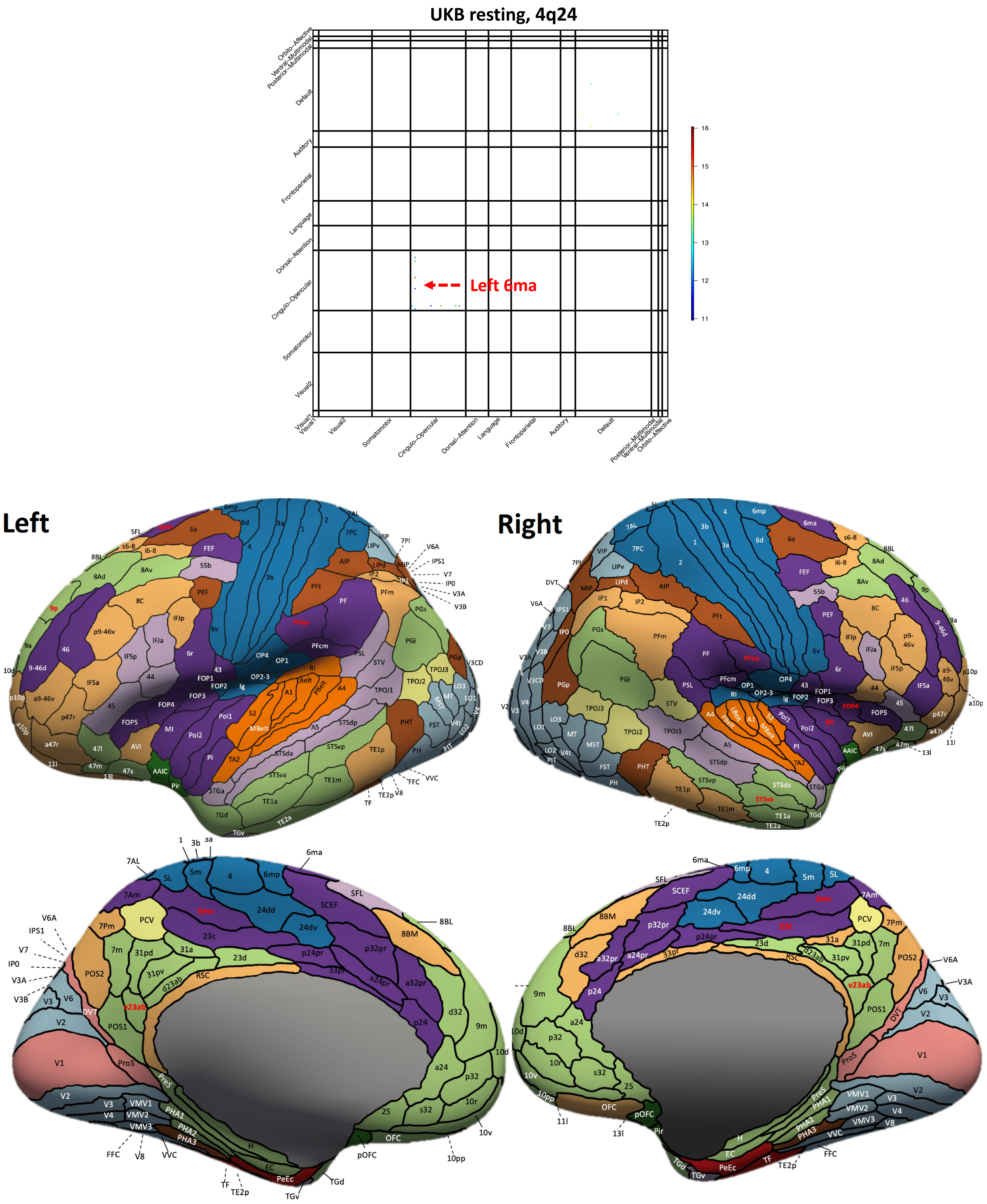


**Fig. S37 Functional areas and networks associated with 4q24 in resting-state fMRI.**

Top: map of -Log10(Pvalue) for significant associations ($P < 2.93 \times 10^{-12}$). Bottom: location of the functional areas that having significant associations (names highlighted in red). Background colors indicate different functional networks, whose names can be found in Figure S1.

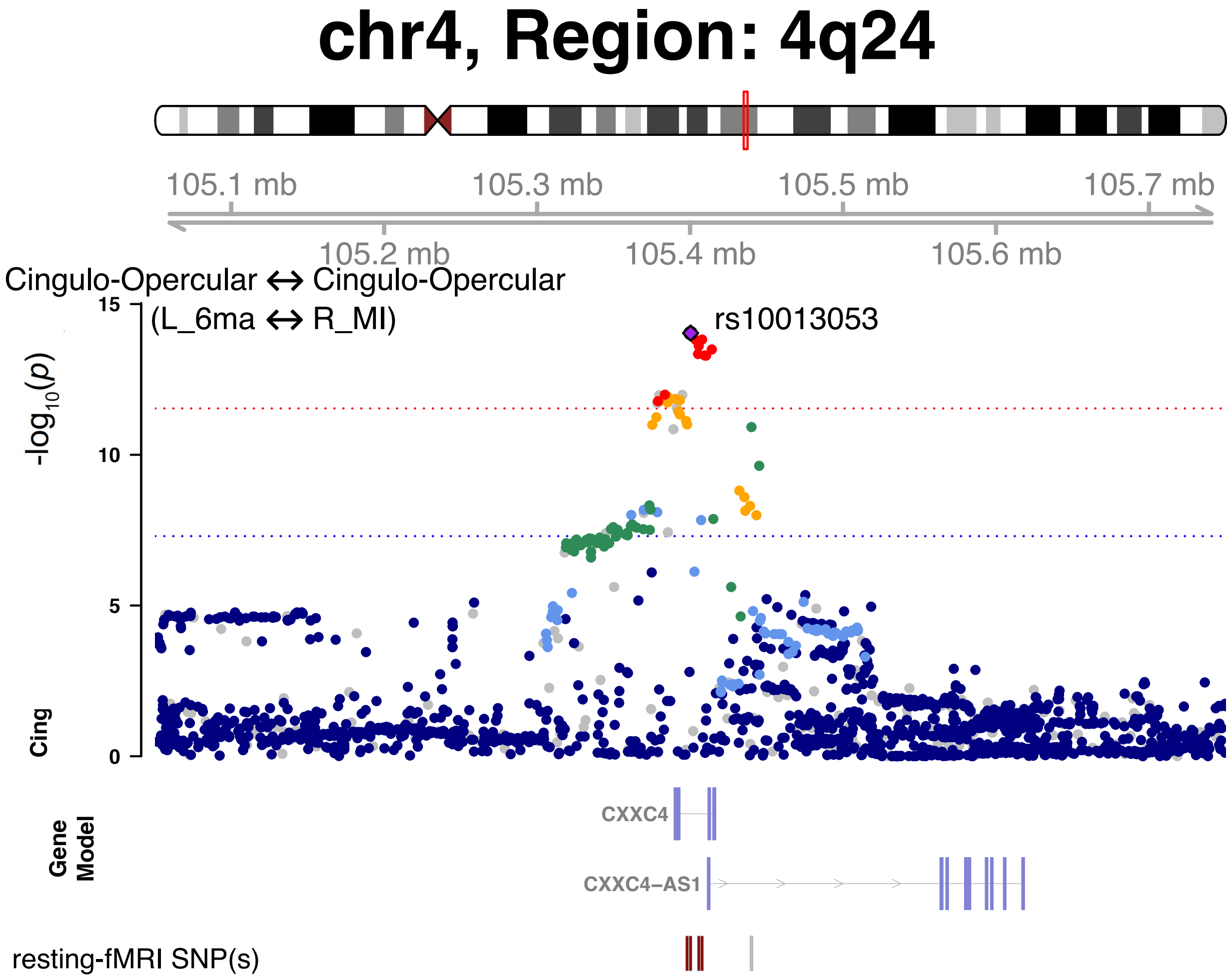


**Fig. S38 Selected functional areas and networks associated with 4q24 in resting-state fMRI.**

We illustrate one functional connectivity trait (between the L_6ma and R_MI areas within the cingulo-opercular network) associated with the 4q24 locus (index variant rs10013053).

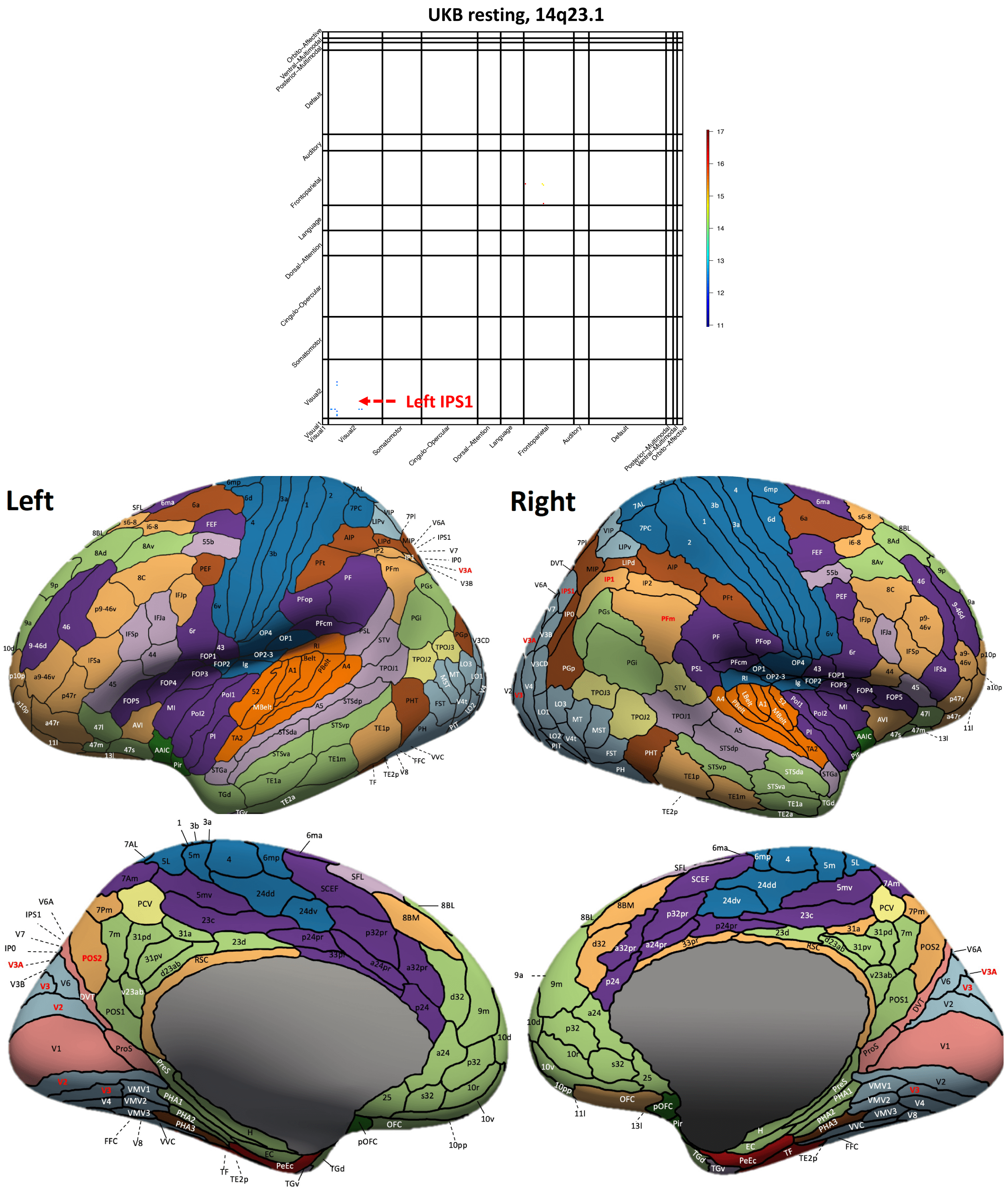


**Fig. S39 Functional areas and networks associated with 14q23.1 in resting-state fMRI.**

Top: map of -Log$_{10}$(Pvalue) for significant associations ($P < 2.93 \times 10^{-12}$). Bottom: location of the functional areas that having significant associations (names highlighted in red). Background colors indicate different functional networks, whose names can be found in Figure S1.

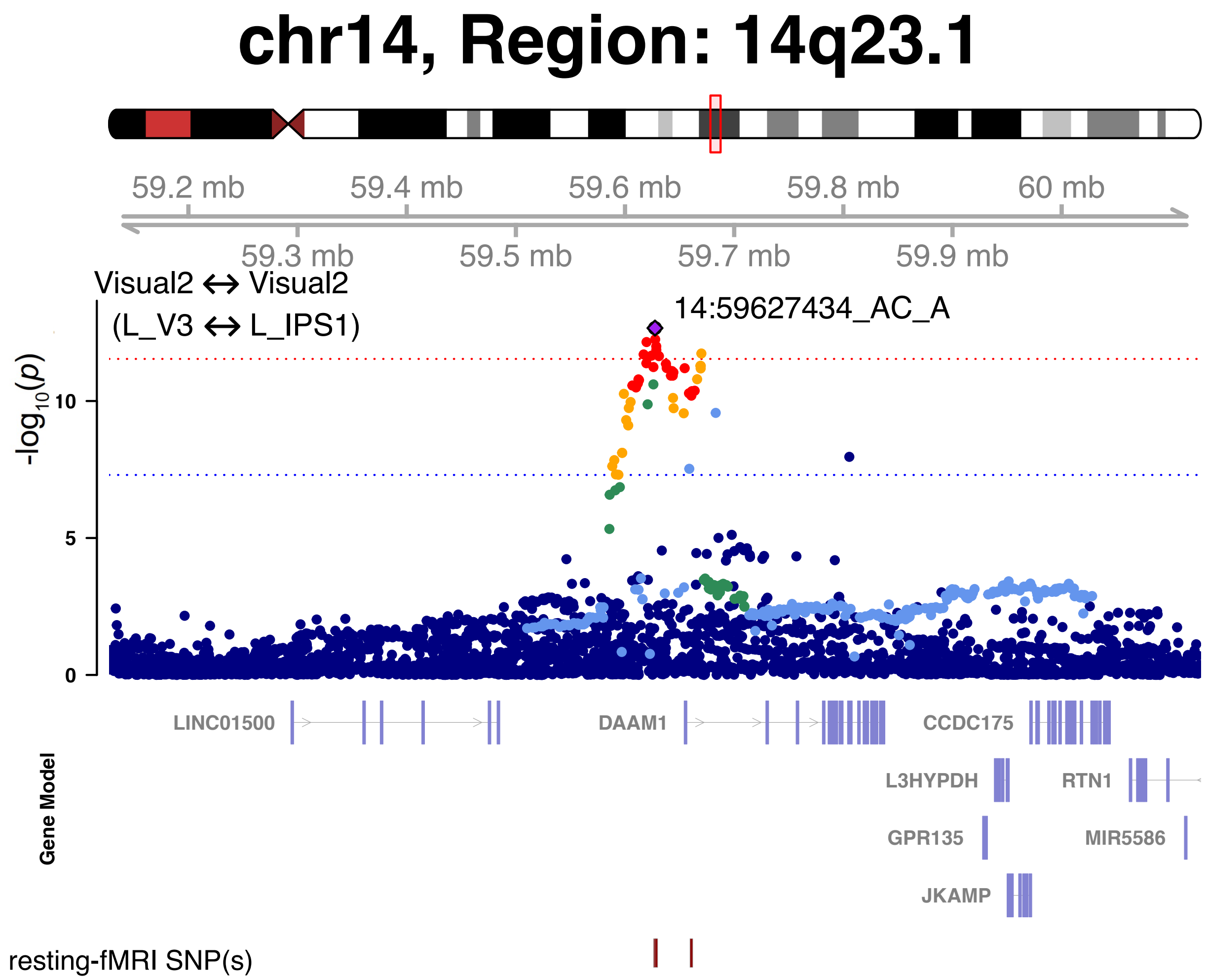


**Fig. S40 Selected functional areas and networks associated with 14q23.1 in resting-state fMRI.**

We illustrate one functional connectivity trait (between the L_V3 and L_IPS1 areas within the secondary visual network) associated with the 14q23.1 locus (index variant 14:59627434_AC_A).

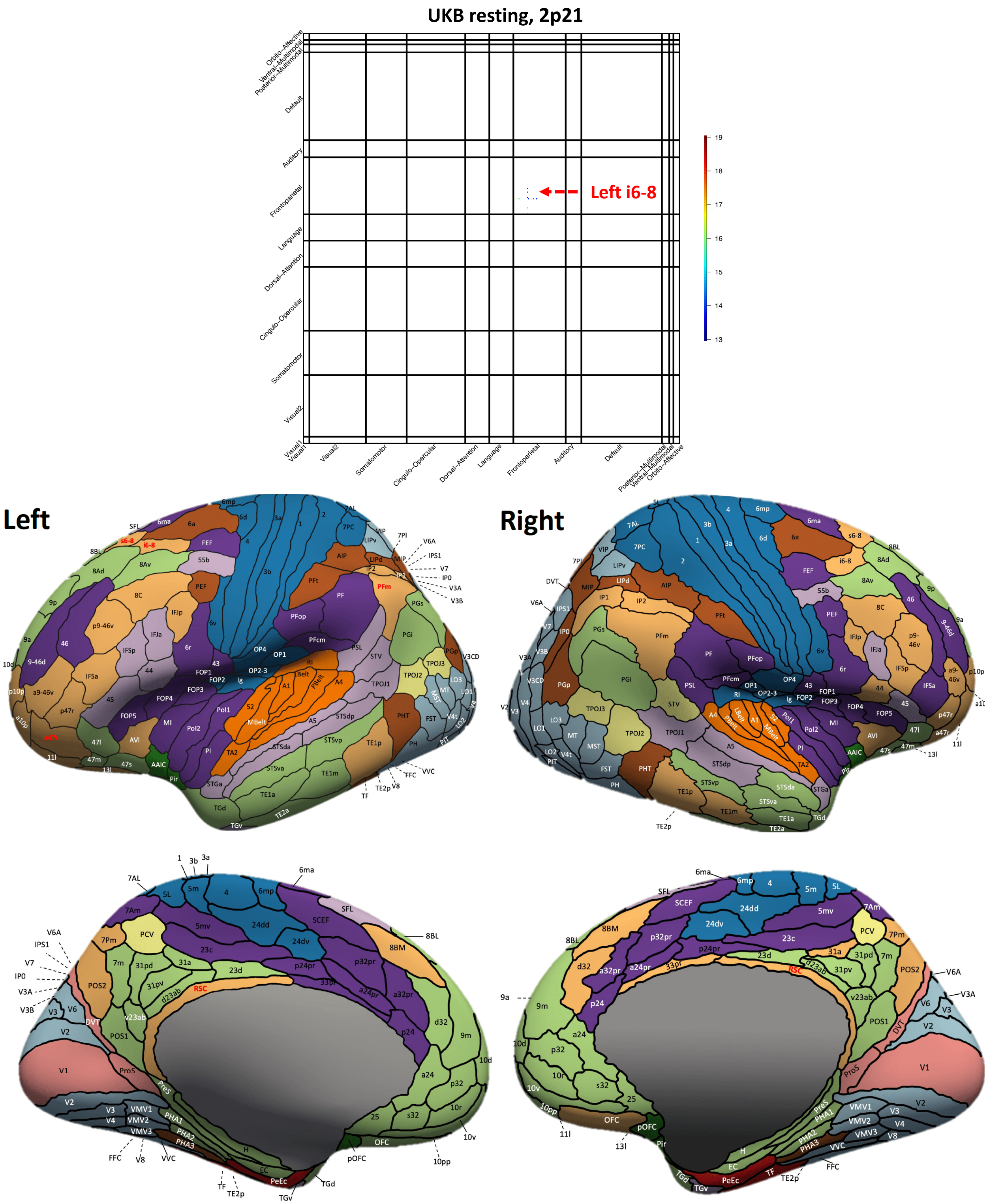


**Fig. S41 Functional areas and networks associated with 2p21 in resting-state fMRI.**

Top: map of -Log$_{10}$(Pvalue) for significant associations ($P < 2.93 \times 10^{-12}$). Bottom: location of the functional areas that having significant associations (names highlighted in red). Background colors indicate different functional networks, whose names can be found in Figure S1.

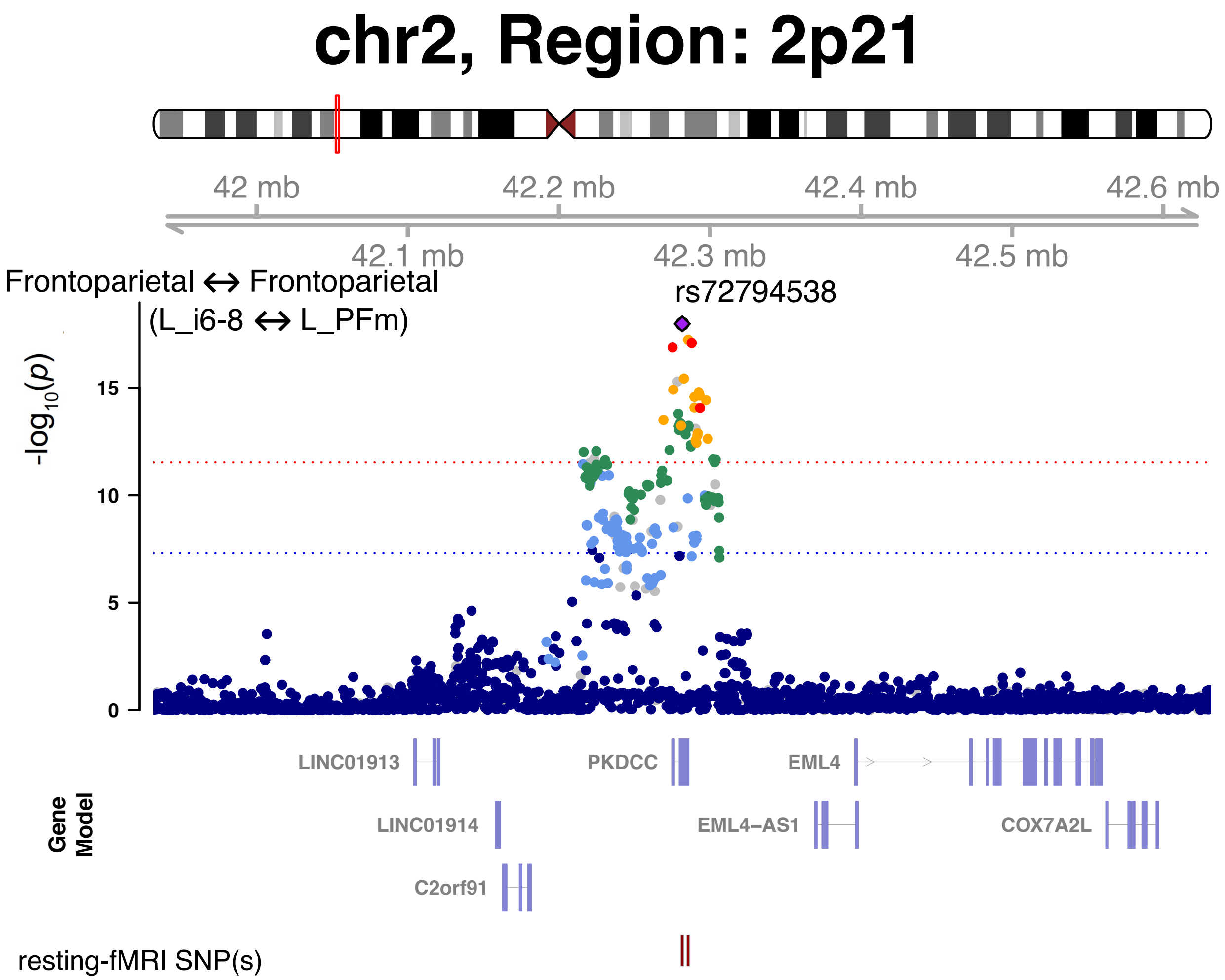


**Fig. S42 Selected functional areas and networks associated with 2p21 in resting-state fMRI.**

We illustrate one functional connectivity trait (between the L_i6-8 and L_PFm areas within the frontoparietal network) associated with the 2p21 locus (index variant rs72794538).

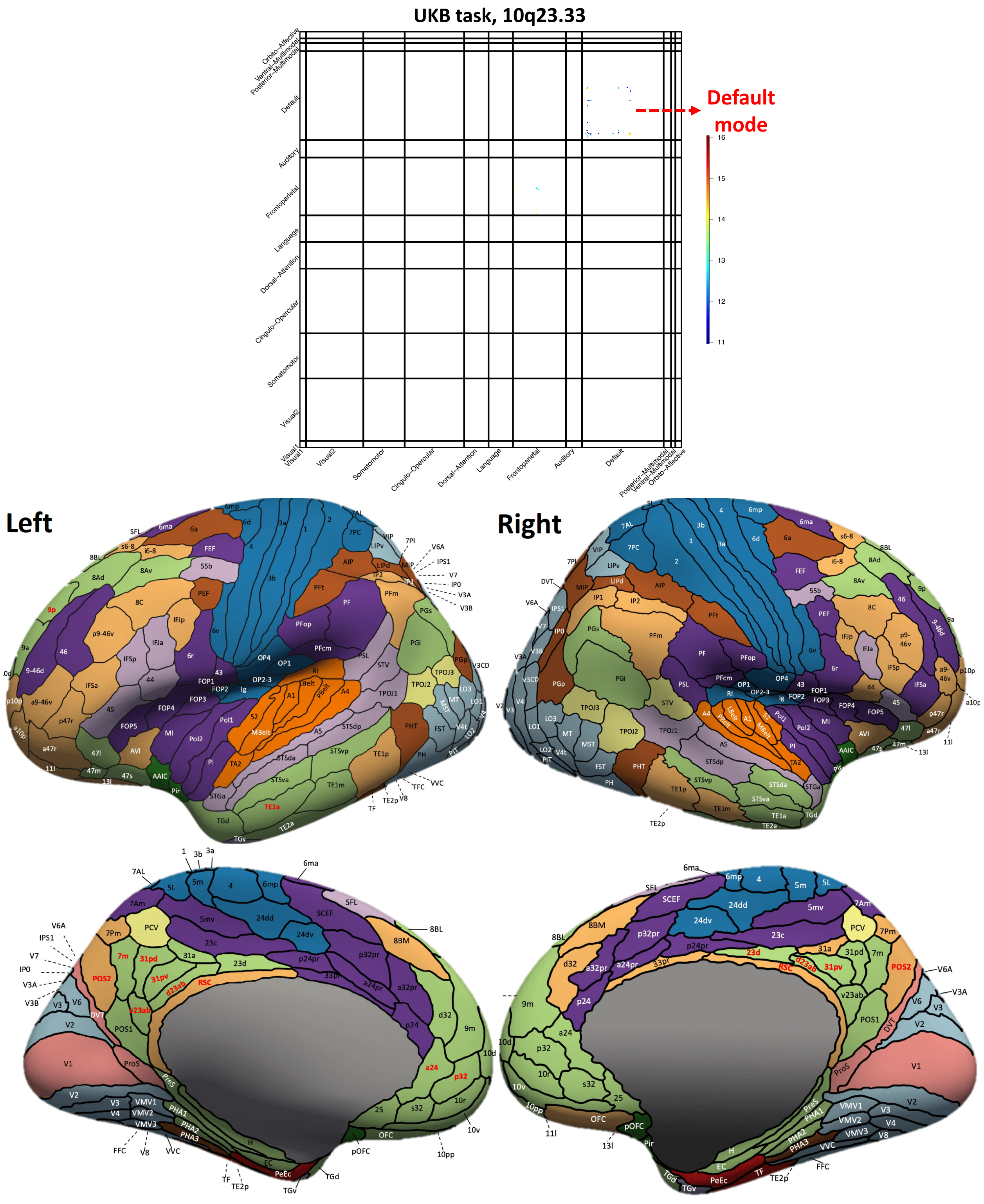


**Fig. S43 Functional areas and networks associated with 10q23.33 in task-evoked fMRI.**

Top: map of -Log$_{10}$(Pvalue) for significant associations ($P < 2.93 \times 10^{-12}$). Bottom: location of the functional areas that having significant associations (names highlighted in red). Background colors indicate different functional networks, whose names can be found in Figure S1.

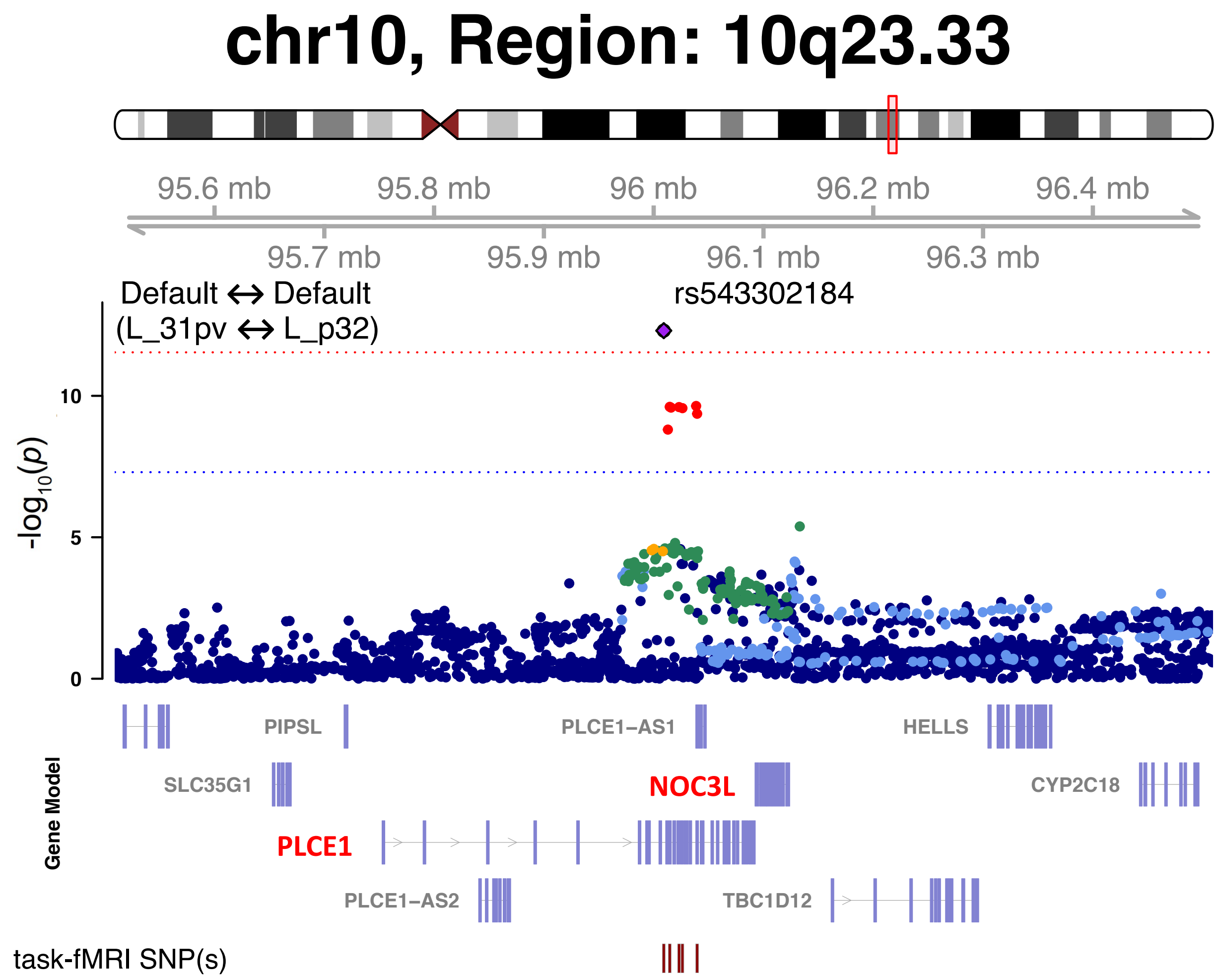


**Fig. S44 Selected Functional areas and networks associated with 10q23.33 in task-evoked fMRI.**

We illustrate one functional connectivity trait (between the L_31pv and L_p32 areas within the frontoparietal network) associated with the 10q23.33 locus (index variant rs543302184). The index variant and its proxy variants (LD $r^2 \geq 0.8$) are known brain eQTLs (https://metabrain.nl/) for the genes *PLCE1* and *NOC3L* (bolded in red).

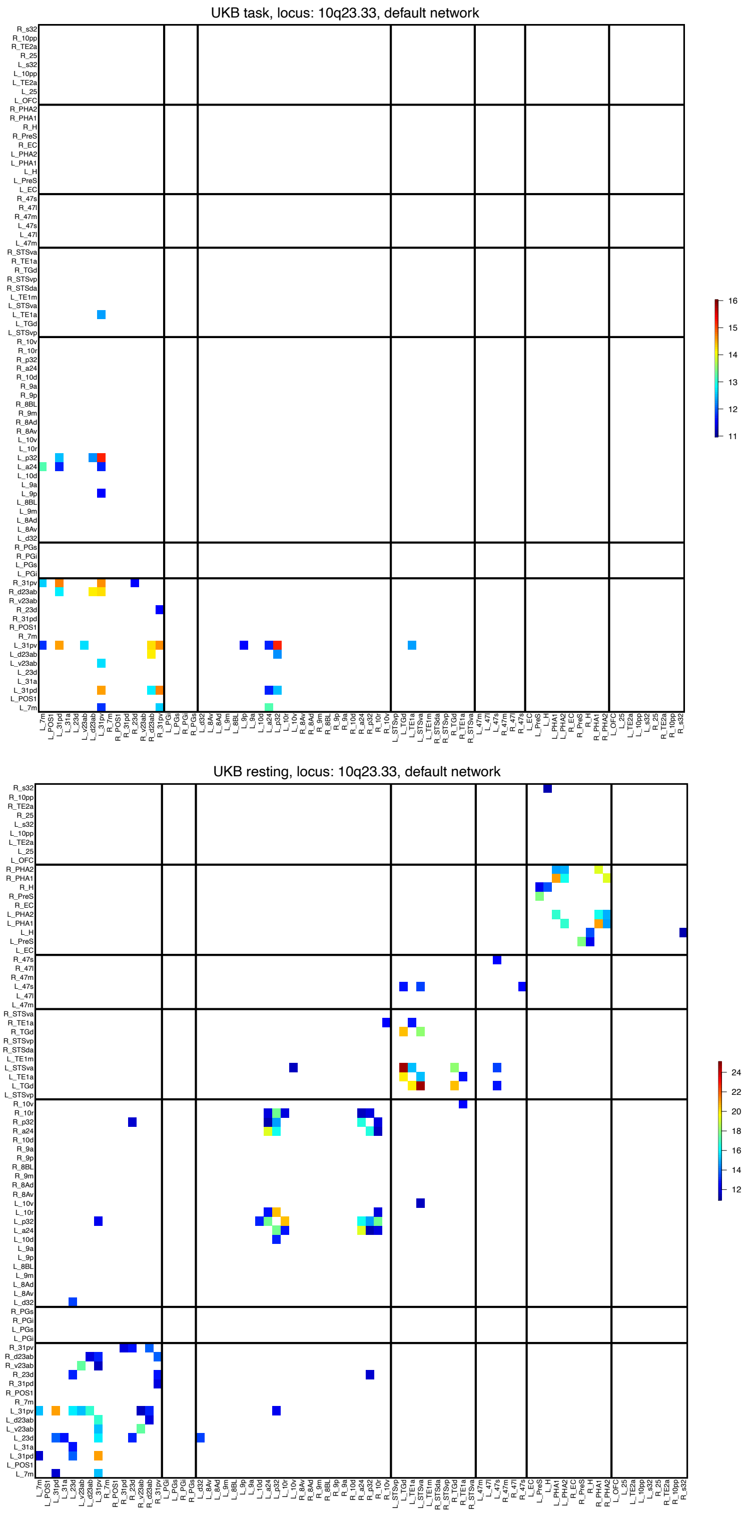


**Fig. S45 Functional areas associated with 10q23.33 in the default mode network.**

We show the -Log$_{10}$(Pvalue) map for significant associations of 10q23.33 in resting-state (top) and task-evoked fMRI (bottom) ($P$ < 2.93 × $10^{-12}$). The location of the functional areas and subclusters of the default mode network can be found in Figure S9.

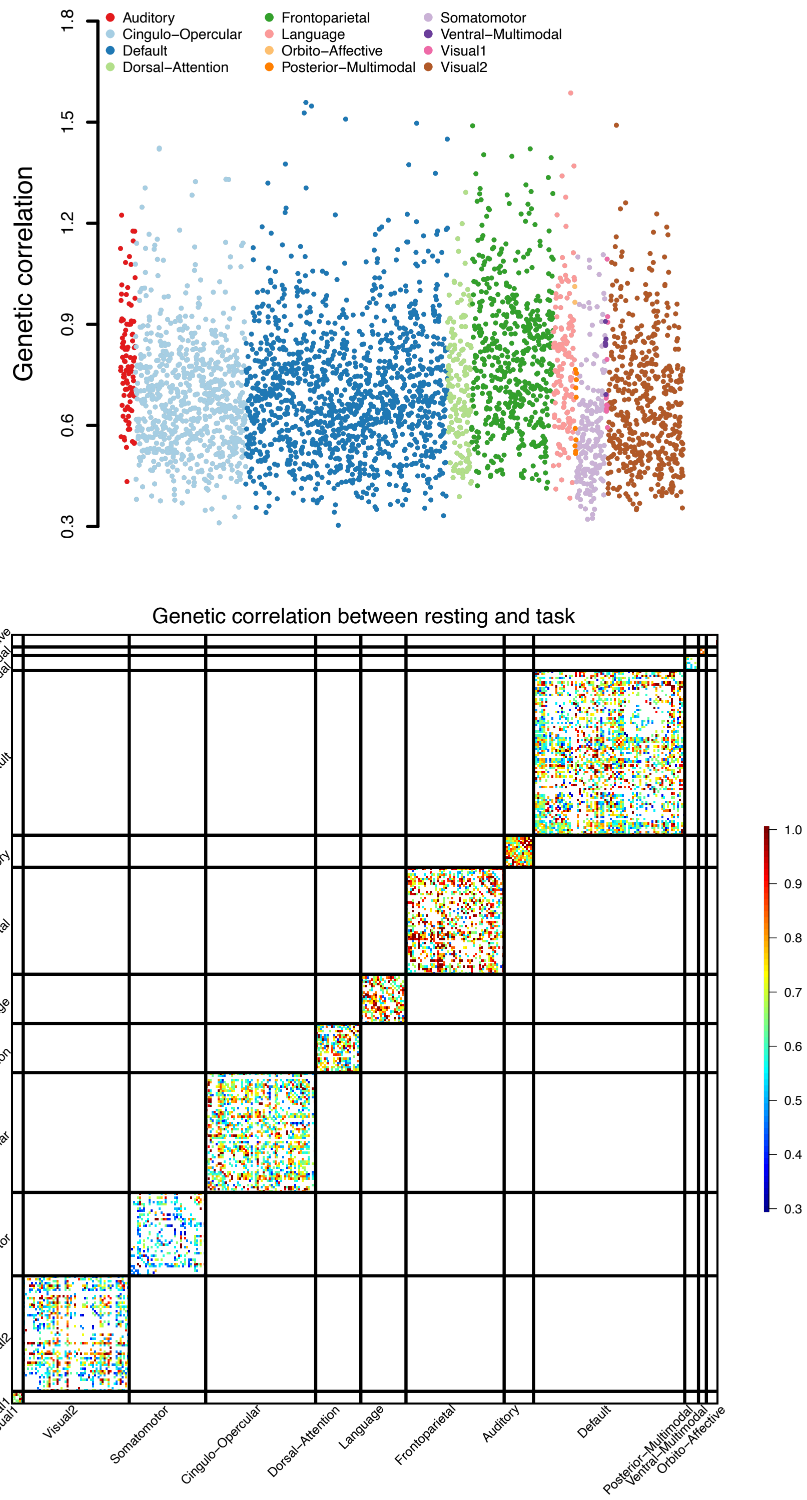


**Fig. S46 Significant genetic correlations between resting-state and task-evoked fMRI.**

The genetic correlations were estimated by the cross-trait LDSC (https://github.com/bulik/ldsc/). We illustrate the significant estimates at the FDR 5% level.

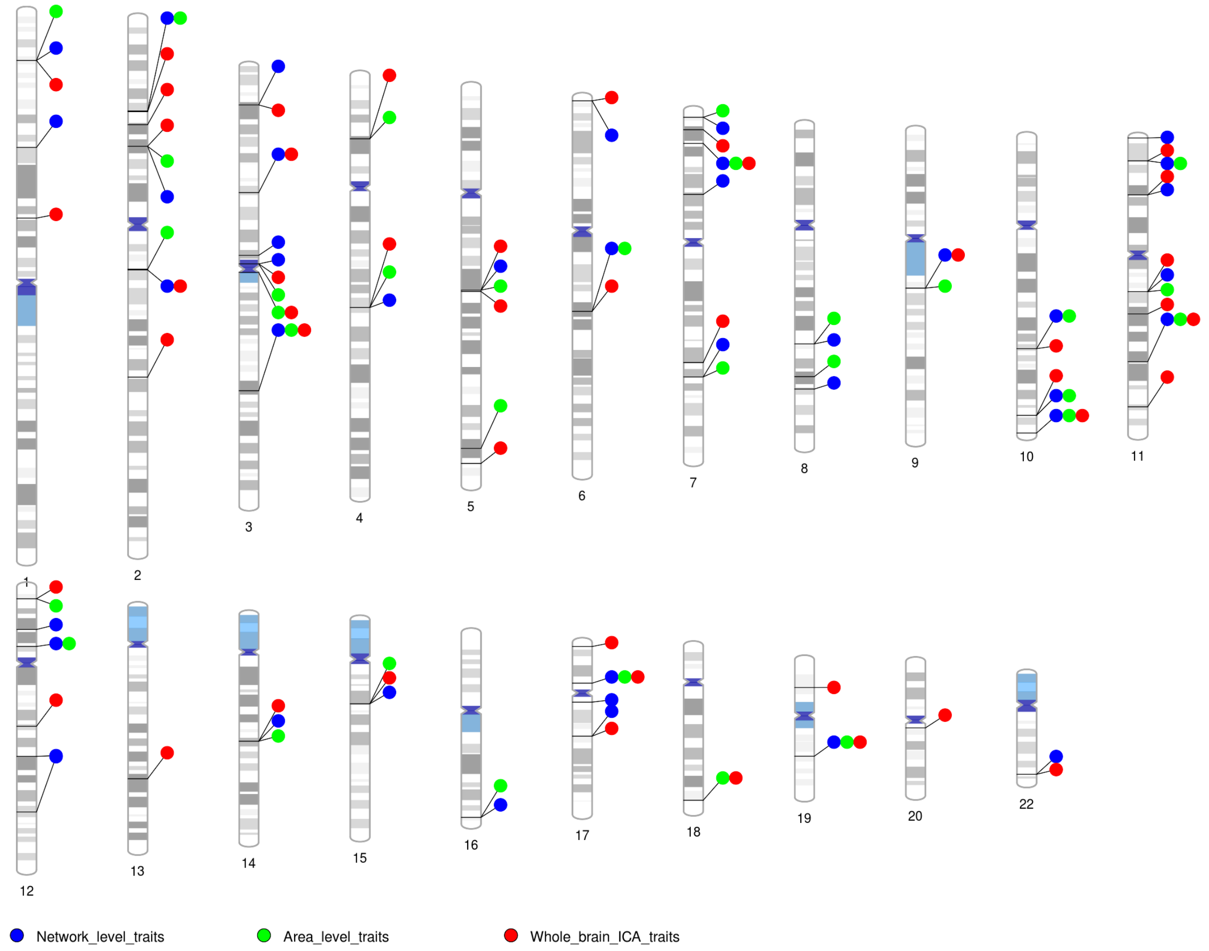


**Fig. S47 Ideogram of genomic regions influencing fMRI traits.**

There were 45 previously identified regions for whole brain ICA-based fMRI traits (Whole_brain_ICA_traits) and 47 regions for fMRI traits identified in the present study (Network_level traits and Area_level_traits). Different colors represent the three categories of fMRI traits. Each signal dot indicates that at least one of the fMRI traits from this trait category was associated with the genomic region.

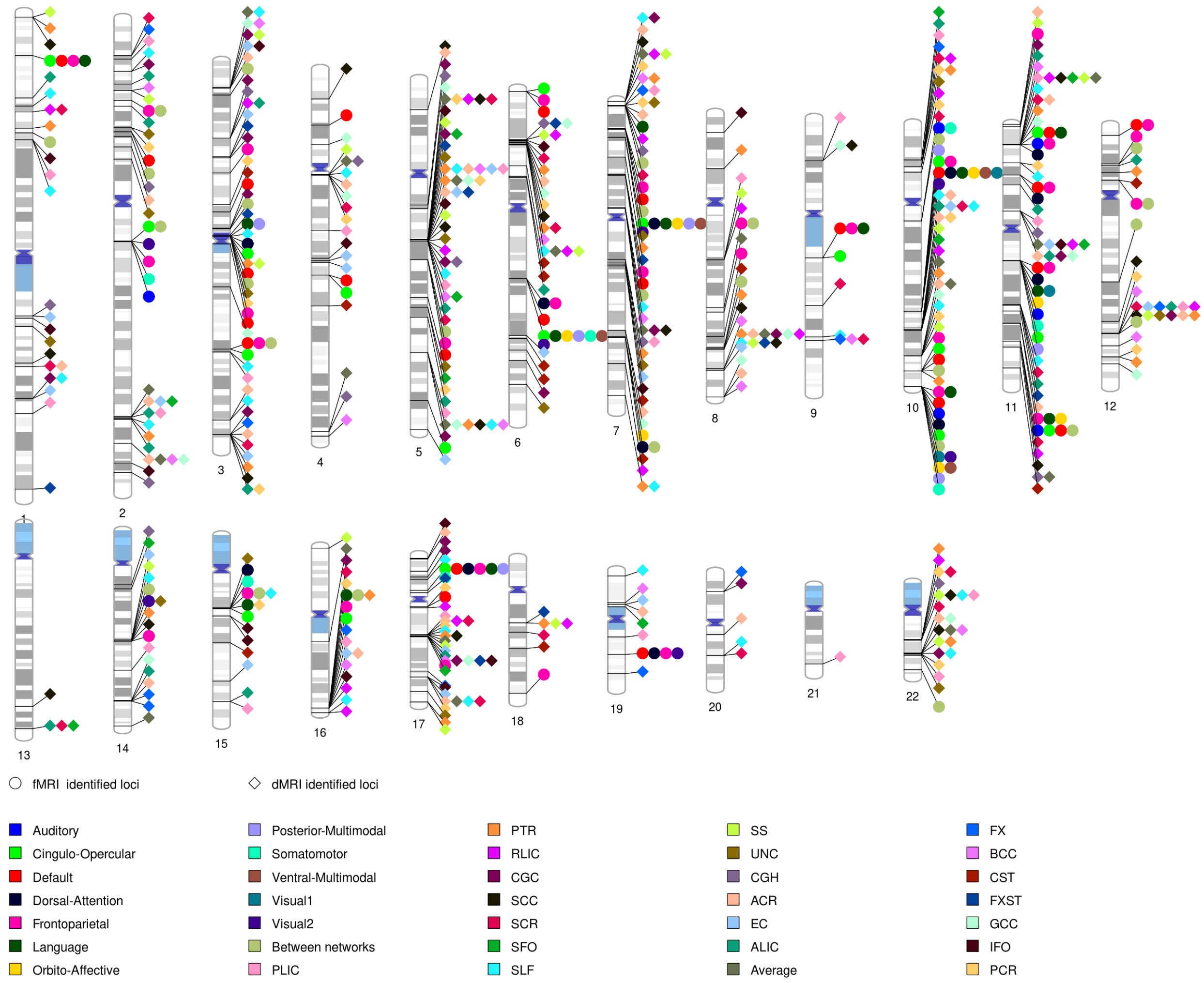


**Fig. S48 Ideogram of genomic regions influencing DTI parameters and fMRI traits.**

There were 151 previously identified regions for DTI parameters and 47 regions for fMRI traits identified in the present study. The colors represent the 21 white matter tracts (and the global average) as well as the 12 functional networks (and the between network functional connectivity). Each signal dot indicates that at least one of the DTI parameters of this tract or fMRI traits of that network was associated with the genomic region.

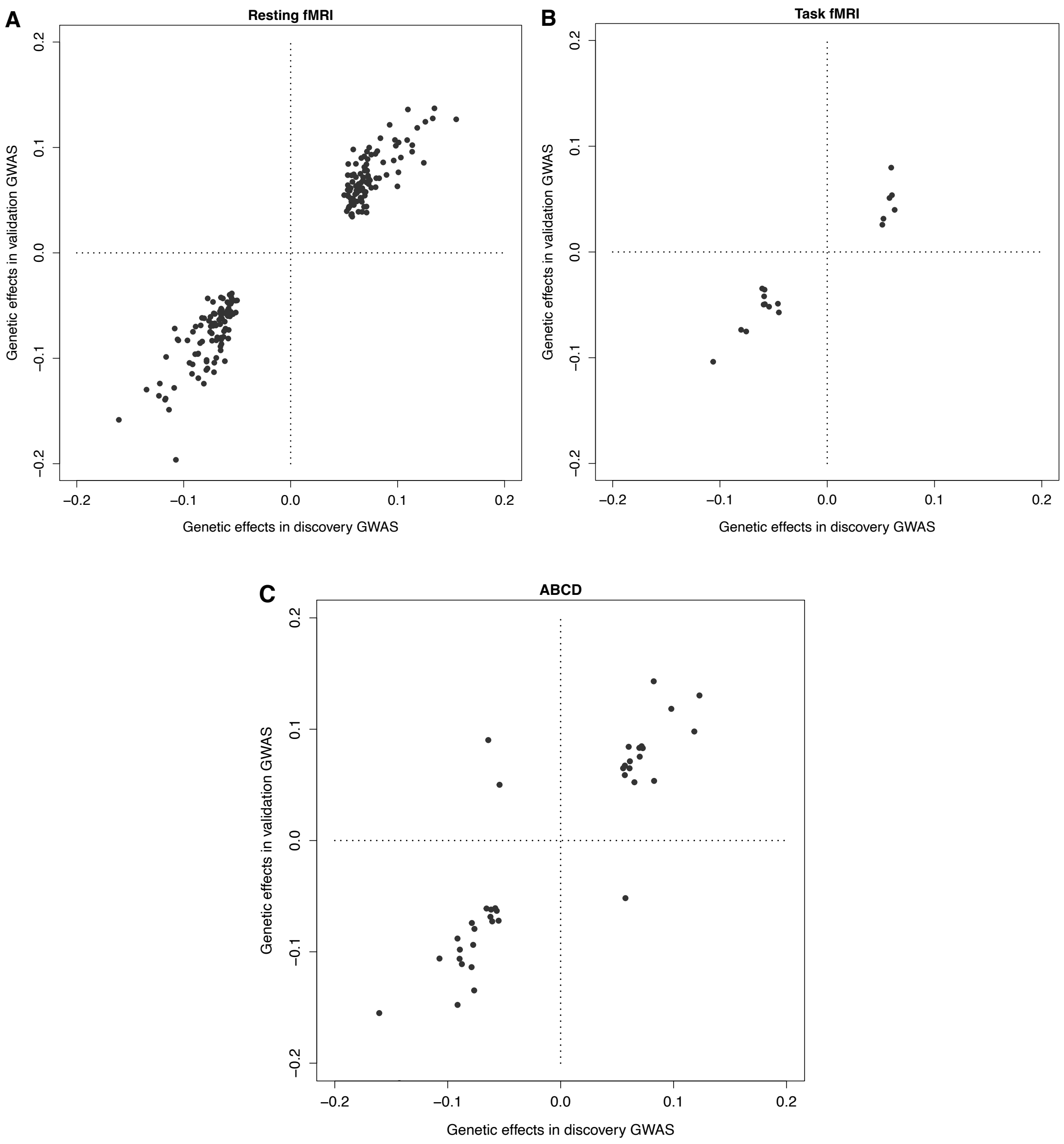


**Fig. S49 Genetic effects in the UKB discovery GWAS and validation GWAS.**

We show the genetic effects (the beta estimates) of associations that passed the nominal significance level (0.05) in the validation GWAS of UKB resting fMRI (**A**), UKB task fMRI (**B**), and ABCD resting fMRI (**C**). Most of these associations had the same effect signs in the discovery GWAS (x axis) and validation GWAS (y axis).

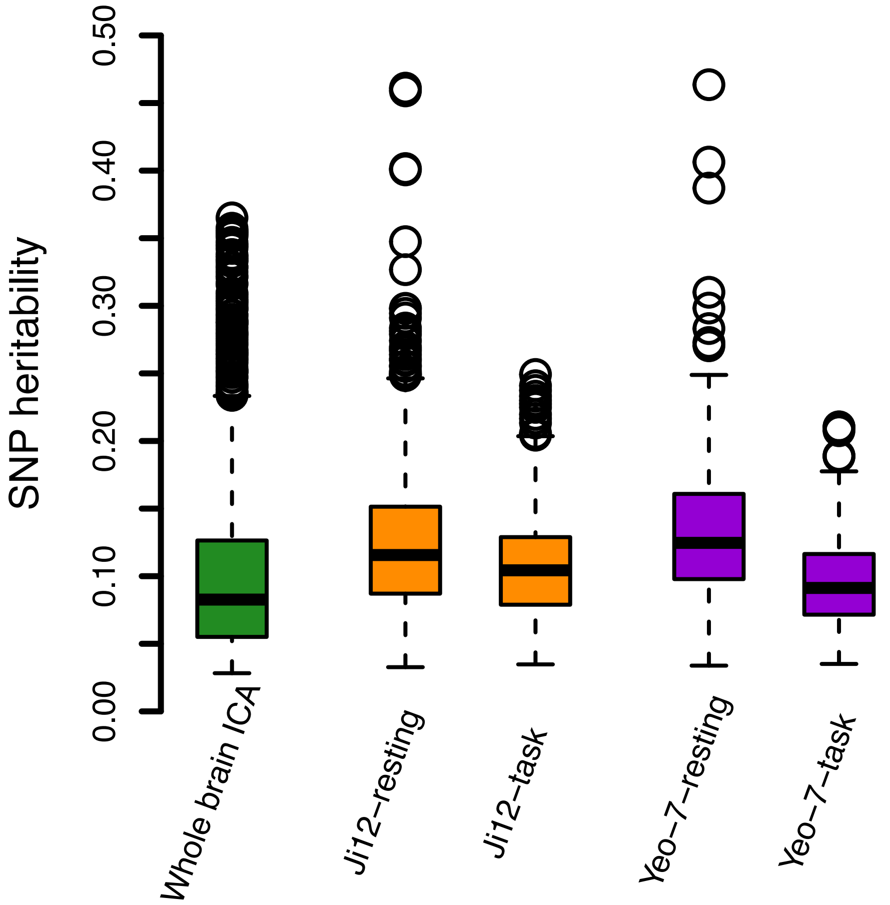


**Fig. S50 SNP heritability estimates from different brain maps.**

Heritability was estimated in both resting-state fMRI and task-evoked fMRI using UKB individuals of British ancestry (*n* = 34,641 for resting and 32,144 for task). We show the results of whole brain ICA (resting fMRI), the 12 networks used in our main analyses (Ji12-resting and Ji12-task), and the Yeo-7 networks (Yeo-7-resting and Yeo-7-task).

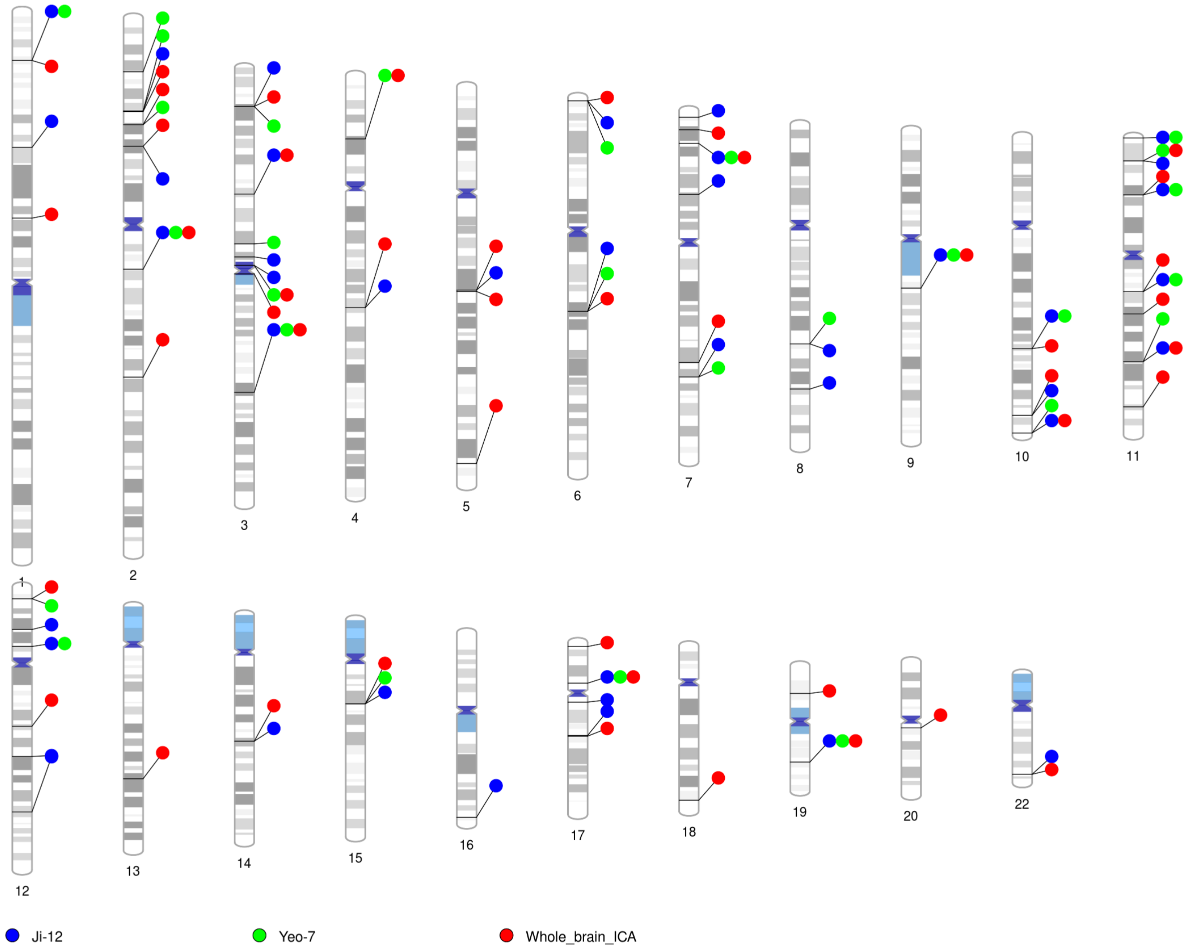


**Fig. S51 Ideogram of genomic regions identified by different brain maps.**

There were 61 genomic regions identified by either whole brain ICA-based fMRI traits (Whole_brain_ICA), network-level fMRI traits defined by the 12 networks (Ji-12), or network-level fMRI traits defined by the Yeo-7 networks (Yeo-7). Different colors represent the three categories of fMRI traits. Each signal dot indicates that at least one of the fMRI traits from this trait category was associated with the genomic region.

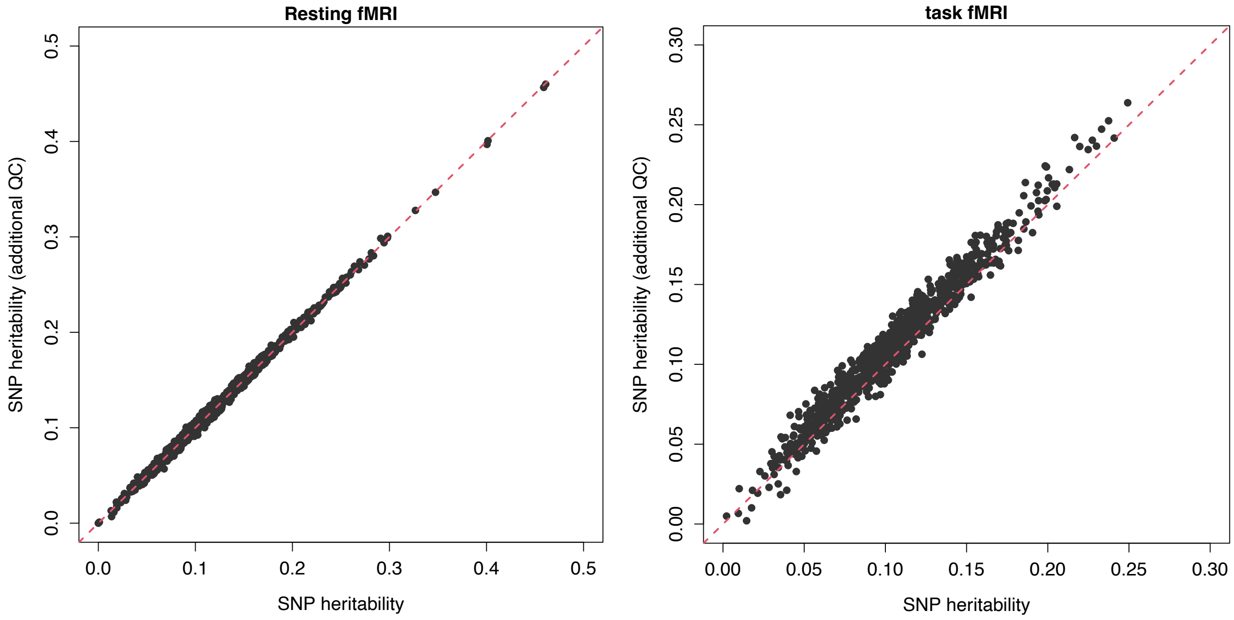


**Fig. S52 SNP heritability estimates before and after additional QC steps.**

We show the results of network-level traits in resting fMRI (left panel) and task fMRI (right panel).

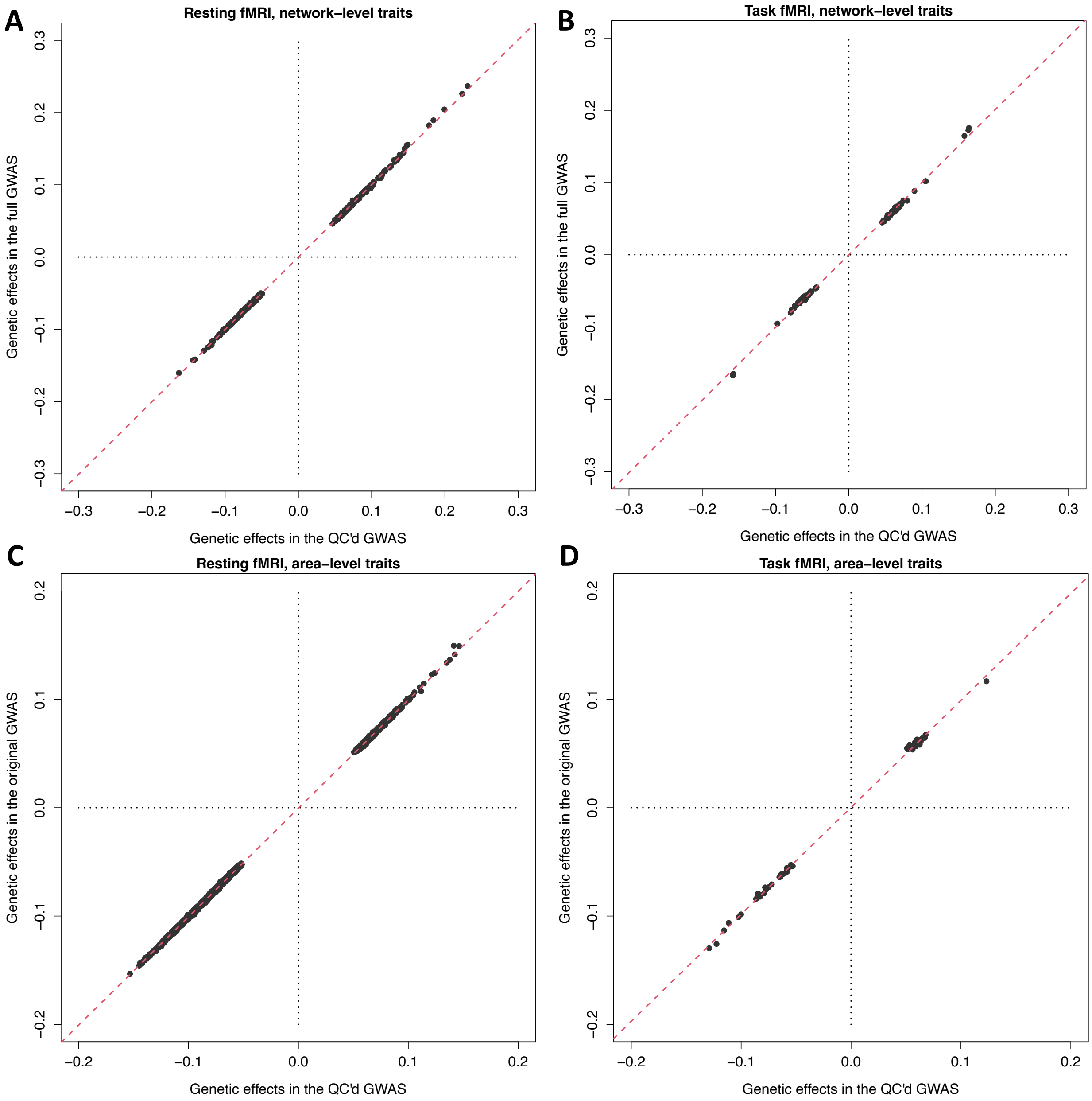


**Fig. S53 Genetic effects before and after additional QC steps.**

We show the genetic effects (the beta estimates) of associations before and after additional QC steps for network-level (**A** for resting fMRI and **B** for task fMRI) and area-level traits (**C** for resting fMRI and **D** for task fMRI).

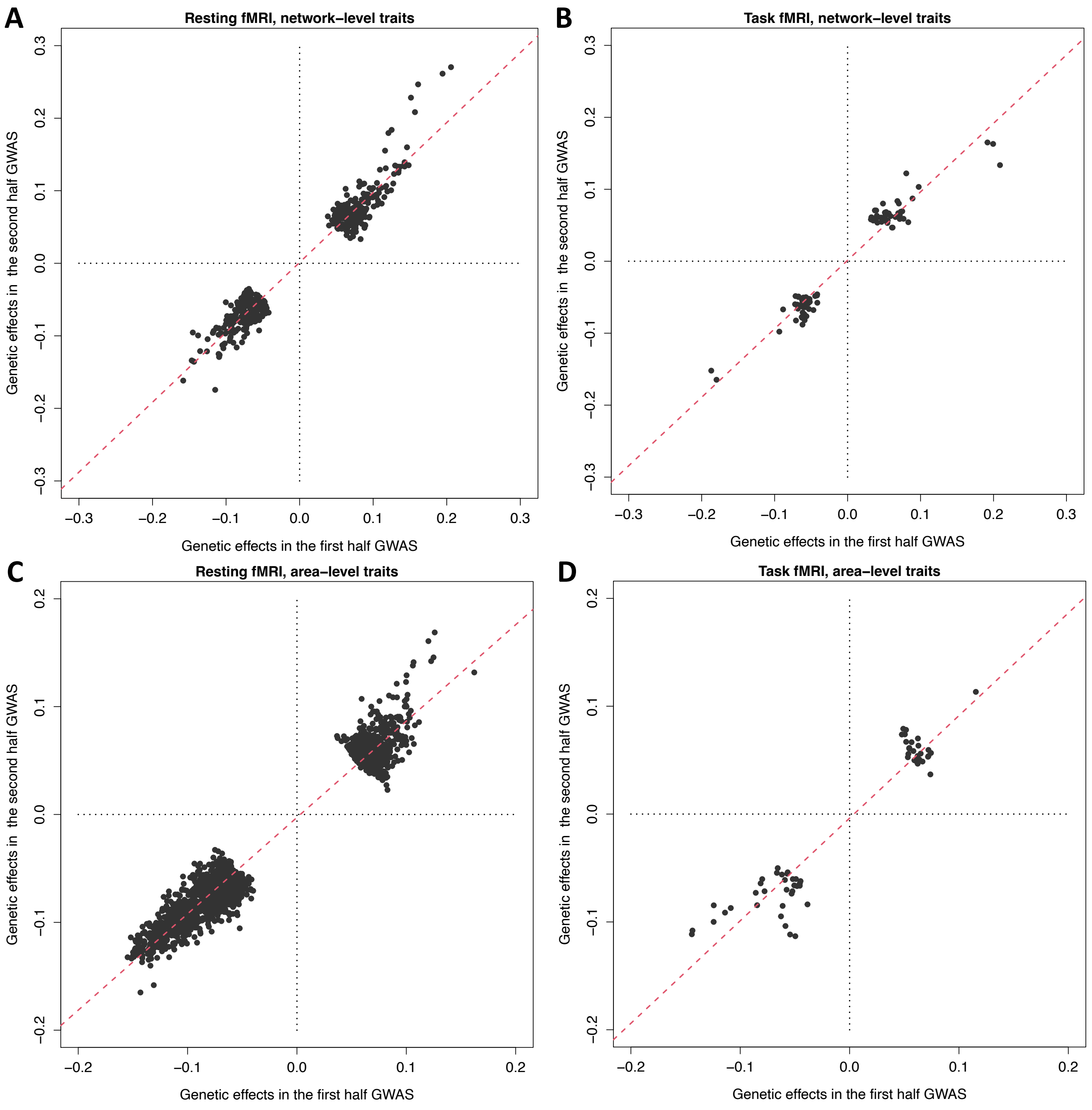


**Fig. S54 Genetic effects in split-half GWAS data analysis.**

For the significant genetic associations identified by our discovery GWAS, we show their estimated genetic effects (the beta estimates) in the first half (x-axis) and the second half (y-axis) GWAS. **A**: network-level traits in resting fMRI; **B**: network-level traits in task fMRI; **C**: area-level traits in resting fMRI; and **D**: area-level traits in task fMRI.

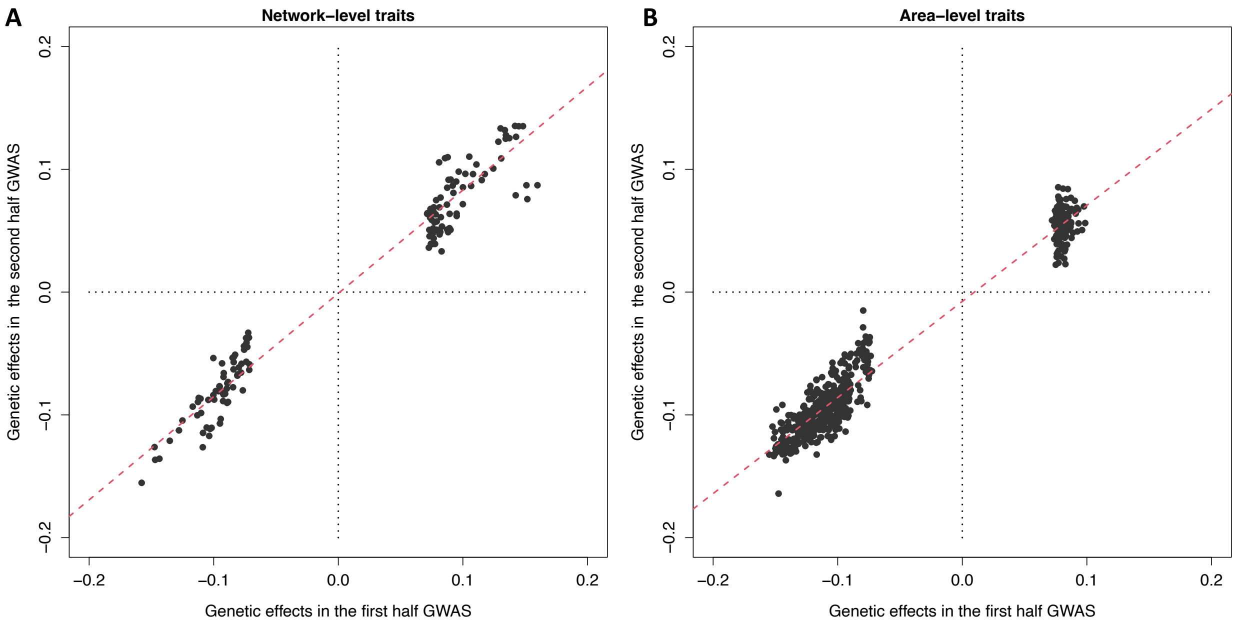


**Fig. S55 Genetic effects in split-half GWAS data analysis.**

We use the first half data as the discovery GWAS to identify significant associations and show their estimated genetic effects (the beta estimates) in the first half (x-axis) and the second half (y-axis) GWAS. **A**: network-level traits in both resting and task fMRI; **B**: area-level traits in both resting and task fMRI.

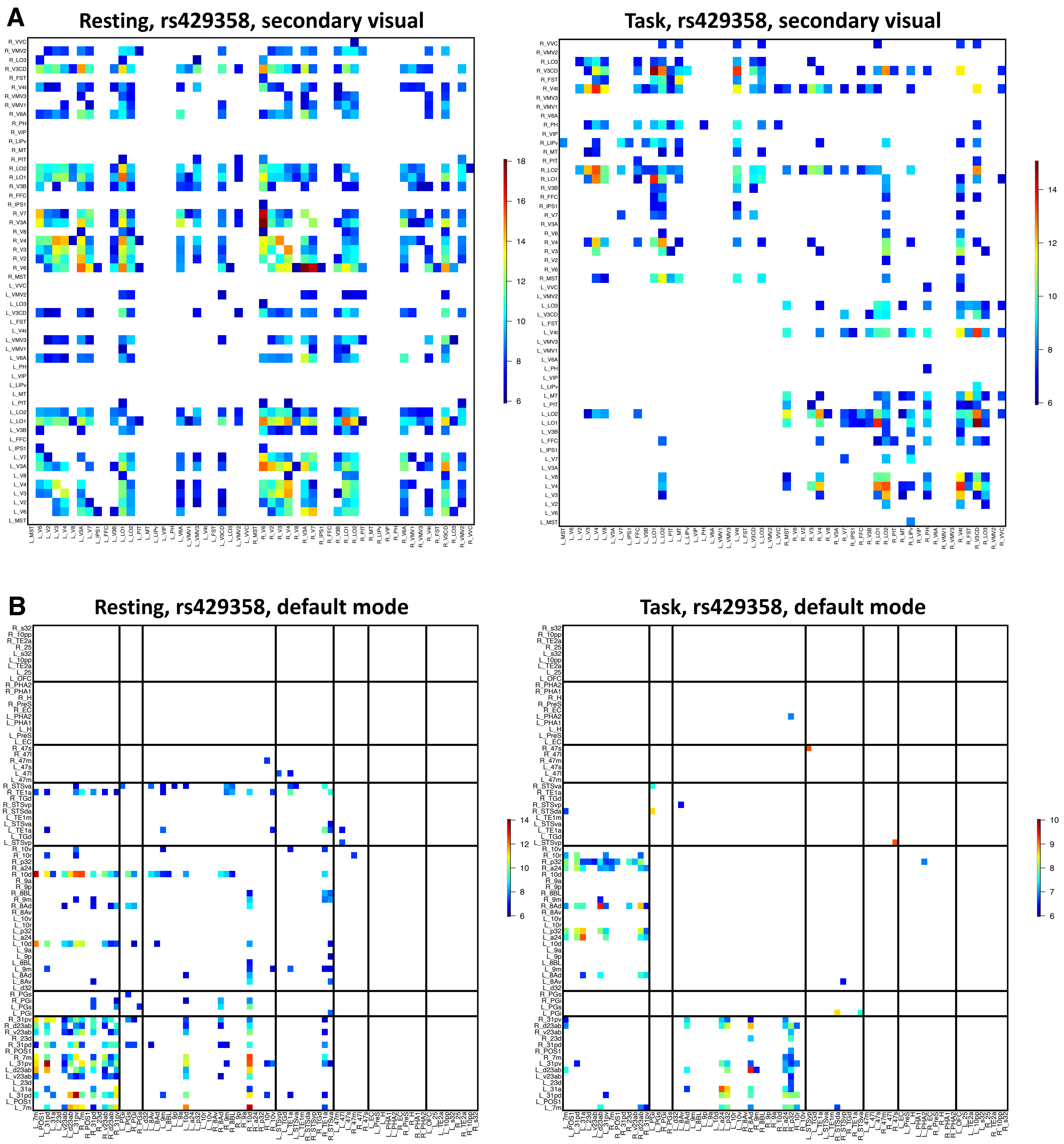


**Fig. S56 Functional areas in the secondary visual and default mode networks associated with index variant of Alzheimer's disease (rs429358) in variant-specific analysis.**

**A**: maps of -$\log_{10}$(Pvalue) for significant associations at Bonferroni significance level ($3.86 \times 10^{-7}$, 0.05/64,620/2) in the secondary visual network. **B**: maps of -$\log_{10}$(Pvalue) for significant associations at Bonferroni significance level ($3.86 \times 10^{-7}$) in the default mode network. The location of the functional areas and subclusters of the default mode network can be found in Figure S9.

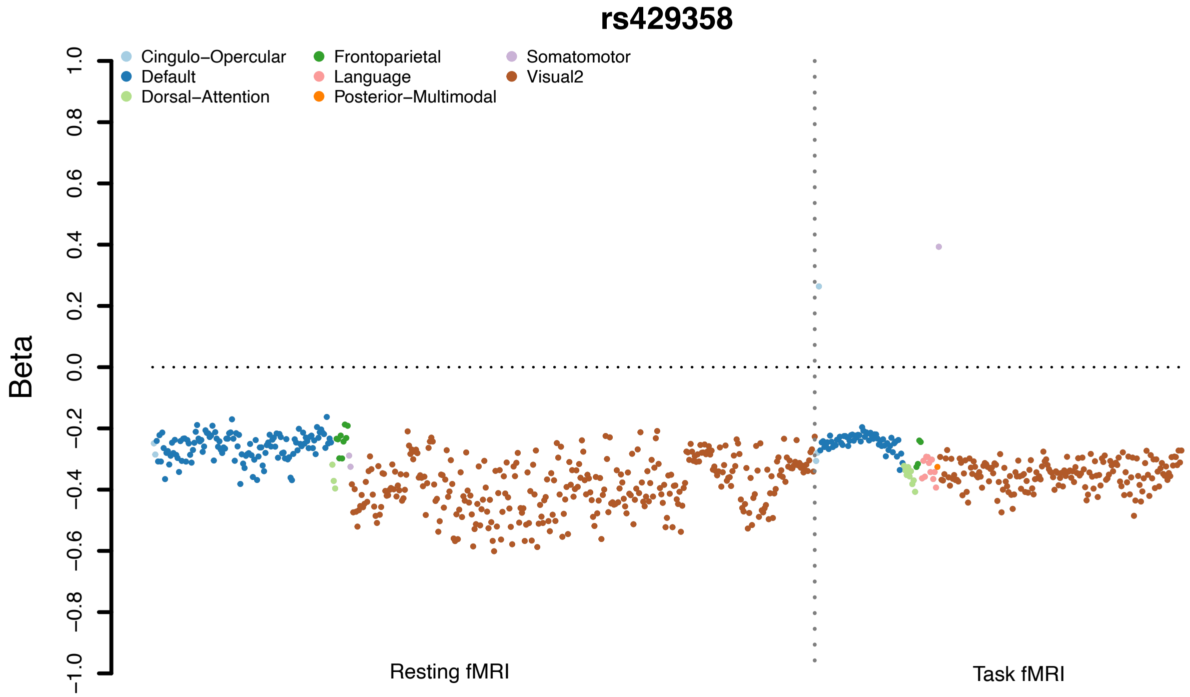


**Fig. S57 Significant genetic effects of rs429358 in the variant-specific analysis of fMRI traits.**

We illustrate the significant ($P$ < 3.86 × $10^{-7}$, Bonferroni significance level in variant-specific analysis) genetic effects of the rs429358 (tested allele: C, MAF 0.15) on resting and task fMRI traits.

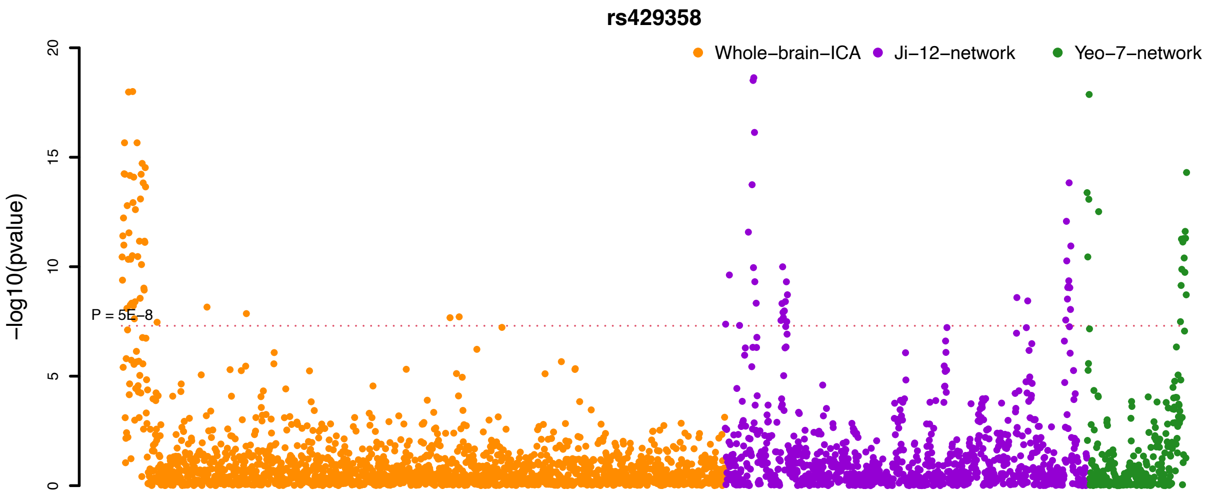


**Fig. S58: Associations between the rs429358 and resting fMRI traits from different brain maps.**

We show the $-\log_{10}$(Pvalue) for associations between the rs429358 and whole brain ICA-based fMRI traits (Whole-brain-ICA), network-level fMRI traits defined by the 12 networks (Ji-12-network), and network-level fMRI traits defined by the Yeo-7 networks (Yeo-7-network). Different colors represent the three categories of fMRI traits. The horizontal dashed line indicates the conventional GWAS significance level ($5 \times 10^{-8}$).

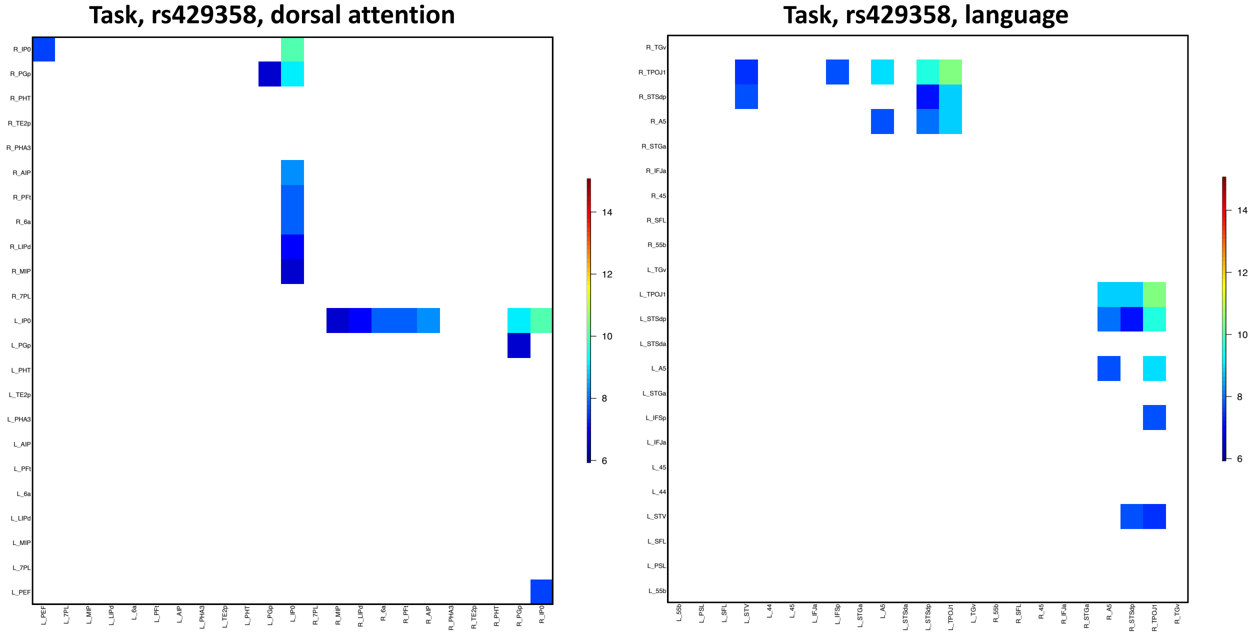


**Fig. S59 Functional areas in the dorsal attention and language networks associated with index variant of Alzheimer's disease (rs429358) in variant-specific analysis of task-evoked fMRI.**

Left: maps of -$Log_{10}$(Pvalue) for significant associations at Bonferroni significance level ($3.86 \times 10^{-7}$, 0.05/64,620/2) in the dorsal attention network. Right: maps of -$Log_{10}$(Pvalue) for significant associations at Bonferroni significance level ($3.86 \times 10^{-7}$) in the language network.

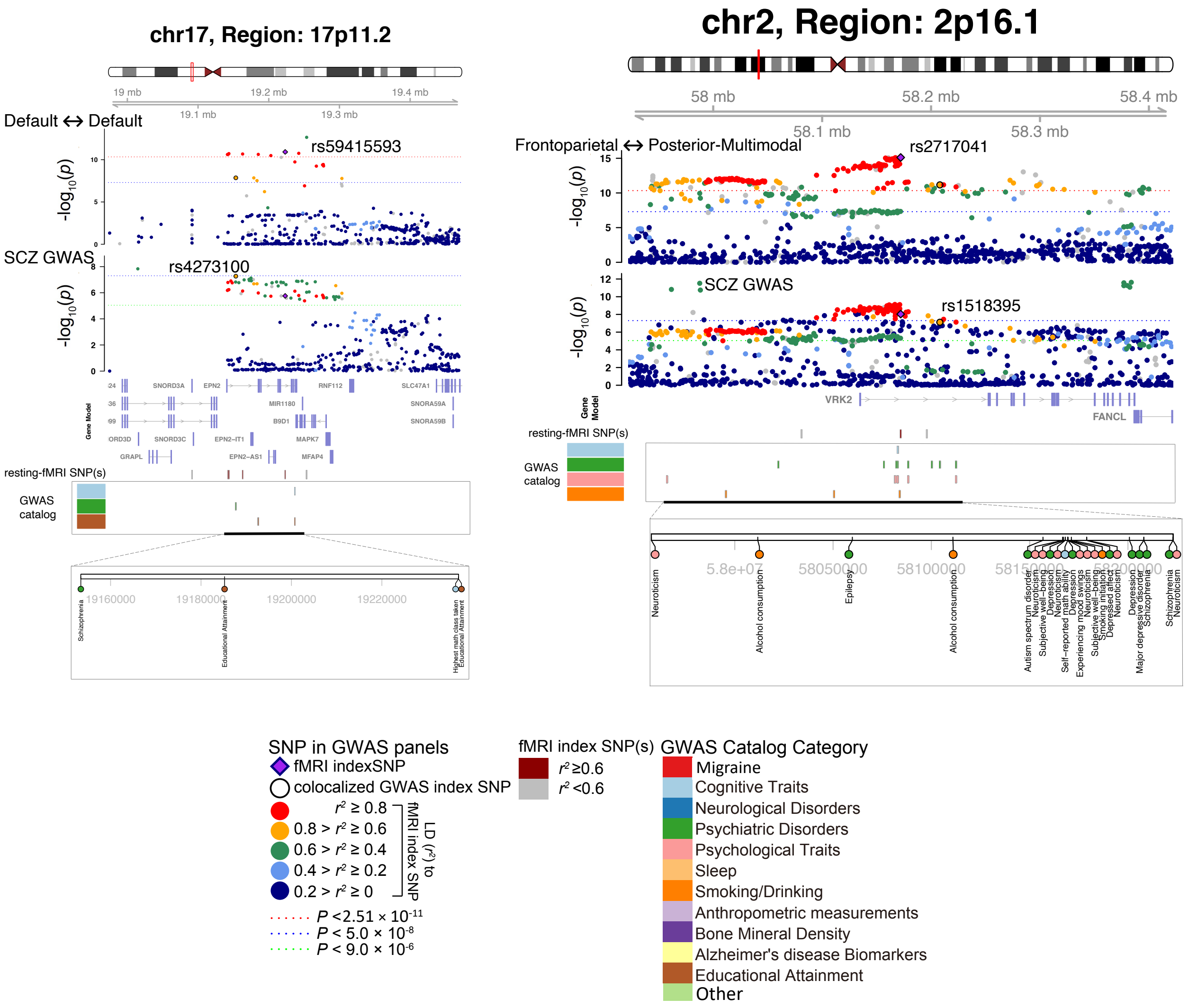


**Fig. S60 Selected genetic loci that were associated with both fMRI traits and psychiatric disorders.**

In 17p11.2 and 2p16.1, we observed shared genetic influences (LD $r^2 \geq 0.6$) between resting-state fMRI traits (index variants rs59415593 and rs2717041) and schizophrenia (SCZ, index variants rs4273100 and rs1518395). Default <-> Default, functional connectivity within the default mode network; Frontoparietal <-> Posterior-Multimodal, functional connectivity between the frontoparietal and the default mode networks.

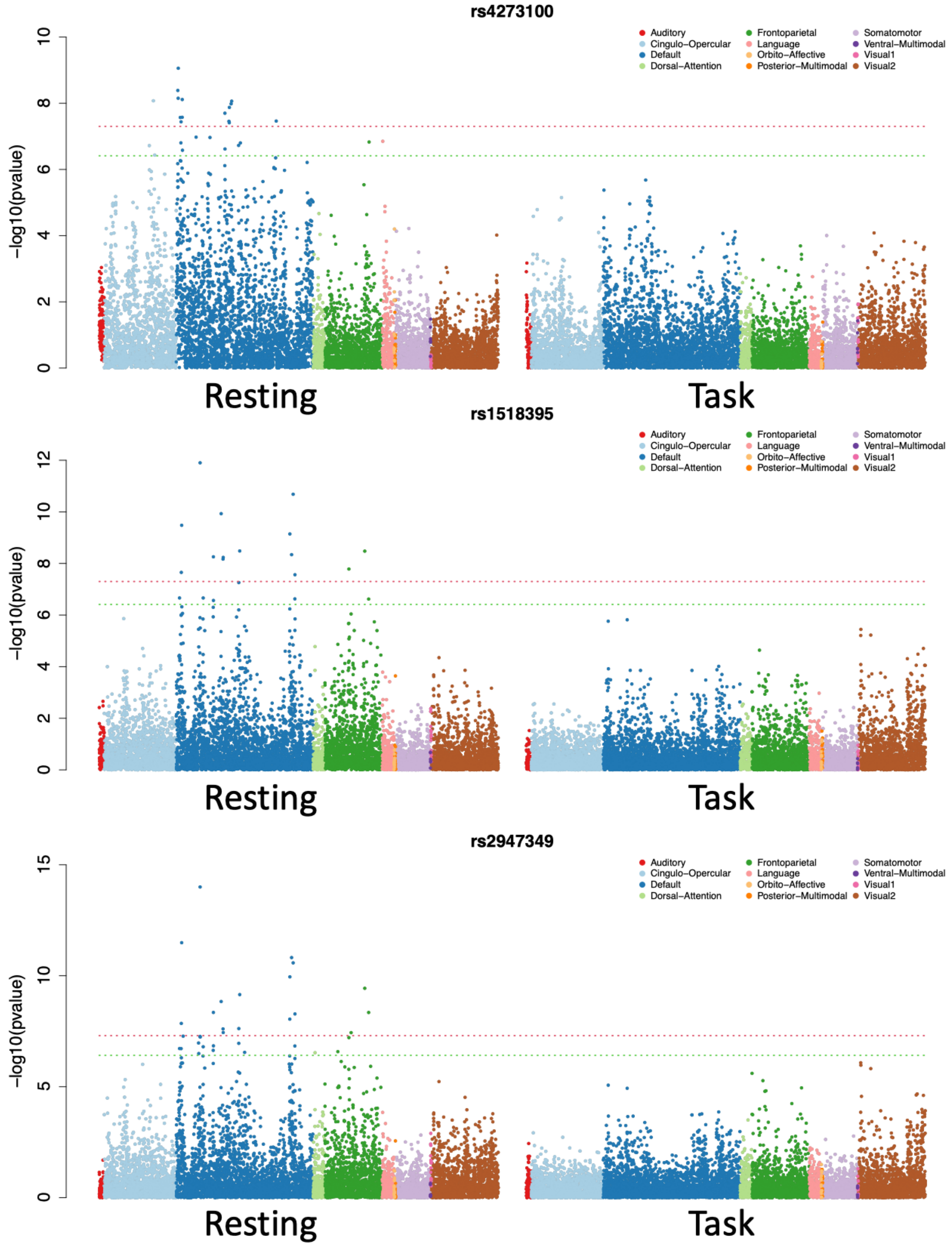


**Fig. S61 Associations between fMRI traits and the index genetic variants of schizophrenia.**

We performed variant-specific association analysis with rs4273100, rs1518395, and rs2947349 in both resting and task fMRI. The red and green lines indicate the genome-wide significant level $5 \times 10^{-8}$ and the Bonferroni significance level of this variant-specific analysis ($3.86 \times 10^{-7}$, 0.05/64,620/2), respectively.

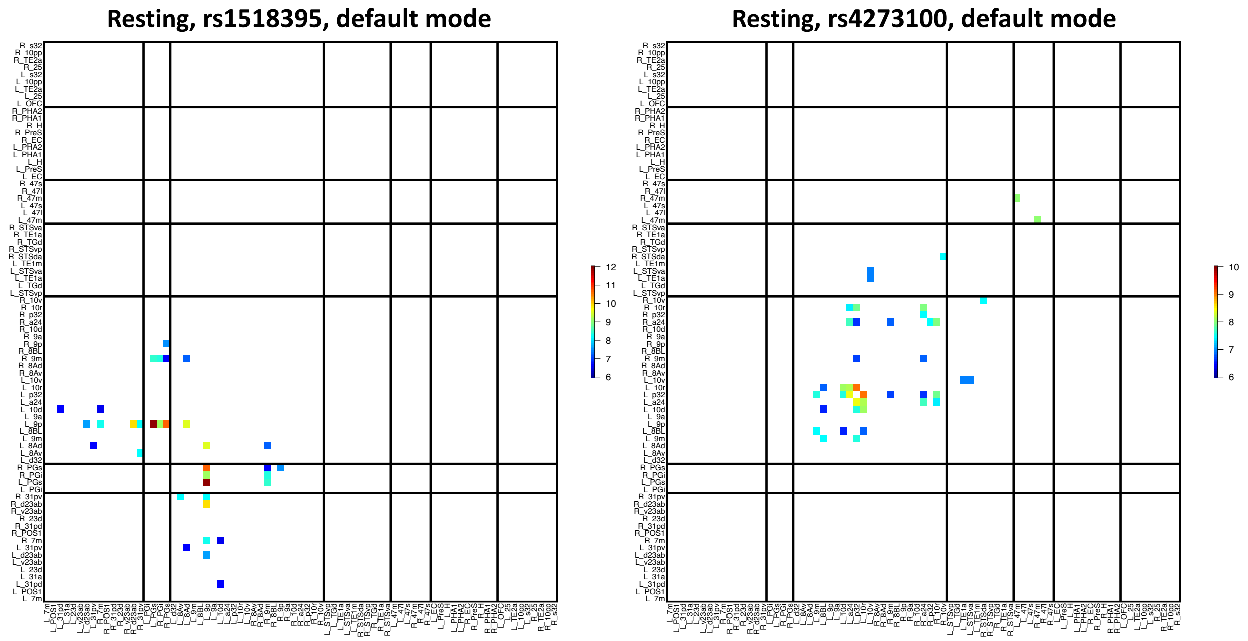


**Fig. S62 Functional areas in the default mode network associated with index genetic variants of schizophrenia (rs1518395 and rs4273100) in variant-specific analysis.**

Left: map of -$\text{Log}_{10}$(Pvalue) for significant associations at Bonferroni significance level ($3.86 \times 10^{-7}$, 0.05/64,620/2) with rs1518395. Right: map of -$\text{Log}_{10}$(Pvalue) for significant associations at Bonferroni significance level ($3.86 \times 10^{-7}$) with rs4273100. The location of the functional areas and subclusters of the default mode network can be found in Figure S9.

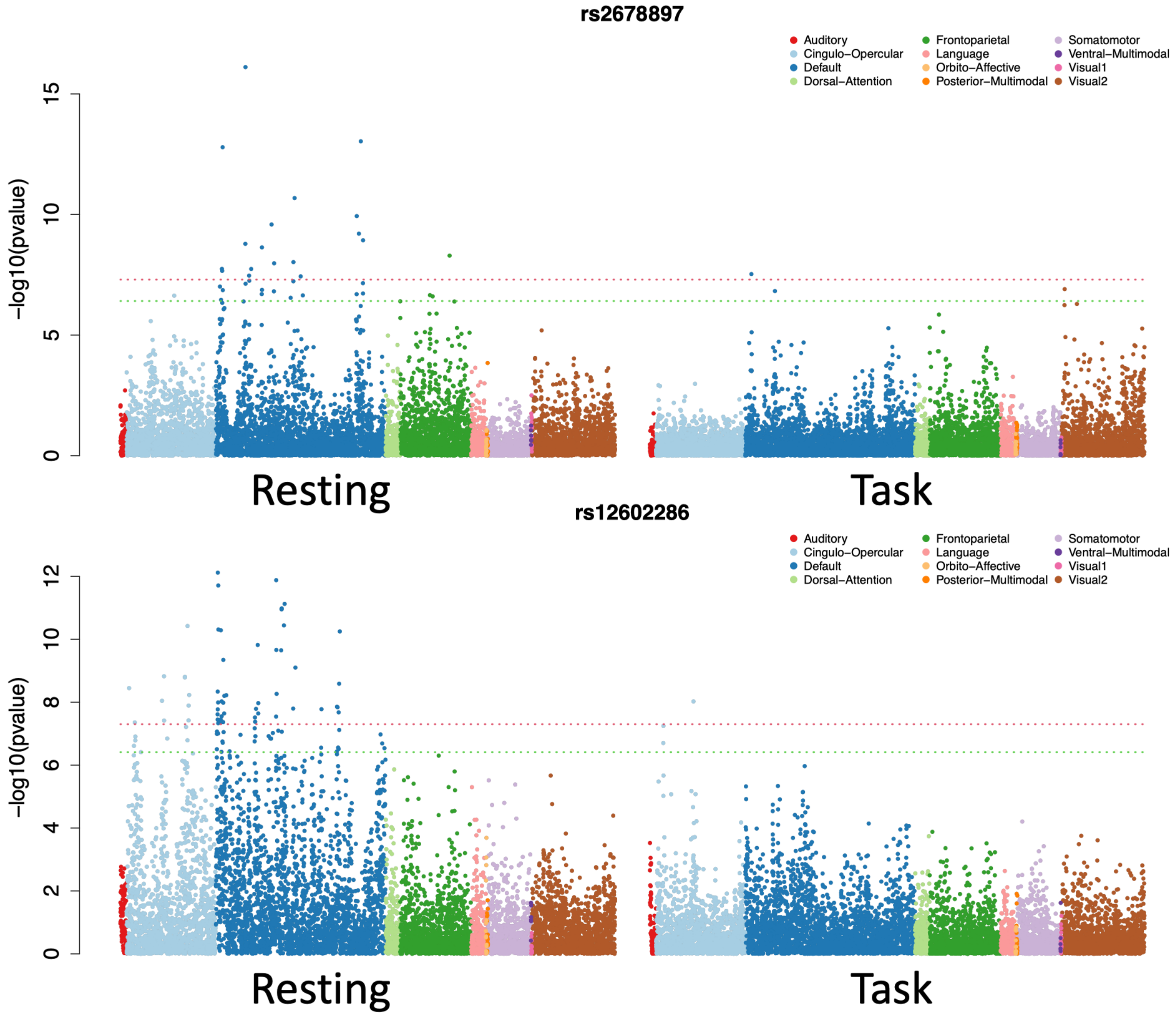


**Fig. S63 Associations between fMRI traits and the index genetic variants of cognitive ability and psychological traits (rs2678897 and rs12602286).**

We performed variant-specific association analysis with rs2678897 and rs12602286 in both resting and task fMRI. The red and green lines indicate the genome-wide significant level $5 \times 10^{-8}$ and the Bonferroni significance level of this variant-specific analysis ($3.86 \times 10^{-7}$, 0.05/64,620/2), respectively.

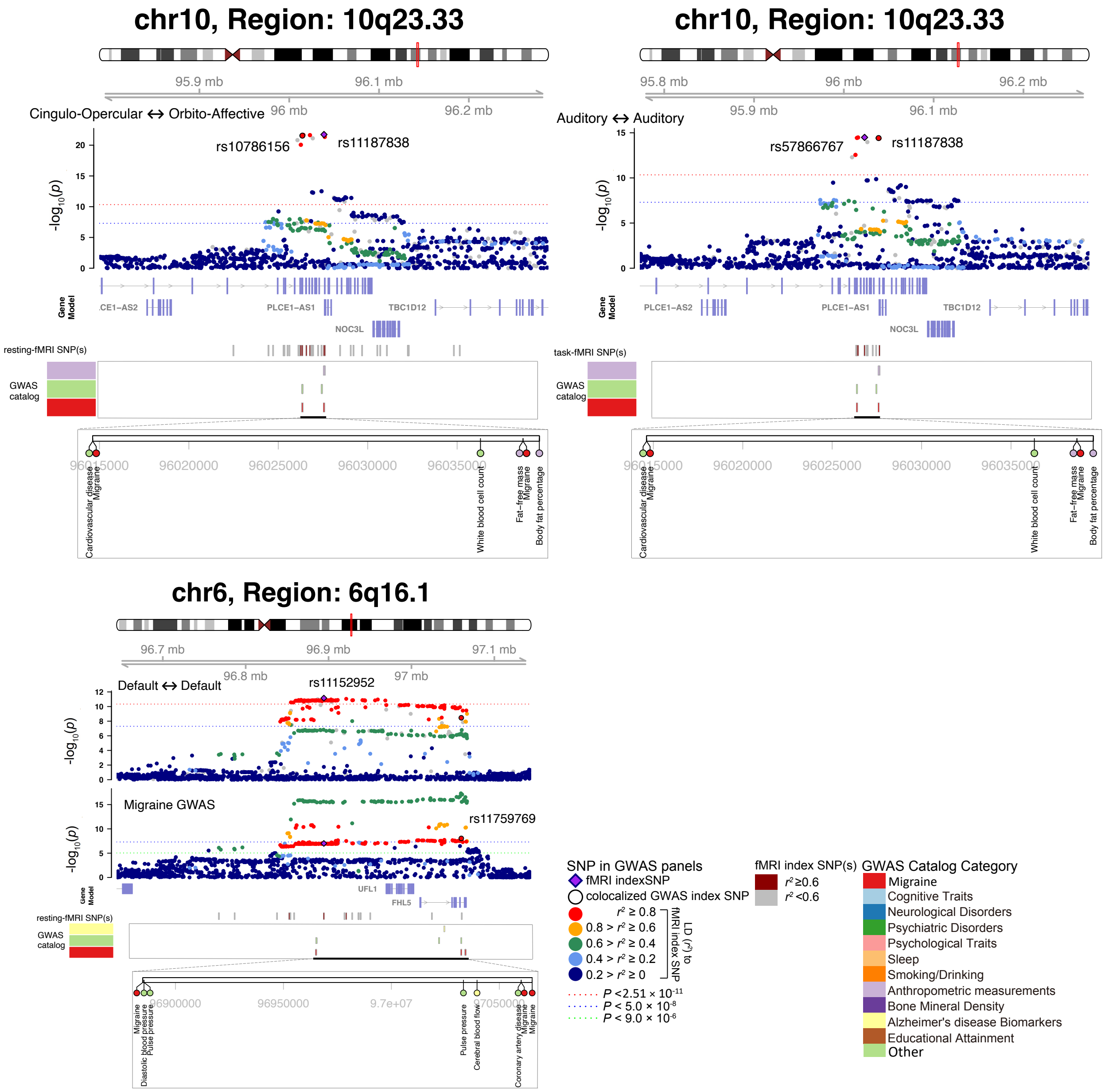


**Fig. S64 Selected genetic locus that were associated with both fMRI traits and migraine.**

In 10q23.33, we observed shared genetic influences (LD $r^2 \geq 0.6$) between both resting (left) and task (right) fMRI traits (index variants rs10786156 and rs57866767) and migraine (index variant rs11187838). In the 6q16.1 region, we observed shared genetic influences (LD $r^2 \geq 0.6$) between migraine (index variant rs11759769) and the functional connectivity of the default mode network (Default <-> Default, index variant rs11152952) in resting fMRI. Cingulo-Opercular <-> Cingulo-Opercular, functional connectivity within the cingulo-opercular network; Auditory <-> Auditory, functional connectivity within the auditory network; Default <-> Default, functional connectivity within the default mode network.

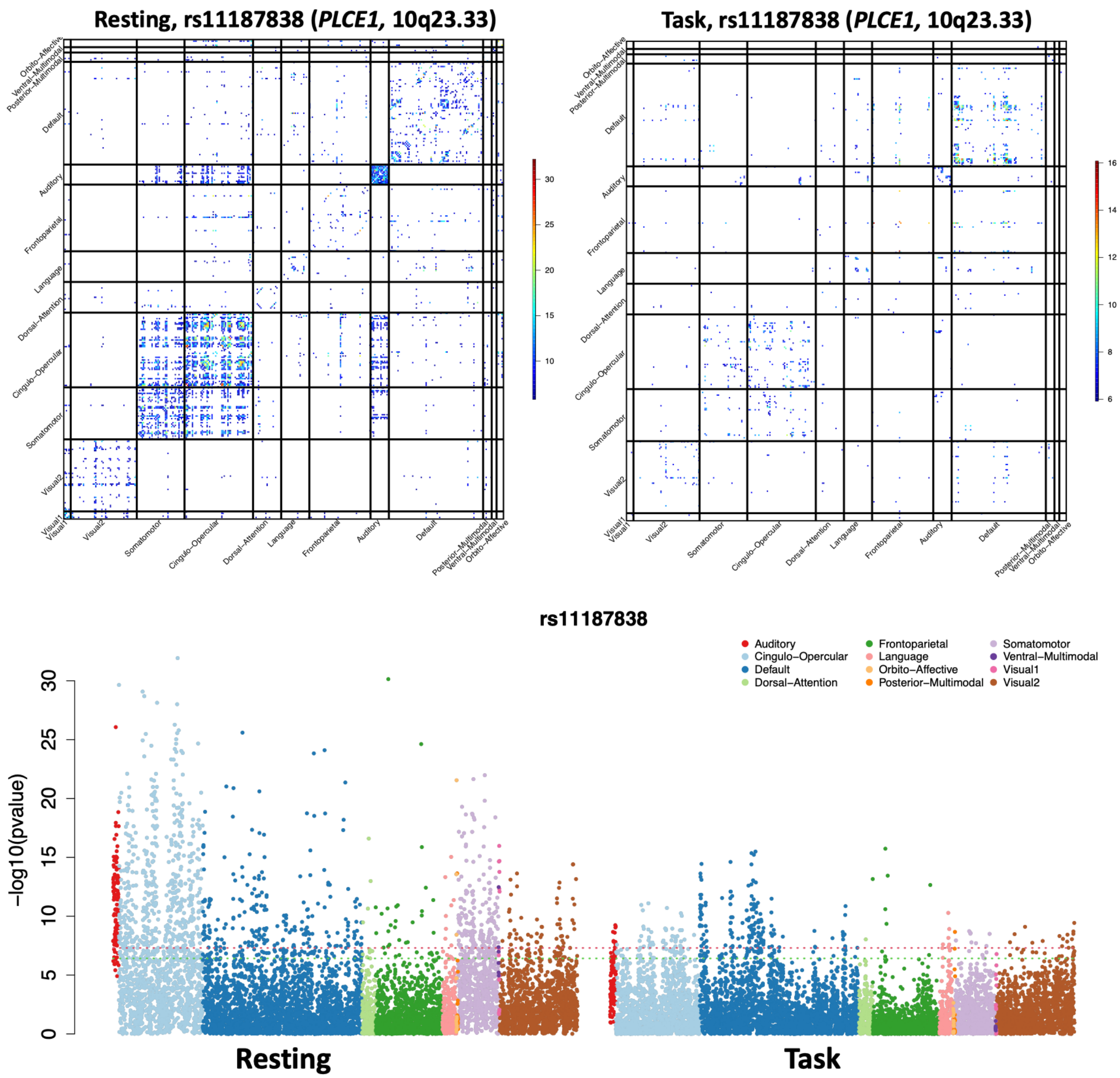


**Fig. S65 Associations between fMRI traits and the index genetic variant of migraine (rs11187838).**

We performed variant-specific association analysis with rs11187838 in both resting and task fMRI. In the bottom panel, the red and green lines indicate the genome-wide significant level $5 \times 10^{-8}$ and the Bonferroni significance level of this variant-specific analysis ($3.86 \times 10^{-7}$, 0.05/64,620/2), respectively. In the top panel, we show the -$Log_{10}$(Pvalue) map for significant associations at Bonferroni significance level ($P < 3.86 \times 10^{-7}$).

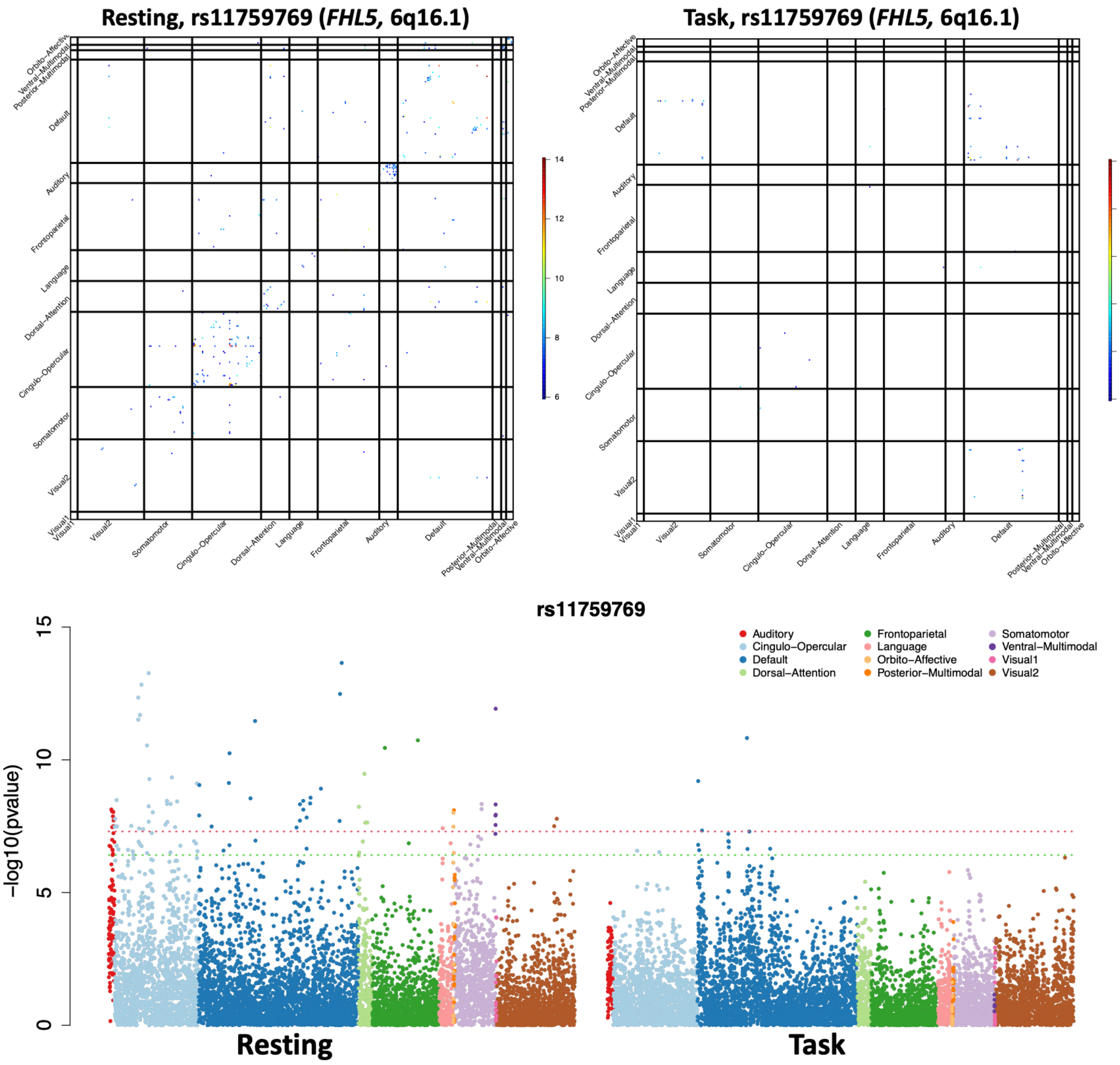


**Fig. S66 Associations between fMRI traits and the index genetic variant of migraine (rs11759769).**

We performed variant-specific association analysis with rs11759769 in both resting and task fMRI. In the bottom panel, the red and green lines indicate the genome-wide significant level $5 \times 10^{-8}$ and the Bonferroni significance level of this variant-specific analysis ($3.86 \times 10^{-7}$, 0.05/64,620/2), respectively. In the top panel, we show the -$Log_{10}$(Pvalue) map for significant associations at Bonferroni significance level ($P < 3.86 \times 10^{-7}$).

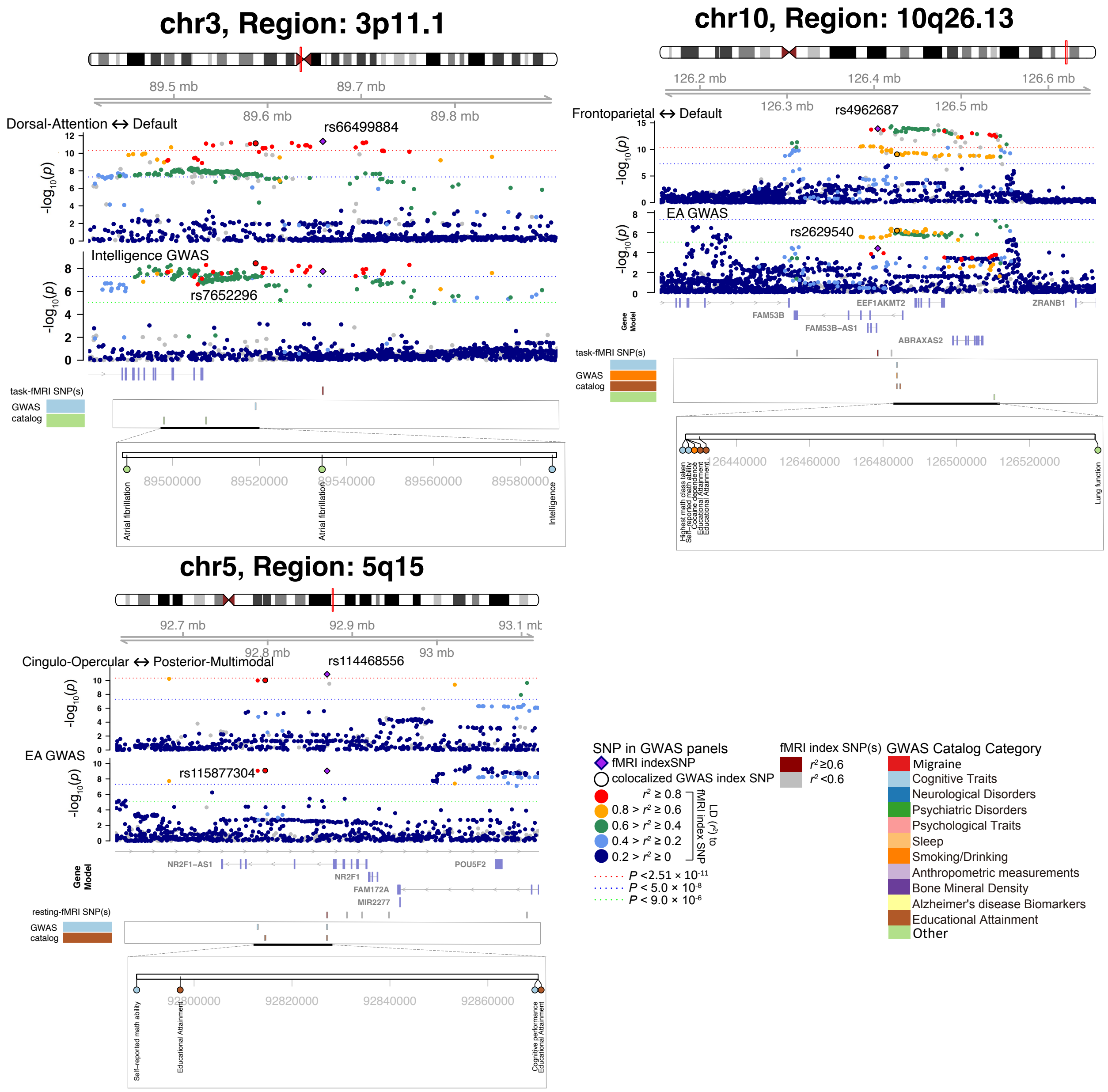


**Fig. S67 Selected genetic loci that were associated with both fMRI traits and cognitive traits.**

In 3p11.1, we observed shared genetic influences (LD $r^2 \geq 0.6$) between task-evoked fMRI trait (index variant rs66499884) and intelligence (index variant rs7652296). In 10q26.13, we observed colocalization between task-evoked fMRI trait (index variant rs4962687) and education (index variant rs2629540). In 5q15, we observed colocalization between resting-state fMRI trait (index variant rs114468556) and education (index variant rs115877304). Dorsal-Attention <-> Default, functional connectivity between the dorsal attention and default mode networks; Frontoparietal <-> Default, functional connectivity between the frontoparietal and the default mode networks; Cingulo-Opercular <-> Posterior-Multimodal, functional connectivity between the cingulo-opercular and the default mode networks.

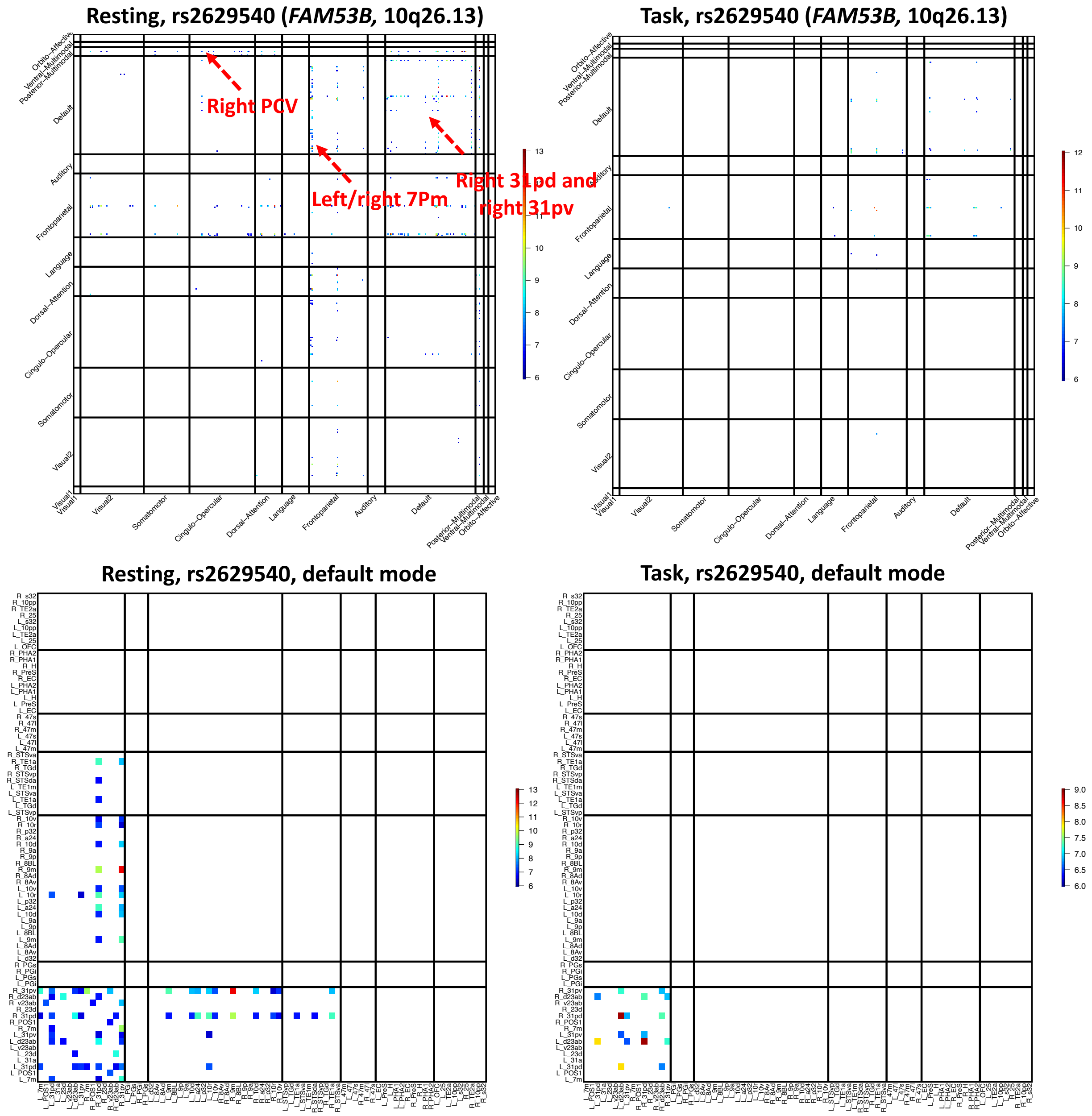


**Fig. S68 Functional areas associated with index genetic variant of education (rs2629540) in variant-specific analysis.**

We show the map of -Log$_{10}$(Pvalue) for significant associations at Bonferroni significance level ($3.86 \times 10^{-7}$, 0.05/64,620/2) with rs2629540 in both resting and task fMRI. The location of the functional areas and subclusters of the default mode network can be found in Figure S9.

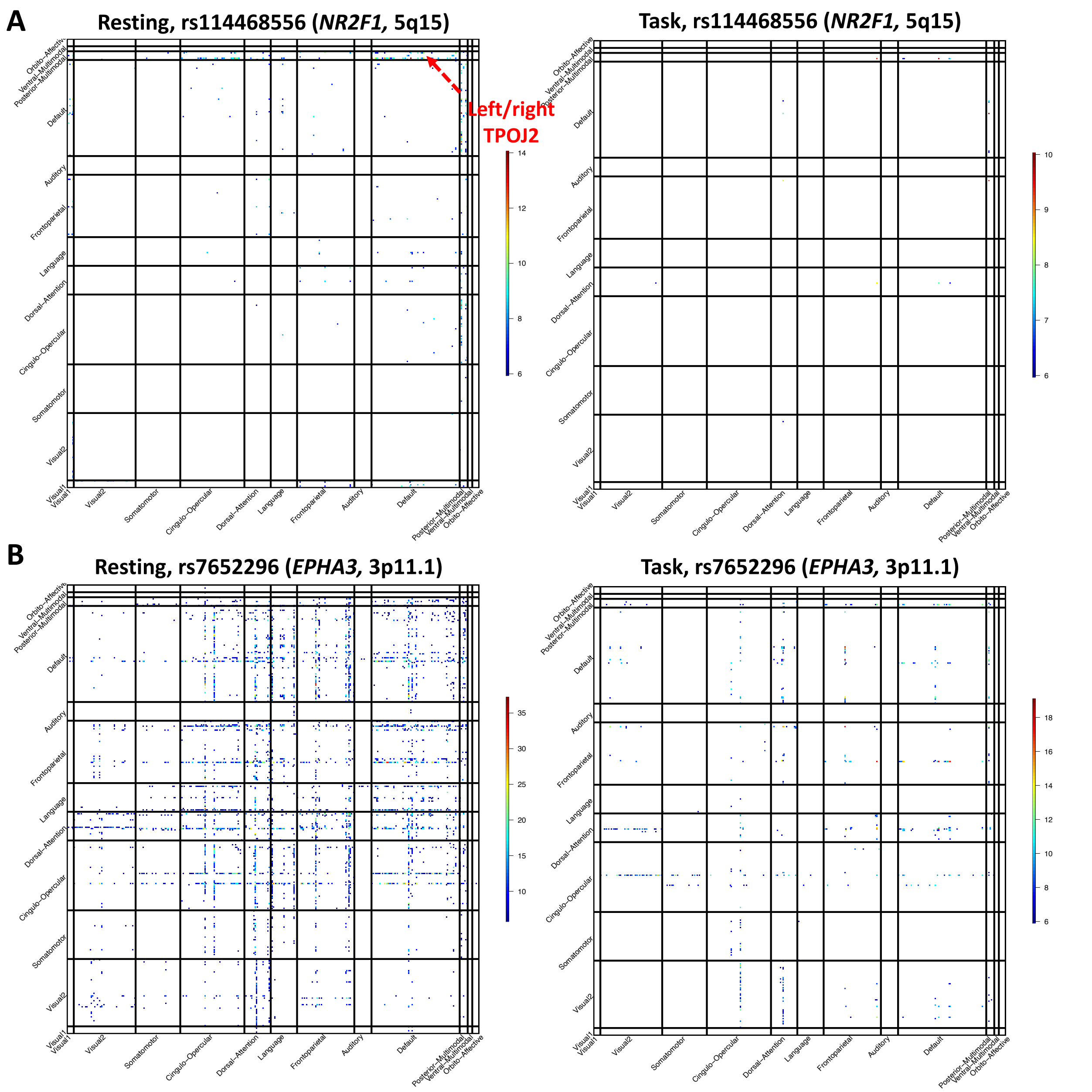


**Fig. S69 Functional areas associated with index genetic variant of education and intelligence (rs114468556 and rs7652296) in variant-specific analysis.**

We show the map of -Log$_{10}$(Pvalue) for significant associations at Bonferroni significance level ($3.86 \times 10^{-7}$, 0.05/64,620/2) with rs114468556 (upper panel) and rs7652296 (lower panel) in both resting and task fMRI.

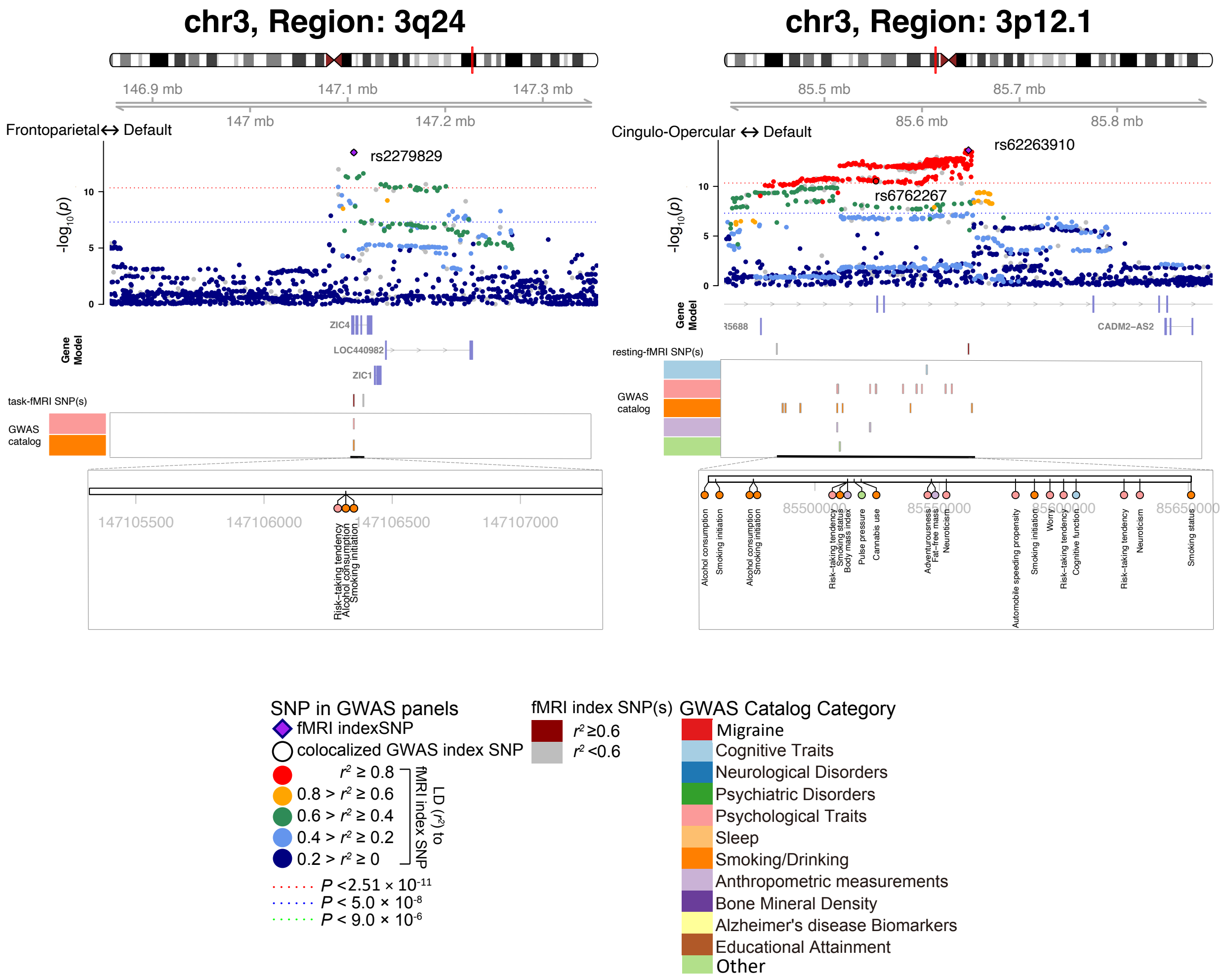


**Fig. S70 Selected genetic loci that were associated with both fMRI traits and risk-taking traits.** In 3p24, we observed shared genetic influences (LD $r^2 \geq 0.6$) between risk-taking (index variant rs2279829) and task-evoked fMRI trait (index variant rs2279829). In 3p12.1, we observed shared genetic influences between risk-taking (index variant rs6762267) and resting-state fMRI trait (index variant rs62263910).

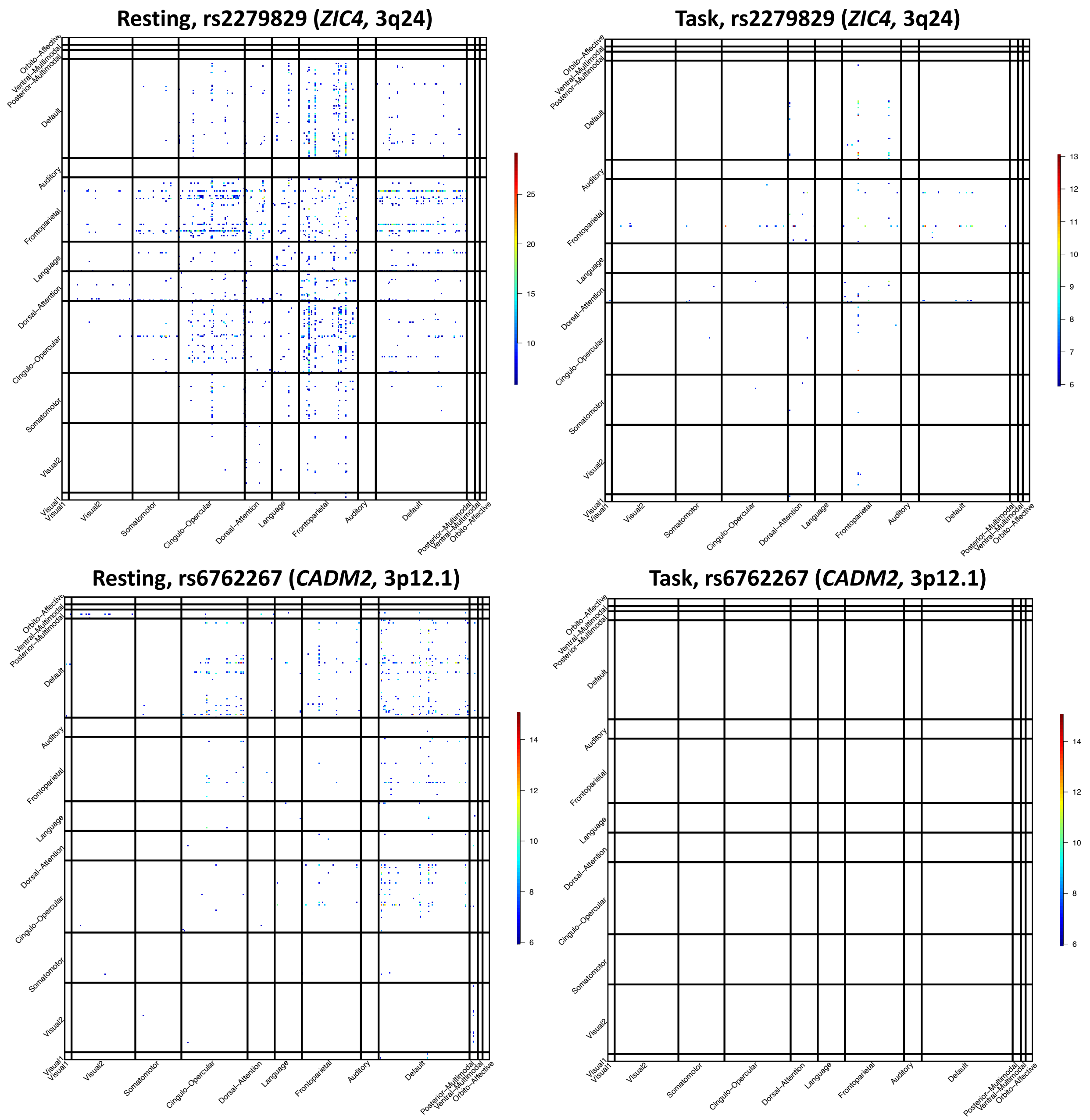


**Fig. S71 Functional areas associated with index genetic variant of risk-taking (rs2279829 and rs6762267) in variant-specific analysis.**

We show the map of -Log$_{10}$(Pvalue) for significant associations at Bonferroni significance level ($3.86 \times 10^{-7}$, 0.05/64,620/2) with rs2279829 (upper panels) and rs6762267 (lower panels) in both resting and task fMRI.

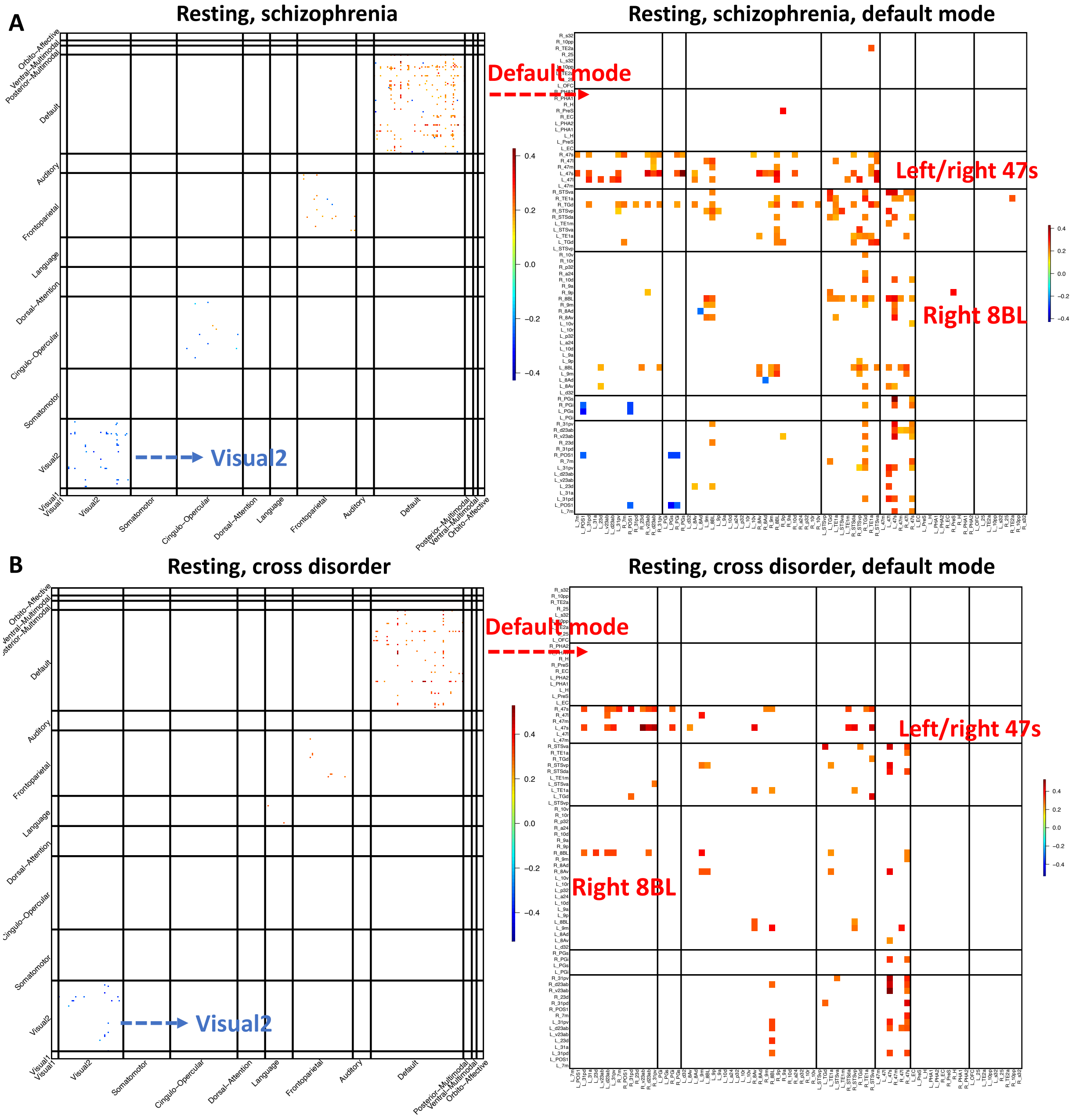


**Fig. S72 Significant genetic correlations with schizophrenia and cross-disorder.**

We show the map of significant genetic correlations at the FDR 5% level (8,351 tests) with schizophrenia (**A**) and cross-disorder (**B**) in resting fMRI. The location of the functional areas and subclusters of the default mode network can be found in Figure S9.

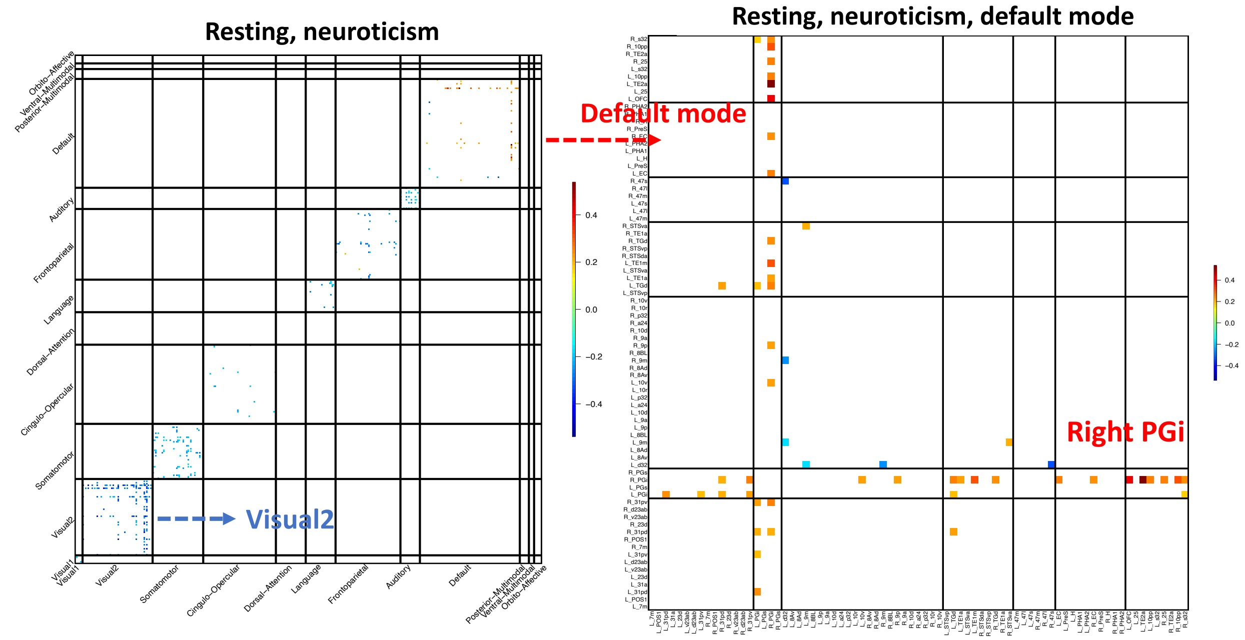


**Fig. S73 Significant genetic correlations with neuroticism (feeling nervous).**

We show the map of significant genetic correlations at the FDR 5% level (8,351 tests) with neuroticism in resting fMRI. The location of the functional areas and subclusters of the default mode network can be found in Figure S9.

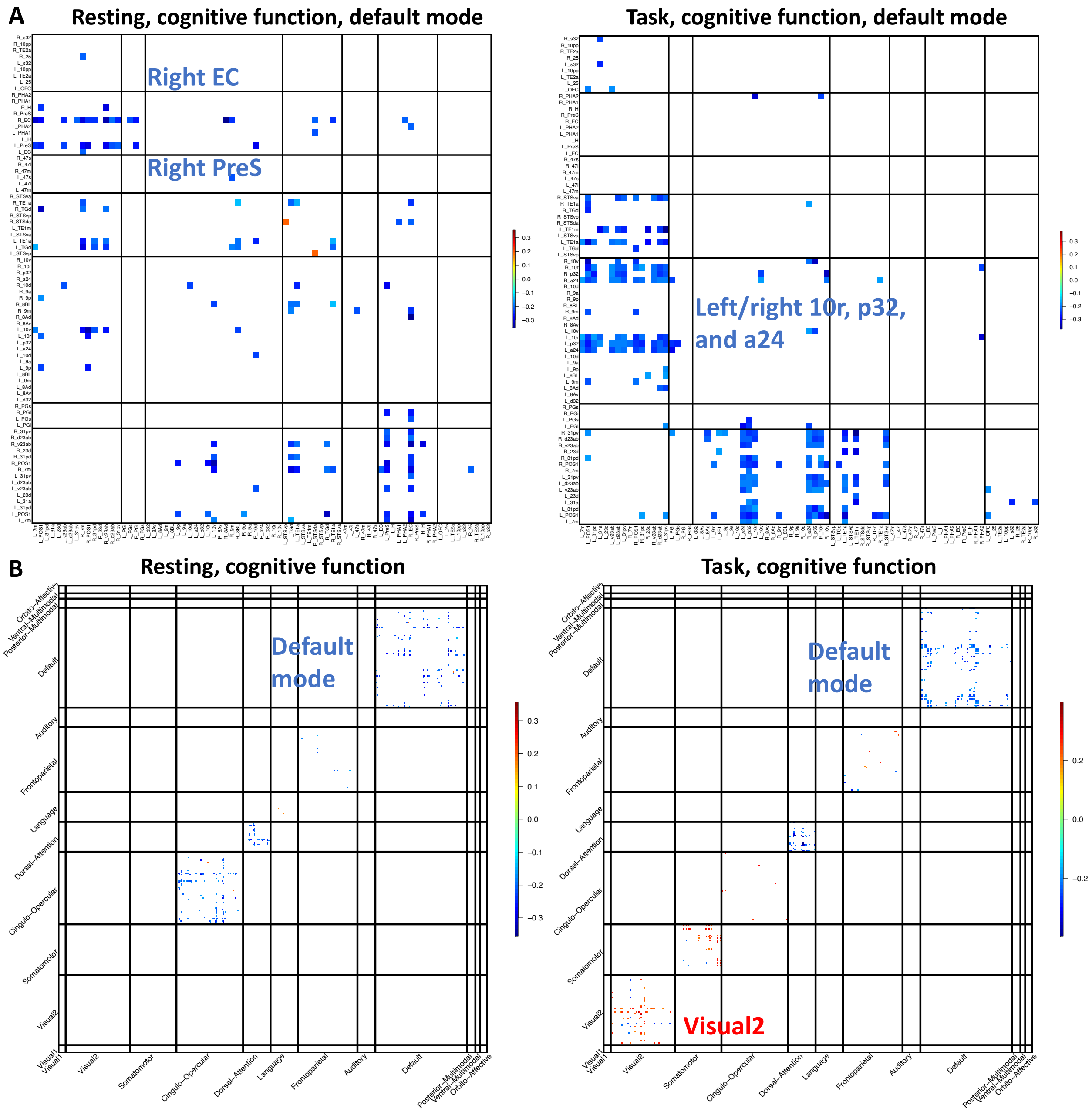


**Fig. S74 Significant genetic correlations with cognitive function.**

We show the map of significant genetic correlations at the FDR 5% level (8,351 tests) with neuroticism in both resting and task fMRI. **A:** the significant genetic correlations within the default mode network. Location of the functional areas and subclusters of default mode network can be found in Figure S11. **B:** the significant genetic correlations across different networks.

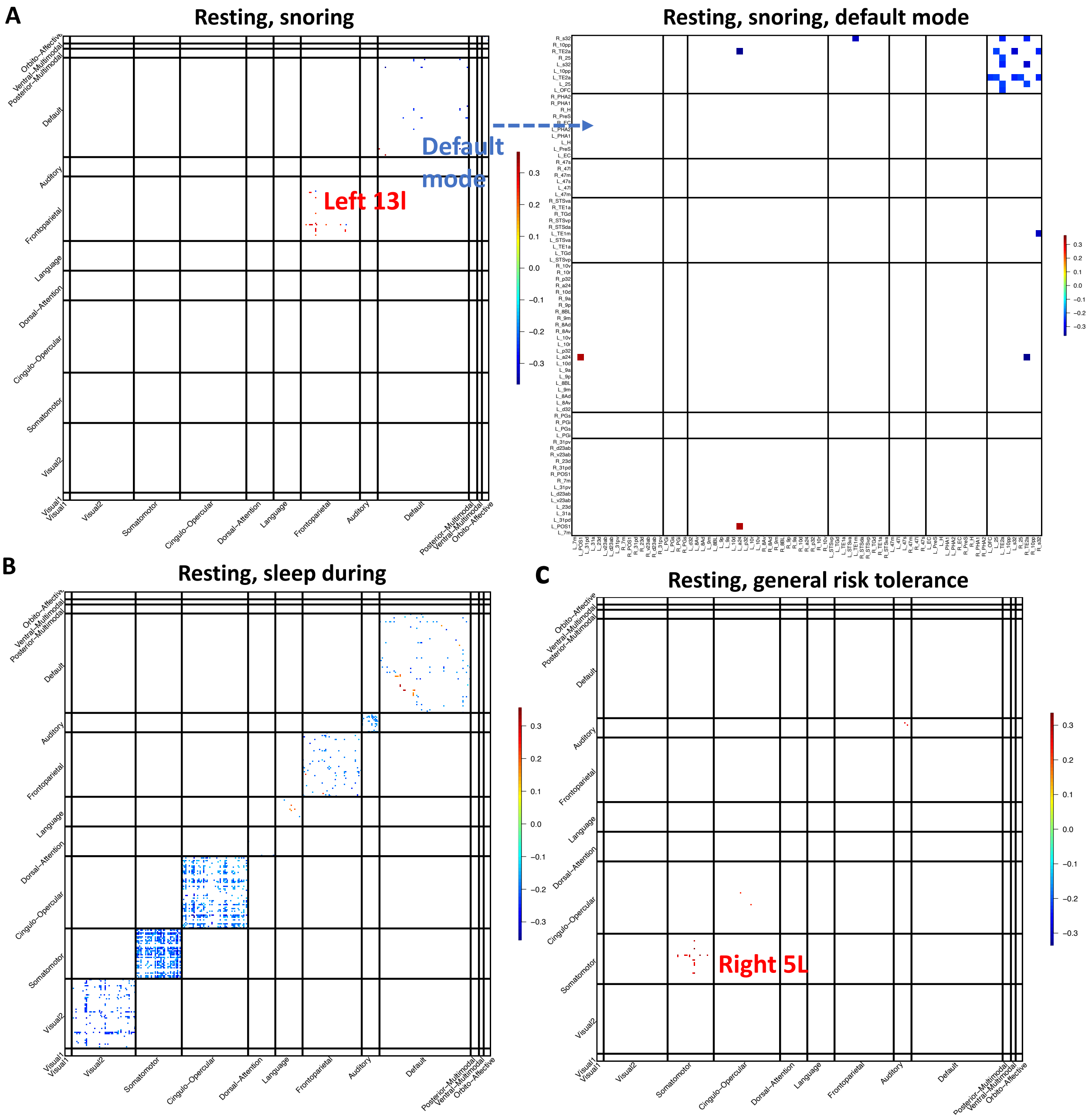


**Fig. S75 Significant genetic correlations with snoring, sleep during, and general risk tolerance.**

We show the map of significant genetic correlations at the FDR 5% level (8,351 tests) with snoring (**A**), sleep during (**B**), and general risk tolerance (**C**) in resting fMRI. The location of the functional areas and subclusters of the default mode network can be found in Figure S9.

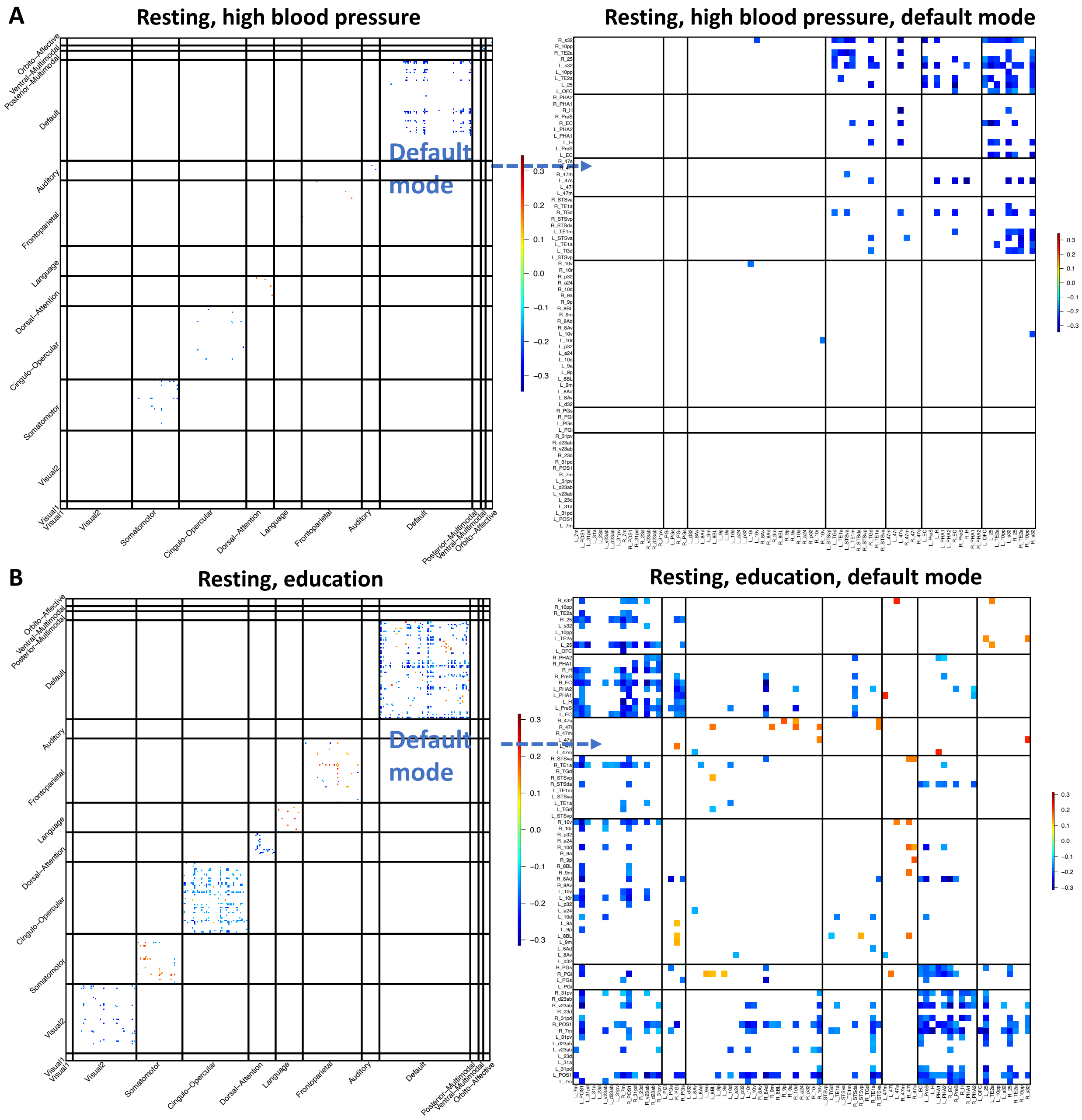


**Fig. S76 Significant genetic correlations with high blood pressure and education.**

We show the map of significant genetic correlations at the FDR 5% level (8,351 tests) with high blood pressure (**A**) and education (**B**) in resting fMRI. The location of the functional areas and subclusters of the default mode network can be found in Figure S9.

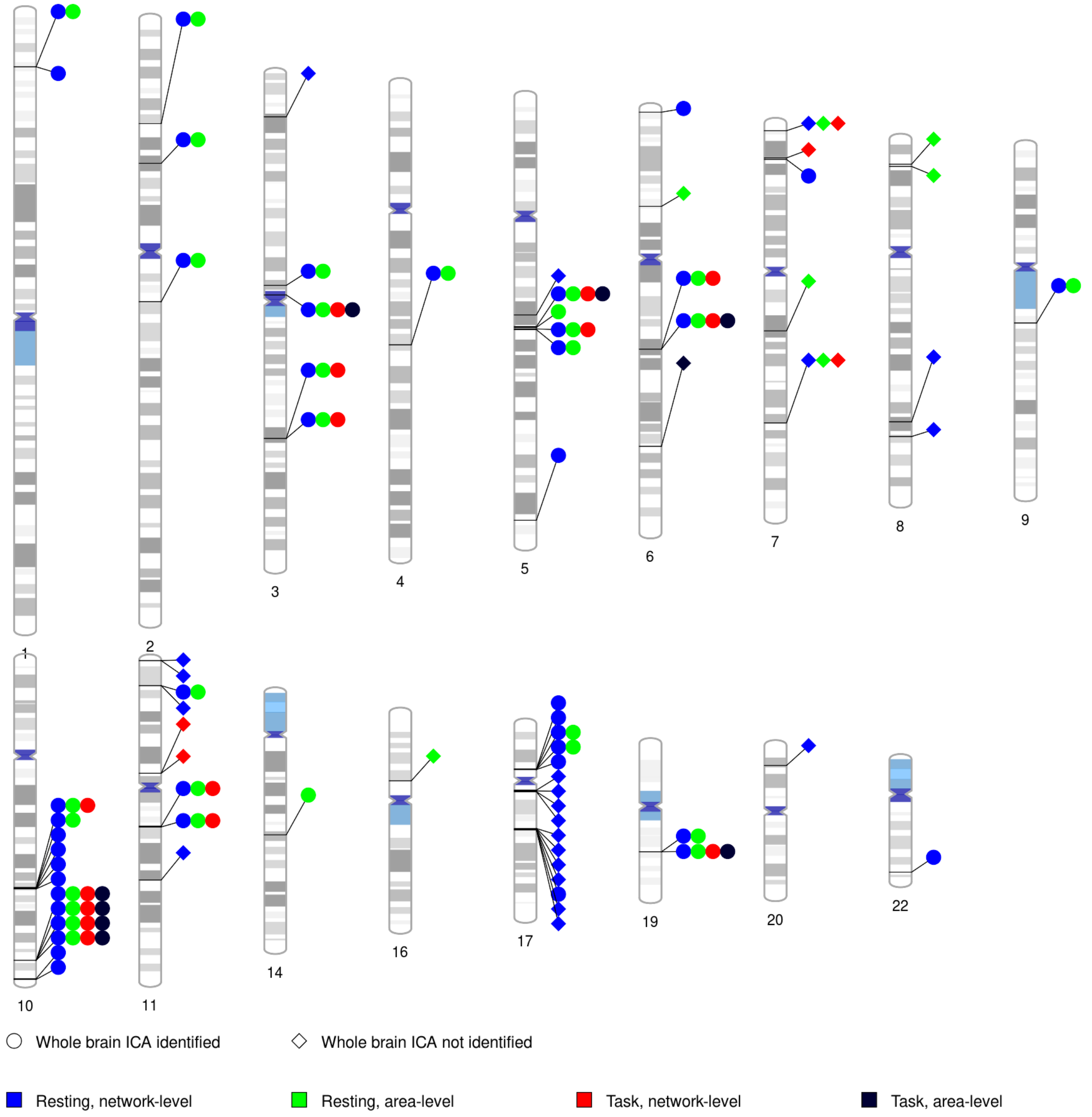


**Fig. S77 MAGMA-significant genes associated with fMRI traits.**

There were 76 MAGMA-significant genes associated with area- or network-level fMRI traits, 45 of which have been previously identified regions in the whole brain ICA analysis. Each signal dot indicates that at least one of the fMRI traits of this category (network-level traits in resting fMRI, network-level traits in task fMRI, area-level traits in resting fMRI, or area-level traits in task fMRI) was associated with this gene.

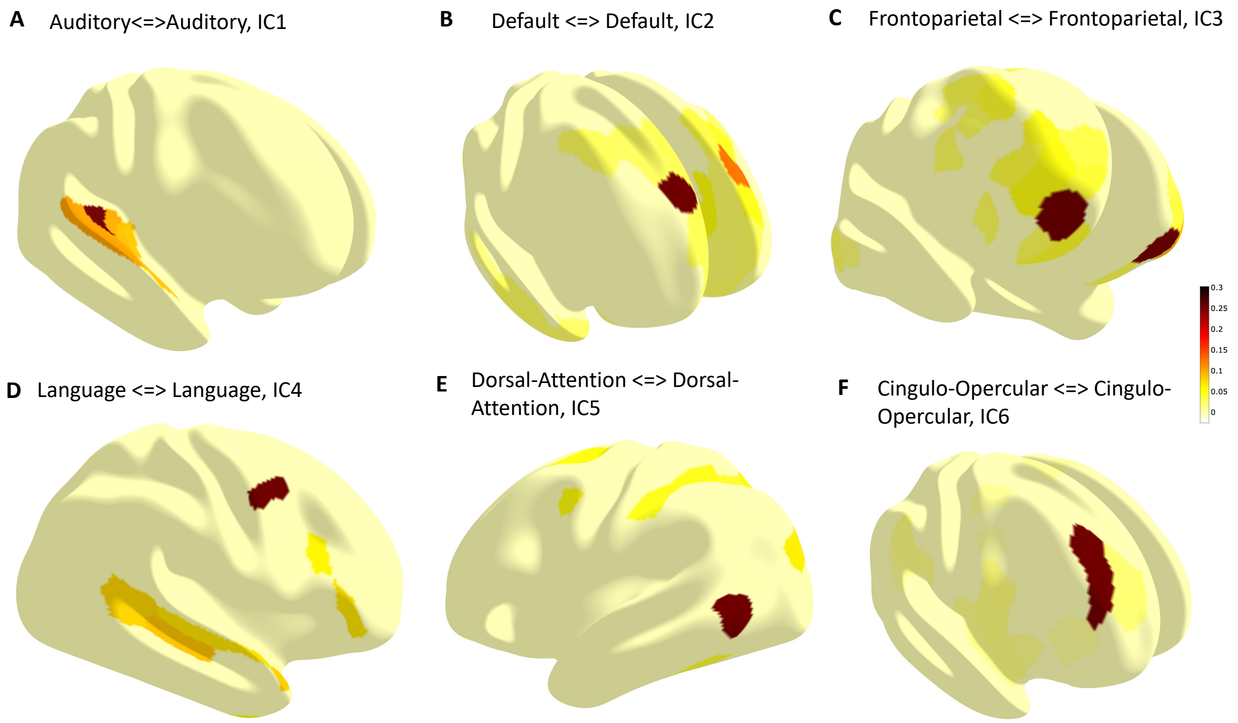


**Fig. S78 Examples of mapping network-level ICA components to functional areas in the Glasser 360 atlas.**

The color indicates the ICA weights. The full set of 3D illustrations of all ICA components can be downloaded at: https://www.dropbox.com/s/np0rjlzvfa4fgae/Resting_fMRI_ICA.zip?dl=0 (for resting fMRI and https://www.dropbox.com/s/rg053royl62z12v/Task_fMRI_ICA.zip?dl=0 (for task fMRI).
